# Building a prediction model for outcomes following treatment in UK NHS Talking Therapies services for depression and anxiety

**DOI:** 10.64898/2026.03.12.26348223

**Authors:** Nour Kanso, Megan Skelton, Katharine A. Rimes, Grace Wong, Thalia C. Eley, Ewan Carr

## Abstract

**Background:** Depression and anxiety are common mental health conditions in the UK. NHS Talking Therapies offers evidence-based therapies and is the largest provider of treatment, yet, only 50% of patients recover. Accurate outcome prediction could identify those at risk of poor outcomes and support more personalised care. This study aimed to develop and internally validate multivariable prediction models using routinely collected data from a large, ethnically diverse sample to enable fair, data-driven treatment decisions.

**Methods:** Data included 30,999 adults who completed high-intensity therapy at a single NHS trust between 2018 and mid-2024. Seven NHS post-treatment outcomes were modelled: reliable improvement, recovery, and reliable recovery for both depression and anxiety, and also functional impairment at the end of treatment. Predictors measured at baseline included sociodemographic and clinical characteristics. Models were developed using elastic net logistic regression and internally validated using bootstrap resampling.

**Results:** The sample was predominantly female (73%) with a median age of 34; 57% identified as White and 22% as Black. Models showed moderate to good discrimination (AUC 0.63–0.77) and strong calibration. Key predictors aligned with clinical expectations, including baseline symptom severity, unemployment, benefit receipt, reporting a disability or long-term condition, psychotropic medication use among other sociodemographic factors.

**Conclusions:** This study highlights the potential of data-driven tools to inform clinical decisions and treatment stratification in NHS Talking Therapies. Early identification of patients less likely to benefit from standard care could support timely review, monitoring, or tailored interventions. External validation and implementation research are needed to ensure generalisability and equity in care.

## Background

Depression and anxiety are among the most prevalent psychiatric conditions, affecting 1 in 6 adults aged 16 or older in England each week [1, 2]. Both conditions are multifactorial, shaped by psychological, environmental, and genetic influences, and are associated with substantial individual and societal burden, including poorer physical health, increased disability, and higher healthcare costs [3].

Psychological talking therapies effectively treat common mental health problems. [4]. In 2008, the UK National Health Service (NHS) commissioned the Improving Access to Psychological Therapies (IAPT) programme, now known as NHS Talking Therapies (NHS-TT) to increase access to NICE (National Institute for Health and Care Excellence)-recommended interventions [5]. Now widely available, NHS-TT saw over 1.26 million referrals in 2024. Of patients referred, approximately 50% meet NHS thresholds for recovery and around 60% show reliable symptom improvement [6]. These figures highlight significant variability in outcomes following NHS-TT: half of patients do not meet definitions of recovery by the end of their treatment. Individuals with the same clinical diagnosis may respond differently to psychological therapy due to their unique characteristics and circumstances [7]. Personalised or stratified care has gained attention as an approach to improve treatment outcomes, which involves predicting treatment outcomes using sociodemographic and clinical variables collected at baseline to identify individuals at risk of non-response who may require adapted or alternative treatments [8].

A growing body of work has examined associations of treatment trajectories or endpoint outcomes for patients receiving NHS-TT treatment. Lower baseline symptom severity, fewer comorbidities (e.g., phobic symptoms), and higher baseline functioning have been consistently associated with better treatment outcomes [9–11]. Employed patients are also more likely to experience improvement [7, 11]. Conversely, poorer outcomes have been linked to higher symptom severity, greater functional impairment, the presence of disability, and the use of prescribed psychotropic medication [7, 9, 11].

While informative, much of this literature focuses on group-level associations rather than individual-level prediction. Many previous studies have also been conducted in predominantly White samples, limiting generalisability. This is clinically significant, as ethnicity is associated with differences in access, experience, and outcomes within psychological services, raising concerns that models developed in homogeneous samples may perform less well or less equitably in routine care [12, 13]. Although more recent work has begun to include more ethnically representative NHS-TT samples, these studies are often constrained by modest effective sample sizes or omit key evaluation metrics such as calibration [11, 14]. Calibration reflects the agreement between predicted risks and observed outcomes and is critical for clinical use where even small miscalibrations can meaningfully affect decisions [15]. In the context of psychological services, this may mean decisions about treatment intensity, monitoring, or referral.

For clinical application, predictors must be integrated into multivariable models capable of generating reliable, individualised risk estimates. Prediction models use statistical or machine-learning approaches to estimate outcomes for new patients based on baseline characteristics [16]. However, clinical prediction research often faces methodological issues like small sample sizes and poor validation, leading to overfitting and inflated performance. As a result, models performing accurately during development may perform substantially worse when applied to new patients, undermining clinical trust and limiting safe implementation [17].

The present study aims to address the gaps of previous literature by developing and internally validating robust multivariable prediction models in a large, ethnically diverse NHS-TT sample. We leverage routinely collected baseline data and recent improvements in data completeness [18] to build models that prioritise both discrimination and calibration. Establishing well-calibrated models in diverse samples is a critical step towards developing tools that are not only accurate, but also equitable and suitable for real-world clinical use. By identifying patients at risk of poorer outcomes, such models would enable more effective treatment planning and directing resources to those most in need based on risk. Such insights can inform healthcare policies and drive targeted strategies to enhance the quality and equity of care.

## Methods

### Sample

We used the Clinical Record Interactive Search (CRIS; [19]) to extract information for patients (N = 155,945) who received a course of NHS-TT psychological therapy under the South London and Maudsley NHS Foundation Trust (SLaM) between January 1, 2018, and August 27, 2024. SLaM covers four boroughs: Lambeth, Southwark, Lewisham, and Croydon. The included patients were aged 18 years or older to maintain consistency with standard practice guidelines and to focus on adult populations. The six-year extraction period ensured both adequate sample size and high-quality data, benefiting from improvements in recording patient information within NHS-TT since 2014 [20].

### Data cleaning

**Supplementary Figure 1** outlines the data processing steps. NHS-TT services offer low and high-intensity treatments, but to increase treatment homogeneity, we restricted the sample to high-intensity therapy only. High-intensity treatment refers to structured, therapist-delivered interventions, such as cognitive behavioural therapy, typically offered to individuals with moderate to severe symptoms of depression or anxiety on a weekly basis [21, 22]. To accurately determine therapy intensity, raw data on intervention types were mapped to treatment types specified in the NHS-TT Data Dictionary.

We restricted the sample to patients attending 3-21 sessions to ensure recorded outcomes reflected response to therapy. Requiring a minimum of 3 sessions ensures post-treatment scores reflect some change after intervention, as the first two high-intensity sessions are often for assessment or planning. Limiting to 21 sessions excludes atypical, extremely long episodes unlikely to be a standard course of therapy, balancing clinical relevance with representativeness across typical treatment lengths [23].

After data cleaning, the analytical sample size was reduced from 155,945 to 30,999 patients who completed 3-21 high-intensity treatment sessions. The largest reductions came from excluding patients who received only low-intensity treatment, or attended fewer than three sessions. Additional exclusions included patients with missing baseline depression or anxiety scores, those whose primary diagnosis was recorded as “Non-IAPT” (typically employment-related concerns), and those with exceptionally high session counts likely indicating miscoded follow-up episodes. These steps ensured a more consistent and clinically relevant sample. **Supplementary Table 1** and **Supplementary Figure 2** summarise the number of sessions per patient and their distributions. The specific high-intensity interventions received by patients are summarised in **Supplementary Table 2.**

### Outcomes

Although NHS-TT treats patients with a wide range of diagnoses, outcomes are routinely monitored symptom scales for depression and anxiety measured by the Patient Health Questionnaire (PHQ-9; [24]) and Generalised Anxiety Disorder Scale (GAD-7; [25]), respectively. These instruments are used as standard across services for service-level reporting and performance monitoring, allowing consistent measurement of treatment response across a heterogeneous patient population. We considered three outcomes each for depression and anxiety:

1. Reliable improvement: A change exceeding the measurement error of the scale (≥6 and ≥4 points, respectively, for PHQ-9 and GAD-7) [26, 27].
2. Recovery: A measure that considers baseline caseness (≥10 and ≥8 for PHQ-9 and GAD-7, respectively). Recovery is achieved when those who are cases at baseline score below these thresholds by the end of treatment [26].
3. Reliable recovery: Combines both criteria (1) and (2) and requires that patients who are cases at baseline score below the clinical threshold at the end of treatment *and* show symptom reduction exceeding scale error (≥6 points for PHQ-9 and ≥4 points for GAD-7; [26, 27]).

We also assessed functional impairment as an outcome. Although not a NHS standard outcome, previous work has suggested that patient functioning is somewhat independent of symptoms of anxiety or depression [28].

4. Functional impairment: Scoring ≥10 on the Work and Social Adjustment Scale (WSAS; [29, 30]) at the end of treatment, irrespective of their status at baseline.

### Candidate predictors

We considered a range of patient sociodemographic and clinical factors at baseline. Sociodemographic predictors included age, gender, religion, sexual orientation, ethnicity, English language proficiency, Index of Multiple Deprivation (IMD) decile, receipt of benefits, Statutory Sick Pay (SSP), and employment status (see **Table 1** for categories, and see **Supplementary Table 3** for descriptions and how data was measured). Clinical predictors included the presence of a long-term condition, disability, psychotropic medication usage, previous NHS-TT referrals, primary diagnosis, and symptom severity of anxiety, depression, social phobia, agoraphobia, specific phobia, and functional impairment (see **Table 2** for categories, and see **Supplementary Table 4** for descriptions and how data was measured).

**Table 1:** Baseline sociodemographic characteristics of the sample (N= 30,999)

| **Sociodemographic Characteristic** | **Category** | **Count** | **Valid Percentage (%)<sup>\*</sup>** |
| --- | --- | --- | --- |
| **Age** | Mean (SD) | 37.31 (13.12) | - |
|  | Median (IQR) | 34 (27–46) | - |
|  | Range | 18–94 | - |
| **IMD Decile (2015)** | Mean (SD) | 3.75 (1.90) | - |
|  | Median (IQR) | 3 (2–5) | - |
|  | Range | 1–10 | - |
|  | Unknown (%)** | 8616 (27.79) | - |
| **Ethnicity** | **White** | **17011** | **57.15** |
|  | Black, Black British, Caribbean, or African | 6645 | 22.33 |
|  | Mixed or multiple ethnicities | 2713 | 9.12 |
|  | Asian or Asian British | 1985 | 6.67 |
|  | Other ethnic group | 1410 | 4.70 |
|  | Unknown (%)** | 1235 (3.98) | - |
| **Gender** | **Woman (cisgender + transgender)** | **22441** | **72.53** |
|  | Man (cisgender + transgender) | 8371 | 27.06 |
|  | Non-binary | 128 | 0.41 |
|  | Unknown (%)** | 59 (0.19) | - |
| **Sexual Orientation** | **Straight** | **23729** | **86.39** |
|  | Bisexual | 1631 | 5.94 |
|  | Lesbian/Gay | 1553 | 5.65 |
|  | Questioning | 263 | 0.96 |
|  | Queer | 238 | 0.87 |
|  | Asexual | 53 | 0.19 |
|  | Unknown (%)** | 3532 (11.39) | - |
| **Employment** | **Employed** | **18177** | **61.01** |
|  | Unemployed | 5301 | 17.79 |
|  | Unemployed due to sickness/disability | 1567 | 5.26 |
|  | Student | 1554 | 5.22 |
|  | Self-employed | 1327 | 4.45 |
|  | Retired | 1012 | 3.40 |
|  | Other | 855 | 2.87 |
|  | Unknown (%)** | 1206 (3.89) | - |
| **Religion** | **Atheist/Agnostic** | **11921** | **51.46** |
|  | Christian | 8660 | 37.38 |
|  | Muslim | 1740 | 7.51 |
|  | Other | 844 | 3.64 |
|  | Unknown(%)** | 7834 (25.27) | - |
| **Receive benefits** | Yes | 5056 | 21.75 |
|  | **No** | **18185** | **78.25** |
|  | Unknown (%)** | 7758 (25.03) | - |
| **Receive statutory sick pay** | Yes | 1692 | 5.75 |
|  | **No** | **27725** | **94.25** |
|  | Unknown (%)** | 1582 (5.10) | - |
| **Speak English** | **Yes** | **23194** | **95.97** |
|  | No | 973 | 4.03 |
|  | Unknown (%)** | 6832 (22.04) | - |
| **Read English** | **Yes** | **22674** | **95.31** |
|  | No | 1115 | 4.69 |
|  | Unknown (%)** | 7210 (23.26) | - |

Values are reported as *n* (%) unless otherwise stated. Percentages are calculated using available (non-missing) data for each variable.
\* Valid percentages exclude missing or unknown responses.
\*\* Percentages for unknown values are calculated using the full sample size (N = 30,999).
IMD = Index of Multiple Deprivation (2015).

**Table 2:** Baseline clinical characteristics of the sample (N= 30,999)

| **Clinical Characteristic** | **Category** | **Value/Count** | **Valid Percentage (%)<sup>\*</sup>** |
| --- | --- | --- | --- |
| **PHQ-9** | Mean (SD) | 14.15 (6.22) | - |
|  | Median (IQR) | 14 (10–19) | - |
|  | Range | 0–27 | - |
| **GAD-7** | Mean (SD) | 12.76 (5.25) | - |
|  | Median (IQR) | 13 (9–17) | - |
|  | Range | 0–21 | - |
| **WSAS** | Mean (SD) | 18.10 (9.49) | - |
|  | Median (IQR) | 18 (11–25) | - |
|  | Range | 0–40 | - |
|  | Unknown (%)** | 979 (3.16) | - |
| **Social Phobia** | Mean (SD) | 3.31 (2.51) | - |
|  | Median (IQR) | 3 (2–6) | - |
|  | Range | 0–8 | - |
|  | Unknown (%)** | 9297 (29.99) | - |
| **Agoraphobia** | Mean (SD) | 2.79 (2.68) | - |
|  | Median (IQR) | 2 (0–5) | - |
|  | Range | 0–8 | - |
|  | Unknown (%)** | 9329 (30.09) | - |
| **Specific Phobia** | Mean (SD) | 2.45 (2.68) | - |
|  | Median (IQR) | 2 (0–4) | - |
|  | Range | 0–8 | - |
|  | Unknown (%)** | 9345 (30.15) | - |
| **Treatment service** | **Lambeth** | **10978** | **35.41** |
|  | Lewisham | 6969 | 22.48 |
|  | Southwark | 6903 | 22.27 |
|  | Croydon | 6149 | 19.84 |
| **Number of previous referrals** | Mean (SD) | 0.19 (0.60) | - |
|  | Median (IQR) | 0 (0–0) | - |
|  | Range | 0–7 | - |
| **Primary diagnosis** | **Depressive episode** | **13694** | **46.39** |
|  | Post-traumatic stress disorder | 3834 | 12.99 |
|  | Generalised anxiety disorder | 3236 | 10.96 |
|  | Panic/phobia | 2245 | 7.60 |
|  | Reaction to severe stress and adjustment disorders | 2242 | 7.59 |
|  | Obsessive-compulsive disorder | 1598 | 5.41 |
|  | Mixed anxiety and depressive disorder | 825 | 2.79 |
|  | Other anxiety disorder | 673 | 2.28 |
|  | Other mood/personality disorder | 660 | 2.24 |
|  | Personal life events | 515 | 1.74 |
|  | Unknown (%)** | 1477 (4.76) | - |
| **Medication** | **Not prescribed** | **19436** | **65.77** |
|  | Prescribed and taking | 8943 | 30.26 |
|  | Prescribed but not taking | 1173 | 4.00 |
|  | Unknown (%)** | 1447 (4.67) | - |
| **≥ 1 Long-term condition\*\*\*** | Yes | 8036 | 25.92 |
|  | No | **22963** | 74.08 |
| **≥ 1 Disability\*\*\*** | Yes | 3652 | 11.78 |
|  | No | **27347** | 88.22 |

Values are reported as mean (standard deviation), median (interquartile range), range, or *n* (%) as appropriate. Percentages are calculated using available (non-missing) data for each variable.
\* Valid percentages exclude missing or unknown responses.
\*\* Percentages for unknown values are calculated using the full sample size (N = 30,999)
\*\*\*Long-term conditions and disabilities were self-reported and obtained from a separately collected dataset that was merged with the main sample; as a result, missing or unknown responses are not applicable for these variables.
PHQ-9 = Patient Health Questionnaire-9;
GAD-7 = Generalised Anxiety Disorder-7;
WSAS = Work and Social Adjustment Scale.

### Model development

Separate prediction models were developed for each of the seven binary outcomes. Continuous predictors were scaled and centred, while categorical predictors were dummy-coded. Predictors with at least 70% data completion across the sample were included. Missing predictor values were imputed using the K-nearest neighbours (KNN) imputer with K = 5 following established guidelines. KNN imputation is a non-parametric method that leverages similarity between observations to estimate missing values without assuming an underlying distribution, making it suitable for heterogeneous clinical data with mixed variable types [31]. We specifically used FastKNNImputer, which is optimised for speed allowing rapid imputation across large samples without compromising accuracy. The source code is available at https://github.com/Zethson/fknni.

We fitted an elastic net regression, which balances bias and variance, reduces overfitting, and ensures interpretability [32]. Hyperparameters were tuned using grid search with 5-fold cross-validation. The elastic net mixing parameter α was evaluated over the values 0.1, 0.3, 0.5, 0.7, and 0.9. The regularisation strength parameter C, which controls the overall degree of penalisation applied to the model (with smaller values indicating stronger regularisation and greater coefficient shrinkage, and larger values indicating weaker regularisation), was evaluated over 0.01, 0.1, 1.0, and 10.0 [33].

To visualise predictor importance, we created heatmaps of the absolute coefficient values for the top 20 predictors for each outcome. Importance was based on the absolute coefficient size, indicating stronger influence regardless of direction. To show consistent patterns, we averaged predictor importance across depression outcomes and separately across anxiety outcomes. The total number of predictors in these averaged top 20 figures may vary because some predictors might not make the averaged list even if they were in the top 20 for an individual outcome. See **Supplementary Figures 3-5** for all predictors selected for depression, anxiety, and functional impairment.

### Model evaluation

Models were internally validated using nested bootstrap resampling and assessed in terms of overall performance (Brier score, area under the curve (AUC), sensitivity, specificity, positive predictive value (PPV), and negative predictive value (NPV)), and calibration (slope, intercept, and calibration plot). Sensitivity, specificity, PPV, and NPV were calculated using a predicted-probability classification threshold of 0.50.

Nested bootstrap was chosen because it provides an explicit correction for optimism, giving a more accurate estimate of model performance than conventional k-fold cross-validation or a holdout set [34]. All modelling steps described above (including pre-processing, missing data imputation, and hyperparameter tuning) were repeated at each bootstrap iteration to avoid data leakage. We used Harrell’s optimism-corrected bootstrap algorithm [35] by drawing 200 samples with replacement from the original dataset. At each iteration, the model was re-trained with re-tuned hyperparameters, and predictions were generated on the original dataset to assess out-of-sample performance.

We calculated optimism-corrected AUC by subtracting the average optimism (mean difference between apparent and test AUCs) from the apparent AUC. Its 95% confidence intervals were derived by subtracting the 2.5th and 97.5th percentiles of the optimism distribution from the apparent AUC [35, 36]. Sensitivity, specificity, PPV, NPV, and Brier score are presented as the median and 95% percentile intervals of bootstrap samples (see **Figures 6-8** in Supplementary Materials). Full validation details are in **Supplementary methods A**.

We assessed calibration using the intercept, slope, and calibration curves [37]. Calibration was assessed using logit-based calibration intercepts and slopes following standard procedures [38]. Optimism was adjusted for by recalculating metrics across 200 bootstrap iterations and applying a correction [35]. We applied nonparametric locally weighted scatterplot smoothing (LOWESS) to visualise the alignment between predicted probabilities and observed outcomes. We derived the optimism-corrected calibration curve by subtracting the average optimism observed across bootstrap iterations from the apparent LOWESS-smoothed calibration estimates. The optimism-corrected calibration intercept and slope were reported as summary measures, and curves were plotted for each outcome (see **Figures 6-8** in Supplementary Materials for details).

We assessed prediction stability, calibration stability, and Mean Absolute Prediction Error (MAPE) [15]. Prediction instability was assessed by comparing individual-level predictions across bootstrap models, with 95% stability intervals smoothed using LOWESS. Calibration instability was visualised by overlaying calibration curves from bootstrap models. Visualisations for calibration, prediction instability, calibration instability, and MAPE are provided in **Supplementary Figures 6-8**, for depression, anxiety, and functional impairment, respectively. Further methodological details are available in **Supplementary methods B**.

Data cleaning, model development, and visualisations were performed using Python version 3.12.12 [39]. The following packages were used: NumPy [40], pandas [41], scikit-learn [42], seaborn [43], statsmodels [44], and SciPy [45].

Code of the full model pipeline and performance evaluation using bootstrapping is available on GitHub (https://github.com/nourkanso/NHS-TT-Baseline-Prediction-Models).

### Sensitivity analyses

We developed alternative models using least absolute shrinkage and selection operator (LASSO), random forest, and gradient boosting for each of the seven outcomes **(Supplementary Tables 5-7).** We also performed a sensitivity analysis using elastic net to evaluate predictive performance after excluding data from March 1, 2020, to December 31, 2021, based on clinical advice to account for potential changes in treatment practices during the COVID-19 pandemic **(Supplementary Table 8).**

### TRIPOD+AI adherence

This study adheres to the Transparent Reporting of a Multivariable Prediction Model for Individual Prognosis or Diagnosis (TRIPOD+AI) guidelines to ensure methodological rigour and transparency in reporting [46] **(Supplementary Table 9).**

## Results

### Sample characteristics

The analytical sample included 30,999 high-intensity patients treated in SLaM NHS-TT services from January 1, 2018, to August 27, 2024. Cognitive behavioural therapy (48.9%) and counselling (46.1%) together accounted for approximately 95% of treatment episodes (**Supplementary Table 2)**.

**Table 1** describes the baseline sociodemographics of the sample. The sample predominantly consisted of women (73%) and had a median age of 34 years. Most patients identified as White ethnicity (57%), but there was also a notable representation of Black, Black British, Caribbean, or African ethnicities (22%).

**Table 2** presents the baseline clinical characteristics of the sample. The mean scores for depression (PHQ-9; [24]), anxiety (GAD-7; [25]]), and functional impairment (WSAS; [29, 30]) indicated moderate symptom severity and significant impairment, according to suggested thresholds. For detailed baseline and last-session PHQ-9, GAD-7, and WSAS measures by outcome (all vs. cases), see **Supplementary Table 10.** The sample also showed a range of primary diagnoses, with the most common being depressive episodes, followed by post-traumatic stress disorder and generalised anxiety disorder.

**Supplementary Table 11** summarises treatment outcomes for depression, anxiety, and functional impairment. In brief, for depression, reliable improvement was achieved by 47.8% of the sample, recovery by 55.7%, and reliable recovery by 46.9%. For anxiety, reliable improvement was seen in 57.9%, recovery in 52.9%, and reliable recovery in 49.2%. Functional impairment after treatment was observed in 58.2% of the sample.

### Model performance

Elastic net models showed robust performance across the outcomes, with good discrimination and calibration overall (see **Table 3**). For example, in predicting depression outcomes, the AUC for reliable improvement was 0.76, meaning the model correctly ranks those who improved higher than those who did not in 76% of cases. Sensitivity was high (0.76), indicating the model effectively identified individuals likely to improve, though specificity was moderate (0.63). In practical terms, of those flagged as likely to improve, 65% went on to do so (PPV), while 74% of those flagged as unlikely to improve did not (NPV).

**Table 3:**
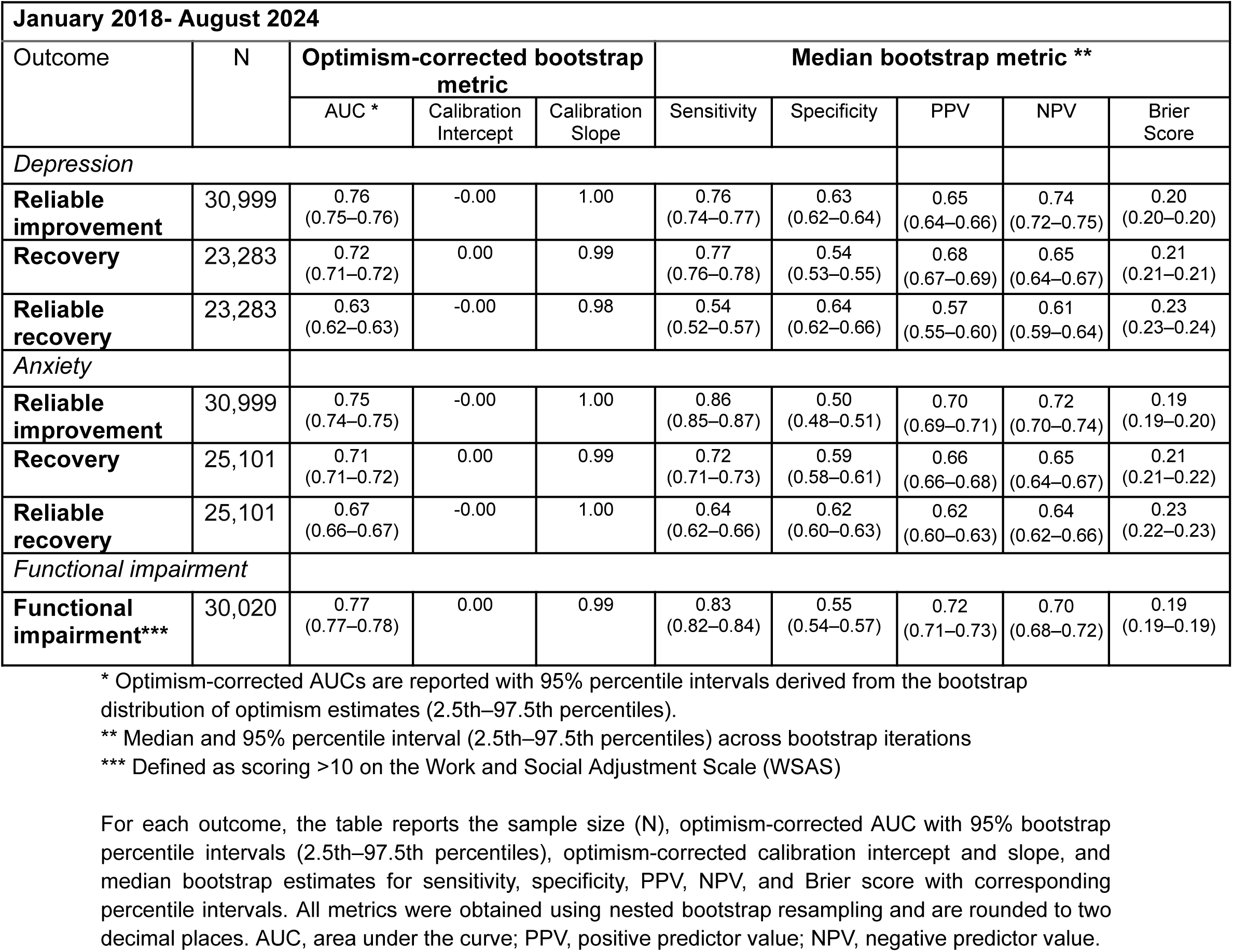
Internal validation performance of baseline prediction models for depression, anxiety, and functional impairment outcomes (January 2018–August 2024)

Functional impairment demonstrated the strongest predictive performance of the three outcomes, achieving a very good AUC (0.77), high sensitivity (0.84), and moderate specificity (0.55). This suggests the model reliably identifies individuals who are more likely to have functional impairment following treatment. PPV (0.72) and NPV (0.70) were both reasonably high, indicating the model performed well at both correctly flagging and correctly ruling out functional impairment.

Calibration measures indicated good alignment between predicted and observed outcomes, with slopes close to 1.00 for all outcomes.

Calibration curves, prediction instability, calibration instability, MAPE distribution plot, as well as distributions of optimism, AUC, sensitivity, specificity and Brier scores per bootstrap for each outcome (depression, anxiety, and functional impairment), can be found in **Supplementary Figures 6-8**, respectively. Overall, the plots demonstrated good stability for reliable improvement across depression and anxiety, with the best stability observed for functional impairment. Recovery outcomes showed slight deviations, while reliable recovery had the least stability among the outcomes.

LASSO models showed similar performance to elastic net but with slightly worse calibration, while random forest models had better discrimination across all outcomes but much poorer calibration (**Supplementary Tables 5-7**). Although gradient boost models offered slightly higher discrimination and comparable calibration to the elastic net, we ultimately chose the elastic net for its balance of reliable calibration, simplicity, and reduced risk of overfitting.

Sensitivity analyses excluding the time frame associated with the COVID-19 pandemic (March 2020 to December 2021) showed no difference in performance (**Supplementary Table 8**).

### Influential predictors

For all outcomes, functional impairment severity, mental health symptom severity, and sociodemographic factors played important roles in predicting outcomes following treatment.

As shown in **Figure 1**, the most important predictor across the depression outcomes was unemployment due to sickness or disability, followed by not being able to speak English, WSAS score, benefits recipient status, and medication status (not prescribed). Other predictors selected by the model included panic or phobia diagnosis, religion Muslim, other anxiety disorders diagnosis (which include somatoform disorders, hypochondriacal disorders), and reporting a long-term condition.

**Figure 1:**
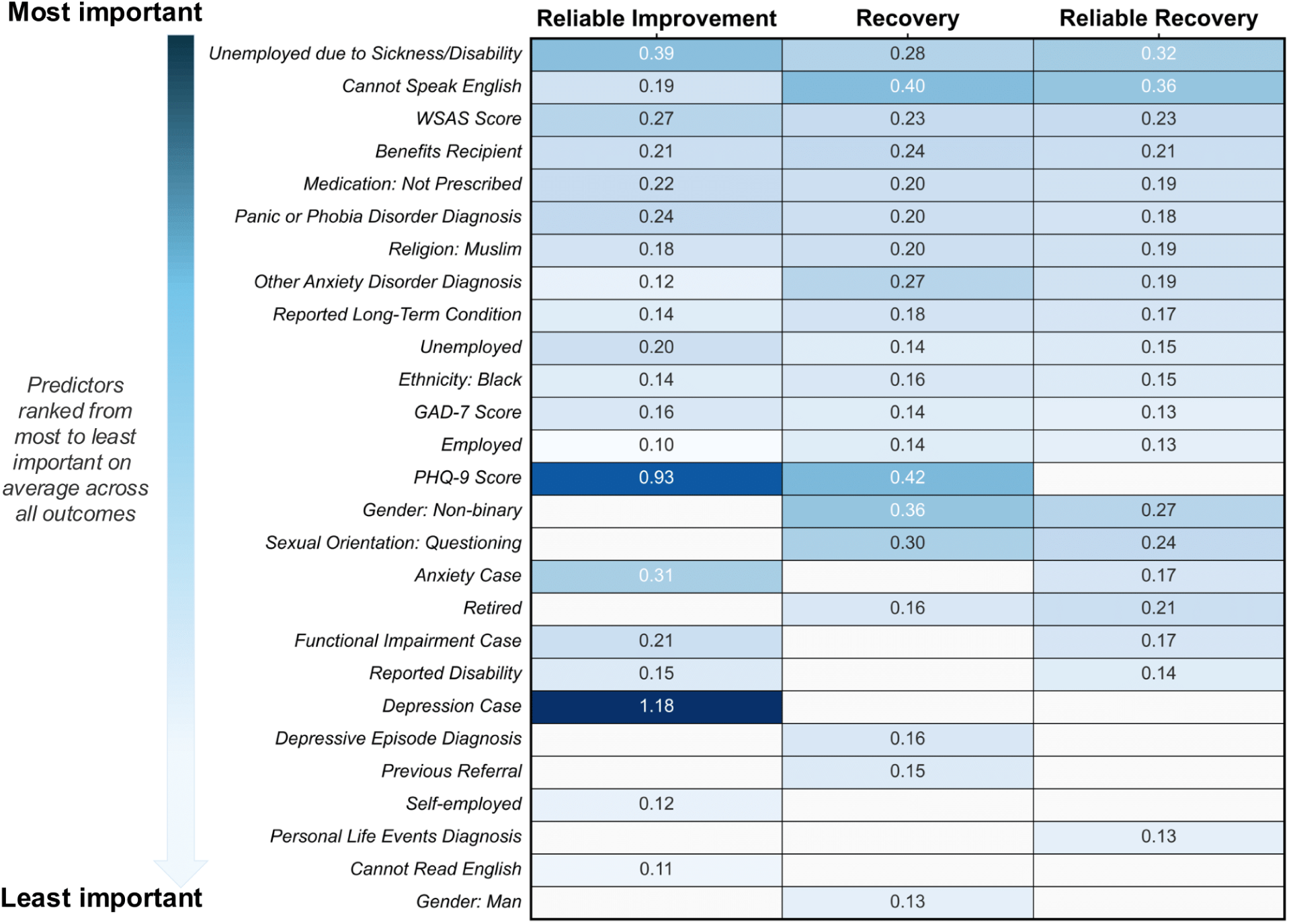
Top 20 predictors of depression (PHQ-9) outcomes (reliable improvement, recovery, and reliable recovery) and their absolute coefficients. Predictors are ranked based on their average importance across the three outcomes, showing only the top 20 features selected by the models. Beta coefficients are presented as absolute values, rounded to two decimal places, with no directionality indicated. For a detailed view of all predictors retained by depression models, see Supplementary Figure 3. WSAS, Work and Social Adjustment Scale; PHQ-9, Patient Health Questionnaire-9.

For anxiety (**Figure 2**), similar to depression outcomes, unemployment due to sickness or disability was the most important predictor, followed by PHQ-9 score. Other anxiety disorders (which include somatoform disorders, hypochondriacal disorders), being unemployed, and panic or phobia diagnosis were moderately associated with treatment outcomes. Other predictors included not being able to speak English, benefits recipient status, religion Muslim, questioning sexual orientation, and WSAS score.

**Figure 2:**
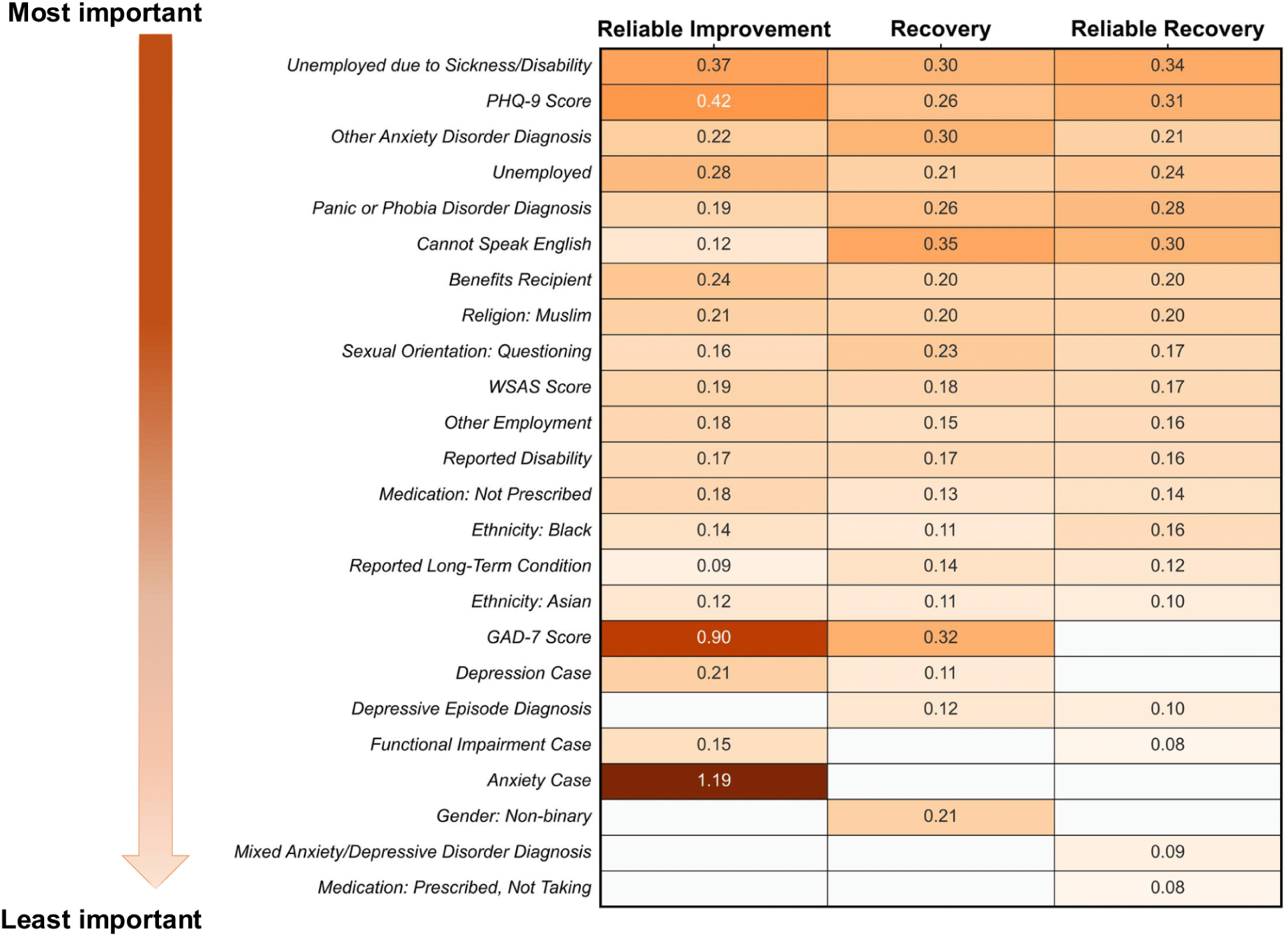
Top 20 predictors of anxiety (GAD-7) outcomes (reliable improvement, recovery, and reliable recovery) and their absolute coefficients. Predictors are ranked based on their average importance across the three outcomes, showing only the top 20 features selected by the models. Beta coefficients are presented as absolute values, rounded to two decimal places, with no directionality indicated. For a detailed view of all predictors retained by anxiety models, see Supplementary Figure 4. PHQ-9, Patient Health Questionnaire-9; GAD-7, Generalised Anxiety Disorder-7; WSAS, Work and Social Adjustment Scale.

For functional impairment (**Figure 3**), functional impairment “caseness” (i.e., being functionally impaired at baseline) was the strongest predictor, followed by WSAS score and other anxiety disorder diagnosis. Several sociodemographic and clinical factors also featured among the top predictors, including sexual orientation, gender, PHQ-9 score, and reporting a disability.

**Figure 3:**
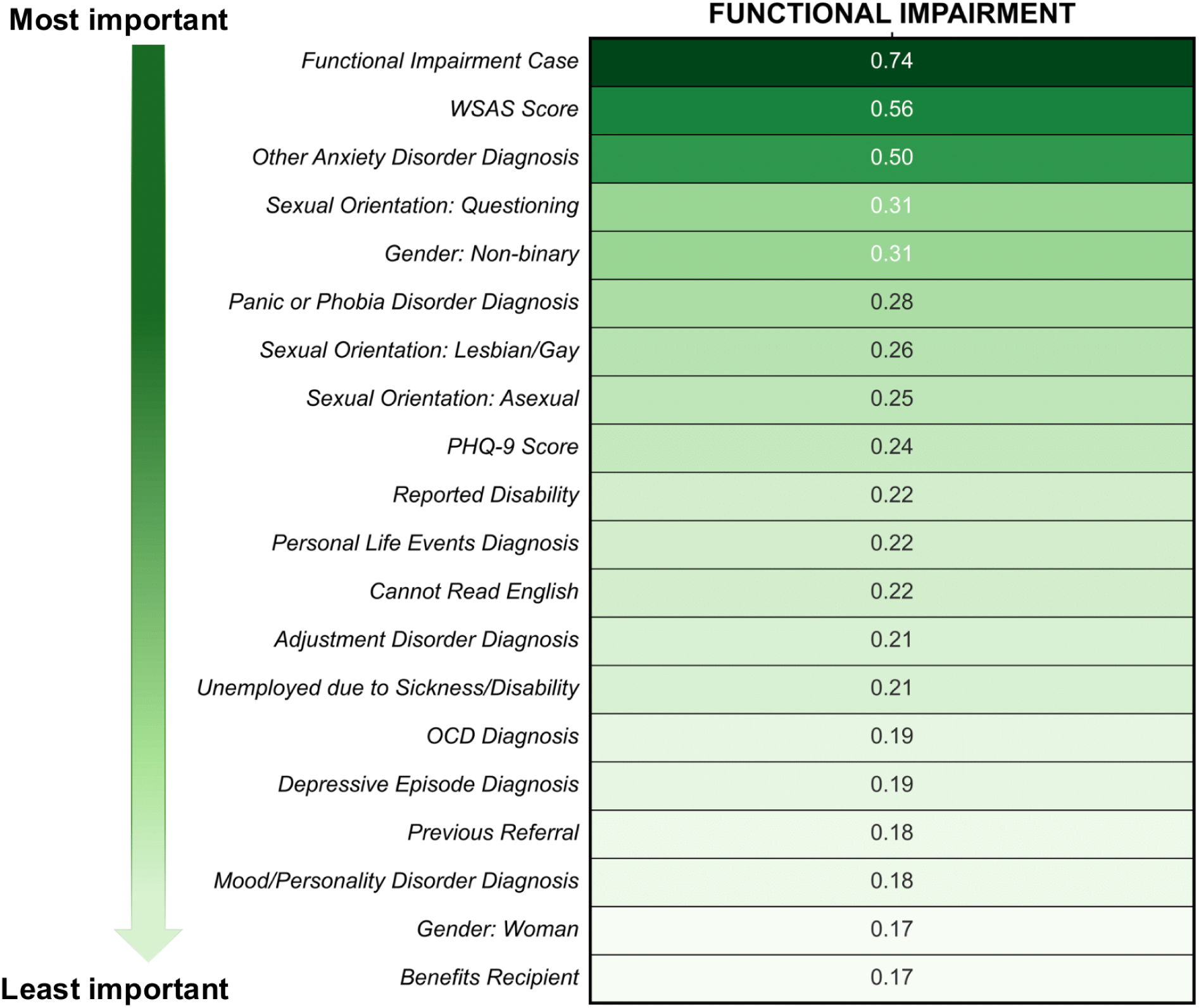
Top 20 predictors of the functional impairment (WSAS) outcome and their absolute coefficients. The graph shows only the top 20 features selected by the model. Beta coefficients are presented as absolute values, rounded to two decimal places, with no directionality indicated. For a detailed view of all predictors retained by the model, see Supplementary Figure 5. WSAS, Work and Social Adjustment Scale; PHQ-9, Patient Health Questionnaire-9; OCD, Obsessive-Compulsive Disorder.

## Discussion

Using routinely collected baseline data, we developed and internally validated models for depression, anxiety, and functional impairment outcomes within the NHS-TT programme. The models demonstrated moderate to good performance, with strong discrimination and robust calibration across outcomes. Consistent with clinical expectations, key predictors included symptom severity, primary diagnosis, presence of a disability or long-term condition, medication usage, and sociodemographic factors such as not being able to speak English, employment status, and receipt of benefits. Our findings provide empirical support that reinforces clinical judgement with evidence, enhancing confidence in using such models to support decision-making.

### Clinical applications and implications

The models’ outcomes, reliable improvement, recovery, and reliable recovery, are standard NHS outcomes reported annually and used to evaluate the service, often defined by pooling changes in PHQ-9/GAD-7 together [26]. We instead defined and built models for these outcomes separately, taking a domain-specific approach, to allow for individual-level prediction modelling. This is clinically and methodologically important, as prior work shows differential treatment trajectories and responses across PHQ-9 and GAD-7 measures within mixed diagnostic groups (e.g., patients may improve in anxiety but not depression, or vice versa). This approach is well supported in the literature on prediction and stratified care (e.g., [7, 47, 48]) and aligns with aims of personalised prediction.

Our prediction models aim to support stratified treatment for anxiety and depression. By accurately identifying, at assessment, patients at risk of poor outcomes, such models enable early intervention, prompting clinical concern, planning, and action to improve patient outcomes [49]. For example, there is evidence to suggest that prediction models may change clinician outcome expectations; therapists tend to be over-optimistic about patient prognosis [50], and patient monitoring could help therapists reduce this positive bias [51]. New information from models may also help therapists to make subtle changes to treatment, such as emphasising all treatment options with patients, and having open discussions on where, when, how, and by whom the therapy would be delivered [23]. Involving the patient in treatment selection would not only improve engagement (and consequently, outcomes), but research has shown that treatments that are more credible to patients are likely to be more effective [52]. At an organisational level, these models can inform resource allocation and inform new service designs by identifying patients benefiting from additional support, such as peer support groups or supplementary medication consultations.

### Summary of results and comparison with previous studies

Our models demonstrated good performance across all outcomes, particularly for reliable improvement and functional impairment, but were less predictive for reliable recovery outcomes, consistent with previous findings. Direct comparison with previous NHS-TT prediction work is complicated by differences in outcome definition and modelling approach. For example, Bone et al. [47] predicted reliable and clinically significant improvement—an outcome analogous to our reliable recovery—using a dynamic model that incorporated symptom scores collected session by session. Their model reached an AUC of approximately 0.80 once several sessions of data had accrued, but their baseline-only model did not exceed an AUC of 0.65. This baseline figure is the appropriate comparator for our approach, which uses only information available at assessment, and is consistent with our reliable recovery models (depression AUC = 0.63, anxiety AUC = 0.67). Discrimination for reliable recovery was consistently lower than for reliable improvement in both depression and anxiety, which likely reflects that reliable recovery combines two criteria (crossing a clinical threshold and exceeding the scale’s measurement error) making it inherently harder to predict, particularly for patients with baseline scores near the threshold or with moderate symptom reductions. Research elsewhere looking at predicting outcomes in community mental health services achieved an AUC of 0.76 for reliable change [53], showing similar performance to the reliable improvement (depression AUC = 0.76, anxiety AUC = 0.75) and functional impairment (AUC = 0.77) models.

Few previous NHS-TT models have considered calibration, despite its crucial role in clinical decision-making, where even small misalignments can have significant implications for patient care [15]. One study predicting depression treatment response reported poor discrimination (AUC = 0.57–0.61 dependent on model type) and poor calibration, indicated by calibration curves [54]. In contrast, our models demonstrated both good discrimination and robust calibration, reinforcing their potential clinical utility. While some metric thresholds have been proposed to be clinically significant for informing treatment decisions [55], determining exactly when they become clinically meaningful is complex. Clinical utility metrics, such as net benefit, are also crucial for assessing whether predictions translate into meaningful treatment decisions.

### Predictor importance

PHQ-9 symptom severity was an important predictor across depression and anxiety outcomes, supporting the conclusions of previous NHS-TT studies [7, 9, 47]. Baseline PHQ-9 scores appear to have limited importance for predicting reliable recovery, with a similar pattern observed for GAD-7 in anxiety outcomes. This may reflect that reliable recovery is defined not only by end-of-treatment scores but also by exceeding the measurement error threshold; patients with very high or very low baseline scores may achieve reliable recovery with smaller absolute changes, reducing the apparent predictive weight of baseline severity. Another possibility is that other baseline factors, such as functional impairment or sociodemographic characteristics, capture variance in treatment response more consistently once initial symptom severity is accounted for. Indeed, baseline WSAS score was an important predictor, suggesting that integrating functional measures into routine assessments could improve predictions and inform more tailored interventions, as demonstrated previously [10, 11].

Other relevant predictors included medication usage, and the presence of disabilities or long-term conditions, both of which remained important even after accounting for baseline symptom severity and functional impairment. These factors have been associated with worse outcomes in prior research [7, 9, 11]. Receiving benefits was also a strong predictor across all outcomes, a finding possibly unique to our sample as this data may not be routinely collected or might be excluded elsewhere due to multicollinearity. Although benefits status overlaps with factors like employment, disability, and deprivation, its independent contribution in our multivariable models suggests it captures distinct variance. Therefore, benefits data should be included in routine assessments and models to improve predictive accuracy and better address patient needs. The relevance of these predictors underscores the necessity for consistent data collection on these variables to inform treatment decisions effectively.

Employment status was also a key predictor for all outcomes. Previous research within NHS-TT has consistently found an association between unemployment and poorer outcomes [9, 11, 56, 57]. While our findings do not imply causality, they highlight the practical importance of employment status as a predictor. Employment status is amenable to intervention, and addressing it may improve patient symptoms and recovery: studies show that combining employment-related support with psychological therapy in NHS-TT settings improves outcomes [58]. Recognising this, since 2022, the Department for Work and Pensions has invested over £120 million to expand Employment Advisors in NHS-TT across England [59]. Over 90% of services now offer this support, leading to a 62% increase in individuals receiving employment advice compared to the previous year [60]. This initiative reflects a growing recognition of the links between employment and mental health and the benefits of addressing vocational issues to improve patient outcomes.

Other key predictors included ethnicity, religion, and ability to speak or read English. Previous work has highlighted that ethnic minority groups often experience worse trajectories during therapy [11] and are more likely to experience a reliable deterioration in symptoms [61]. It is essential to stress that such associations are not causal; rather, they serve as indicators of underlying structural barriers, such as racism, discrimination, reduced access to services, and potential cultural differences that may affect treatment [62]. For example, previous research within SLaM found that minority ethnic groups were less likely to receive a psychological assessment compared to the White British group, and among those who were assessed, they were also less likely to receive treatment [13]. More effort is needed to ensure mental health services are appropriately adapted for culturally diverse populations. Culturally tailored interventions have shown improvements in treatment outcomes for ethnic minority groups, indicating that addressing these factors may lead to better patient outcomes [63, 64]. A recent initiative by the NHS Race and Health Observatory has called on healthcare leaders to tackle these inequalities by improving resources, providing cultural competence training, and diversifying the workforce within the NHS-TT, which is currently predominantly white and female [65]. These predictors highlight the impact of structural and systemic factors that may influence treatment access and effectiveness.

Finally, gender and sexual orientation were also important predictors, particularly for functional impairment. Findings for small subgroups, like those questioning their sexual orientation or identifying as non-binary, require caution due to small sample size [66]. Previous research shows sexual and gender minority groups often have poorer treatment outcomes, such as higher baseline impairment and lower improvement odds for lesbian and bisexual women versus heterosexual women [67, 68]. Broader cohort data also show interactions between sexual orientation, gender, and recovery outcomes even after adjusting for clinical and sociodemographic factors [69]. Stigma, fear of disclosure, and limited practitioner understanding may drive these disparities, suggesting structural and social influences on therapy engagement [70]. NHS initiatives, including the Advancing Mental Health Equalities strategy and positive practice guidance for LGBTQ+ communities, aim to improve inclusivity and outcomes through training and culturally sensitive resources [71].

### Study strengths and limitations

Our study followed best practices for prediction modelling, using bootstrap-based bias correction to improve generalisability [35, 36]. Unlike previous NHS-TT models, which focused on AUC and either omitted or showed poor calibration [54], we reported both discrimination and calibration in line with TRIPOD+AI guidance. For all outcomes, calibration intercepts and slopes were close to ideal, supporting the reliability of our models for clinical decision-making.

Although our analytical sample is large, it comes from a single South London Trust (four boroughs) and does not fully represent the wider English population. According to the 2023 population estimates, 43% of the Central London population (which includes Southwark and Lambeth) identify as Black and Minority Ethnic (BME) and 44% are non-UK-born. In comparison, the rest of England only has 14% of the population identifying as BME and 13% being non-UK-born [72]. While not nationally representative, the sample’s high ethnic diversity is a strength, allowing assessment of model performance in an often underrepresented population.

Additionally, given the heterogeneity of low-intensity therapy, particularly in terms of the number of sessions received and delivery methods (e.g., individual to group settings), we decided to focus on high-intensity cases which represent more severe clinical presentations [21]. This limitation should be considered when interpreting the findings in the context of clinical practice.

A further consideration is outcome measurement. NHS-TT use the PHQ-9 for depression and the GAD-7 as the primary transdiagnostic anxiety measure, despite its original focus on generalised anxiety disorder. While validated as a reliable transdiagnostic screener for other anxiety disorders, such as panic disorder [73], the GAD-7 may not capture disorder-specific change as well as newer, disorder-specific measures as recommended by the NHS-TT manual [74]. However, the latter measures have high missingness in routine data, forcing a trade-off between the standardised, large sample size enabled by using PHQ-9/GAD-7, and the granularity of outcome measurement.

Our results are also based on internal validation, which quantifies optimism and overfitting within the same dataset. Nested bootstrap provides important insight into model reliability and generalisability, suggesting how informative models will likely remain for new patients, though it does not replace external validation in independent datasets [75]. To ensure the models are clinically useful and can be generalised beyond South London, retaining utility in other Trusts or regions, external validation using independent datasets is essential.

## Conclusion

The NHS-TT aims for equity, meaning clinical outcomes should not be affected by characteristics such as age, ethnicity, or gender [23]. Alarmingly, almost all of these are important predictors of outcomes in our models. This highlights not only the need to address inequities in care delivery, but also the importance of developing and validating models using diverse samples. Building accurate, reliable, and fair prediction models is a crucial step towards stratified care, but there is a need for research on how to best integrate prediction models into practice. Models’ accuracy and impact need careful, transparent external validation across England, and their clinical utility, net benefit, and impact on delivery within the existing NHS framework must also be evaluated [76]. Future work may include conducting prospective comparative studies, such as cluster-randomised trials [51], alongside qualitative research informed by patient and clinician perspectives of these models. Ultimately, the present study provides a strong foundation for initiatives to create a more equitable and effective healthcare system.

## Data Availability

The data used in this study are not publicly available because they are derived from anonymised electronic health records held by South London and Maudsley NHS Foundation Trust and accessed via the Clinical Record Interactive Search (CRIS) system under approved governance procedures (Project ID: 22-078). Access to these data is subject to approval by the South London and Maudsley NHS Foundation Trust CRIS Oversight Committee.
All code used for the analyses is publicly available at: https://github.com/nourkanso/NHS-TT-Baseline-Prediction-Models

## List of abbreviations

AUC: Area under the receiver operating characteristic curve
BME: Black and Minority Ethnic
CBT: Cognitive behavioural therapy
CRIS: Clinical Record Interactive Search
GAD-7: Generalised Anxiety Disorder-7
IAPT: Improving Access to Psychological Therapies
IMD: Index of Multiple Deprivation
KNN: K-nearest neighbours
LASSO: Least absolute shrinkage and selection operator
LOWESS: Locally weighted scatterplot smoothing
MAPE: Mean Absolute Prediction Error
NHS: National Health Service
NHS-TT: NHS Talking Therapies
NICE: National Institute for Health and Care Excellence
NPV: Negative predictive value
PHQ-9: Patient Health Questionnaire-9
PPV: Positive predictive value
SLaM: South London and Maudsley NHS Foundation Trust
SSP: Statutory Sick Pay
TRIPOD+AI: Transparent Reporting of a multivariable prediction model for Individual Prognosis Or Diagnosis + Artificial Intelligence
WSAS: Work and Social Adjustment Scale

## Declarations

### Ethics approval and consent to participate

This study used de-identified data from the Clinical Record Interactive Search (CRIS) system. CRIS operates within a robust governance framework and received ethical approval as an anonymised data resource for secondary analysis. The use of these data was approved by the CRIS Oversight Committee (Project 22-078).

### Consent for publication

Not applicable.

### Availability of data and materials

The data used in this study were obtained from the CRIS system at the South London and Maudsley NHS Foundation Trust and are not publicly available due to information governance restrictions protecting patient confidentiality. Access to CRIS data can be requested subject to approval from the CRIS Oversight Committee. The code for the full model pipeline and performance evaluation is available on GitHub at https://github.com/nourkanso/NHS-TT-Baseline-Prediction-Models.

## Acknowledgments

NK is funded by the King’s College London DRIVE-Health Centre for Doctoral Training. This study is part-funded by the National Institute for Health and Care Research (NIHR) Maudsley Biomedical Research Centre (BRC). The views expressed are those of the authors and not necessarily those of the NHS, NIHR or the Department of Health and Social Care. We would like to thank the CRIS-SLaM data extraction team for their support and data provision.

## References

1. Baker C. Mental health statistics: prevalence, services and funding in England. London: Commons Library Research Briefing; 2021.

2. McManus S, Bebbington P, Jenkins R, Brugha T. Mental health and wellbeing in England: Adult psychiatric morbidity survey 2014. 2016.

3. Tiller JWG. Depression and anxiety. Med J Aust. 2013;199. 10.5694/mja12.10628.

4. Michl LC, McLaughlin KA, Shepherd K, Nolen-Hoeksema S. Rumination as a mechanism linking stressful life events to symptoms of depression and anxiety: Longitudinal evidence in early adolescents and adults. J Abnorm Psychol. 2013;122:339–52. 10.1037/a0031994.

5. McHugh RK, Barlow DH. The dissemination and implementation of evidence-based psychological treatments: A review of current efforts. Am Psychol. 2010;65:73–84. 10.1037/a0018121.

6. Population Health, Clinical Audit and Specialist Care Team, NHS Digital. NHS Talking Therapies, for anxiety and depression, Annual reports, 2023-24. England: NHS; 2024.

7. Delgadillo J, Moreea O, Lutz W. Different people respond differently to therapy: A demonstration using patient profiling and risk stratification. Behav Res Ther. 2016;79:15–22. 10.1016/j.brat.2016.02.003.

8. Delgadillo J, De Jong K, Lucock M, Lutz W, Rubel J, Gilbody S, et al. Feedback-informed treatment versus usual psychological treatment for depression and anxiety: a multisite, open-label, cluster randomised controlled trial. Lancet Psychiatry. 2018;5:564–72. 10.1016/S2215-0366(18)30162-7.

9. Saunders R, Cape J, Fearon P, Pilling S. Predicting treatment outcome in psychological treatment services by identifying latent profiles of patients. J Affect Disord. 2016;197:107–15. 10.1016/j.jad.2016.03.011.

10. Saunders R, Buckman JEJ, Cape J, Fearon P, Leibowitz J, Pilling S. Trajectories of depression and anxiety symptom change during psychological therapy. J Affect Disord. 2019;249:327–35. 10.1016/j.jad.2019.02.043.

11. Skelton M, Carr E, Buckman JEJ, Davies MR, Goldsmith KA, Hirsch CR, et al. Trajectories of depression and anxiety symptom severity during psychological therapy for common mental health problems. Psychol Med. 2022;:1–11. 10.1017/S0033291722003403.

12. Arundell L-LC, Saunders R, Buckman JEJ, Lewis G, Stott J, Singh S, et al. Differences in psychological treatment outcomes by ethnicity and gender: an analysis of individual patient data. Soc Psychiatry Psychiatr Epidemiol. 2024;59:1519–31. 10.1007/s00127-024-02610-8.

13. Harwood H, Rhead R, Chui Z, Bakolis I, Connor L, Gazard B, et al. Variations by ethnicity in referral and treatment pathways for IAPT service users in South London. Psychol Med. 2023;53:1084–95. 10.1017/S0033291721002518.

14. Delamain H, Buckman JEJ, O’Driscoll C, Suh JW, Stott J, Singh S, et al. Predicting post-treatment symptom severity for adults receiving psychological therapy in routine care for generalised anxiety disorder: a machine learning approach. Psychiatry Res. 2024;336:115910. 10.1016/j.psychres.2024.115910.

15. Riley RD, Collins GS. Stability of clinical prediction models developed using statistical or machine learning methods. Biom J. 2023;65:2200302. 10.1002/bimj.202200302.

16. Geisser S. The Predictive Sample Reuse Method with Applications. J Am Stat Assoc. 1975;70:320–8. 10.1080/01621459.1975.10479865.

17. Collins GS, Dhiman P, Ma J, Schlussel MM, Archer L, Van Calster B, et al. Evaluation of clinical prediction models (part 1): from development to external validation. BMJ. 2024;:e074819. 10.1136/bmj-2023-074819.

18. Clark DM. Realizing the Mass Public Benefit of Evidence-Based Psychological Therapies: The IAPT Program. Annu Rev Clin Psychol. 2018;14:159–83. 10.1146/annurev-clinpsy-050817-084833.

19. Stewart R, Soremekun M, Perera G, Broadbent M, Callard F, Denis M, et al. The South London and Maudsley NHS Foundation Trust Biomedical Research Centre (SLAM BRC) case register: development and descriptive data. BMC Psychiatry. 2009;9:51. 10.1186/1471-244X-9-51.

20. Saunders R, Cape J, Leibowitz J, Aguirre E, Jena R, Cirkovic M, et al. Improvement in IAPT outcomes over time: are they driven by changes in clinical practice? Cogn Behav Ther. 2020;13:e16. 10.1017/S1754470X20000173.

21. Bennett-Levy J, Richards D, Farrand P, Christensen H, Griffiths K, Kavanagh D, et al., editors. Oxford Guide to Low Intensity CBT Interventions. Oxford University Press; 2015. 10.1093/med:psych/9780199590117.001.0001.

22. NICE. Common mental health problems: identification and pathways to care. London; 2011.

23. The National Collaborating Centre for Mental Health. NHS Talking Therapies for anxiety and depression Manual. 2024.

24. Kroenke K, Spitzer RL, Williams JBW. The PHQ-9: Validity of a brief depression severity measure. J Gen Intern Med. 2001;16:606–13. 10.1046/j.1525-1497.2001.016009606.x.

25. Spitzer RL, Kroenke K, Williams JBW, Löwe B. A Brief Measure for Assessing Generalized Anxiety Disorder: The GAD-7. Arch Intern Med. 2006;166:1092. 10.1001/archinte.166.10.1092.

26. National Collaborating Centre for Mental Health. The Improving Access to Psychological Therapies Manual. Guidance. England: NHS; 2023.

27. Jacobson NS, Truax P. Clinical significance: A statistical approach to defining meaningful change in psychotherapy research. J Consult Clin Psychol. 1991;59:12–9. 10.1037/0022-006X.59.1.12.

28. Fried EI, Nesse RM. The Impact of Individual Depressive Symptoms on Impairment of Psychosocial Functioning. PLoS ONE. 2014;9:e90311. 10.1371/journal.pone.0090311.

29. Marks IM. Behavioural psychotherapy: Maudsley pocket book of clinical management. Bristol: Wright; 1986.

30. Mundt JC, Marks IM, Shear MK, Greist JM. The Work and Social Adjustment Scale: a simple measure of impairment in functioning. Br J Psychiatry. 2002;180:461–4. 10.1192/bjp.180.5.461.

31. Zhang S. Nearest neighbor selection for iteratively kNN imputation. J Syst Softw. 2012;85:2541–52. 10.1016/j.jss.2012.05.073.

32. Zou H, Hastie T. Regularization and Variable Selection via the Elastic Net. J R Stat Soc Ser B Stat Methodol. 2005;67:301–20.

33. Rockova V, Lesaffre E, Luime J, Löwenberg B. Hierarchical Bayesian formulations for selecting variables in regression models. Stat Med. 2012;31:1221–37. 10.1002/sim.4439.

34. Wilimitis D, Walsh CG. Practical Considerations and Applied Examples of Cross-Validation for Model Development and Evaluation in Health Care: Tutorial. JMIR AI. 2023;2:e49023. 10.2196/49023.

35. Harrell FE, Lee KL, Mark DB. MULTIVARIABLE PROGNOSTIC MODELS: ISSUES IN DEVELOPING MODELS, EVALUATING ASSUMPTIONS AND ADEQUACY, AND MEASURING AND REDUCING ERRORS. Stat Med. 1996;15:361–87. 10.1002/(SICI)1097-0258(19960229)15:4<361::AID-SIM168>3.0.CO;2-4.

36. Noma H, Shinozaki T, Iba K, Teramukai S, Furukawa TA. Confidence intervals of prediction accuracy measures for multivariable prediction models based on the bootstrap-based optimism correction methods. Stat Med. 2021;40:5691–701. 10.1002/sim.9148.

37. Austin PC, Steyerberg EW. Graphical assessment of internal and external calibration of logistic regression models by using loess smoothers. Stat Med. 2014;33:517–35. 10.1002/sim.5941.

38. Steyerberg EW. Clinical Prediction Models: A Practical Approach to Development, Validation, and Updating. Cham: Springer International Publishing; 2019. 10.1007/978-3-030-16399-0.

39. Python Software Foundation. Python 3.12.12. 2025.

40. Harris CR, Millman KJ, Van Der Walt SJ, Gommers R, Virtanen P, Cournapeau D, et al. Array programming with NumPy. Nature. 2020;585:357–62. 10.1038/s41586-020-2649-2.

41. The pandas development team. pandas-dev/pandas: Pandas. 2024. 10.5281/ZENODO.3509134.

42. Buitinck L, Louppe G, Blondel M, Pedregosa F, Mueller A, Grisel O, et al. API design for machine learning software: experiences from the scikit-learn project. 2013. 10.48550/ARXIV.1309.0238.

43. Waskom M. seaborn: statistical data visualization. J Open Source Softw. 2021;6:3021. 10.21105/joss.03021.

44. Seabold S, Perktold J. Statsmodels: Econometric and Statistical Modeling with Python. Austin, Texas; 2010. p. 92–6. 10.25080/Majora-92bf1922-011.

45. Virtanen P, Gommers R, Oliphant TE, Haberland M, Reddy T, Cournapeau D, et al. SciPy 1.0: fundamental algorithms for scientific computing in Python. Nat Methods. 2020;17:261–72. 10.1038/s41592-019-0686-2.

46. Collins GS, Moons KGM, Dhiman P, Riley RD, Beam AL, Van Calster B, et al. TRIPOD+AI statement: updated guidance for reporting clinical prediction models that use regression or machine learning methods. BMJ. 2024;385:e078378. 10.1136/bmj-2023-078378.

47. Bone C, Simmonds-Buckley M, Thwaites R, Sandford D, Merzhvynska M, Rubel J, et al. Dynamic prediction of psychological treatment outcomes: development and validation of a prediction model using routinely collected symptom data. Lancet Digit Health. 2021;3:e231–40. 10.1016/S2589-7500(21)00018-2.

48. Delgadillo J, McMillan D, Lucock M, Leach C, Ali S, Gilbody S. Early changes, attrition, and dose–response in low intensity psychological interventions. Br J Clin Psychol. 2014;53:114–30. 10.1111/bjc.12031.

49. McAleavey AA, De Jong K, Nissen-Lie HA, Boswell JF, Moltu C, Lutz W. Routine Outcome Monitoring and Clinical Feedback in Psychotherapy: Recent Advances and Future Directions. Adm Policy Ment Health Ment Health Serv Res. 2024;51:291–305. 10.1007/s10488-024-01351-9.

50. Kaiser T, Herzog P, Voderholzer U, Brakemeier E-L. Out of sight, out of mind? High discrepancy between observer- and patient-reported outcome after routine inpatient treatment for depression. J Affect Disord. 2022;300:322–5. 10.1016/j.jad.2022.01.019.

51. Hisler GC, Young KS, Cumpanasoiu DC, Palacios JE, Duffy D, Enrique A, et al. Incorporating a deep-learning client outcome prediction tool as feedback in supported internet-delivered cognitive behavioural therapy for depression and anxiety: A randomised controlled trial within routine clinical practice. Couns Psychother Res. 2025;25:e12771. 10.1002/capr.12771.

52. Kumpasoğlu GB, Campbell C, Saunders R, Fonagy P. Therapist and treatment credibility in treatment outcomes: A systematic review and meta-analysis of clients’ perceptions in individual and face-to-face psychotherapies. Psychother Res. 2025;35:139–54. 10.1080/10503307.2023.2298000.

53. Chai KEK, Graham-Schmidt K, Lee CMY, Rock D, Coleman M, Betts KS, et al. Predicting anxiety treatment outcome in community mental health services using linked health administrative data. Sci Rep. 2024;14:20559. 10.1038/s41598-024-71557-2.

54. Coley RY, Boggs JM, Beck A, Simon GE. Predicting outcomes of psychotherapy for depression with electronic health record data. J Affect Disord Rep. 2021;6:100198. 10.1016/j.jadr.2021.100198.

55. Forsell E, Jernelöv S, Blom K, Kaldo V. Clinically sufficient classification accuracy and key predictors of treatment failure in a randomized controlled trial of Internet-delivered Cognitive Behavior Therapy for Insomnia. Internet Interv. 2022;29:100554. 10.1016/j.invent.2022.100554.

56. Buckman JEJ, Stott J, Main N, Antonie DM, Singh S, Naqvi SA, et al. Understanding the psychological therapy treatment outcomes for young adults who are not in education, employment, or training (NEET), moderators of outcomes, and what might be done to improve them. Psychol Med. 2023;53:2808–19. 10.1017/S0033291721004773.

57. Delgadillo J, Asaria M, Ali S, Gilbody S. On poverty, politics and psychology: the socioeconomic gradient of mental healthcare utilisation and outcomes. Br J Psychiatry. 2016;209:429–30. 10.1192/bjp.bp.115.171017.

58. Thew GR, Popa A, Allsop C, Crozier E, Landsberg J, Sadler S. The addition of employment support alongside psychological therapy enhances the chance of recovery for clients most at risk of poor clinical outcomes. Behav Cogn Psychother. 2024;52:93–9. 10.1017/S1352465823000474.

59. Department for Work and Pensions. £122 million employment boost for people receiving mental health support. 2022.

60. NHS England. NHS supports thousands more people back into work. 2025.

61. Arundell L-LC, Saunders R, Buckman JEJ, Lewis G, Stott J, Singh S, et al. Differences in psychological treatment outcomes by ethnicity and gender: an analysis of individual patient data. Soc Psychiatry Psychiatr Epidemiol. 2024;59:1519–31. 10.1007/s00127-024-02610-8.

62. Ladin K, Cuddeback J, Duru OK, Goel S, Harvey W, Park JG, et al. Guidance for unbiased predictive information for healthcare decision-making and equity (GUIDE): considerations when race may be a prognostic factor. Npj Digit Med. 2024;7:290. 10.1038/s41746-024-01245-y.

63. Arundell L-L, Barnett P, Buckman JEJ, Saunders R, Pilling S. The effectiveness of adapted psychological interventions for people from ethnic minority groups: A systematic review and conceptual typology. Clin Psychol Rev. 2021;88:102063. 10.1016/j.cpr.2021.102063.

64. Lie DA, Lee-Rey E, Gomez A, Bereknyei S, Braddock CH. Does Cultural Competency Training of Health Professionals Improve Patient Outcomes? A Systematic Review and Proposed Algorithm for Future Research. J Gen Intern Med. 2011;26:317–25. 10.1007/s11606-010-1529-0.

65. Mahase E. NHS must hire more diverse talking therapists to reduce inequalities, says report. BMJ. 2023;:p2537. 10.1136/bmj.p2537.

66. Harrell, FE. Regression Modeling Strategies: With Applications to Linear Models, Logistic and Ordinal Regression, and Survival Analysis. Cham: Springer International Publishing; 2015. 10.1007/978-3-319-19425-7.

67. Rimes KA, Broadbent M, Holden R, Rahman Q, Hambrook D, Hatch SL, et al. Comparison of Treatment Outcomes Between Lesbian, Gay, Bisexual and Heterosexual Individuals Receiving a Primary Care Psychological Intervention. Behav Cogn Psychother. 2018;46:332–49. 10.1017/S1352465817000583.

68. Rimes KA, Ion D, Wingrove J, Carter B. Sexual orientation differences in psychological treatment outcomes for depression and anxiety: National cohort study. J Consult Clin Psychol. 2019;87:577–89. 10.1037/ccp0000416.

69. Kent T, Suh JW, Lewis G, Saunders R, Davies NM, Lewis G, et al. Psychological therapy outcomes by sexual orientation and gender: a retrospective cohort study. Psychol Med. 2025;55:e212. 10.1017/S0033291725101220.

70. Morris DDA, Fernandes V, Rimes KA. Sexual minority service user perspectives on mental health treatment barriers to care and service improvements. Int Rev Psychiatry. 2022;34:230–9. 10.1080/09540261.2022.2051445.

71. Beattie S, Laville A. NHS Talking Therapies for Anxiety and Depression: LGBTQ+ Positive Practice Guide. 2024. https://babcp.com/wp-content/uploads/2025/06/LGBTQ-Positive-Practice-Guide.pdf.

72. Trust for London. London’s geography and population. 2024.

73. Kroenke K, Spitzer RL, Williams JBW, Monahan PO, Löwe B. Anxiety Disorders in Primary Care: Prevalence, Impairment, Comorbidity, and Detection. Ann Intern Med. 2007;146:317–25. 10.7326/0003-4819-146-5-200703060-00004.

74. NHS Digital. NHS Talking Therapies for Anxiety and Depression: Annual Reports, 2024–25. England: NHS Digital; 2025.

75. Iba K, Shinozaki T, Maruo K, Noma H. Re-evaluation of the comparative effectiveness of bootstrap-based optimism correction methods in the development of multivariable clinical prediction models. BMC Med Res Methodol. 2021;21:9. 10.1186/s12874-020-01201-w.

76. Jung K, Kashyap S, Avati A, Harman S, Shaw H, Li R, et al. A framework for making predictive models useful in practice. J Am Med Inform Assoc. 2021;28:1149–58. 10.1093/jamia/ocaa318.

