## Supplementary material for "Building a prediction model for outcomes following treatment in UK NHS Talking Therapies services for depression and anxiety"

Figure 1: Flowchart of data cleaning procedure. The sample includes all high-intensity patients who had between 3 and 21 sessions between the beginning of 2018–mid-2024 (N= 30,999) 2
Table 1: Proportion and cumulative proportion (%) of the total number of sessions per patient 4
Figure 2: Distribution of the total number of sessions per patient (N=30,999) 5
Table 2: Descriptions of sociodemographic variables 6
Table 3: Descriptions of clinical variables 8
Figure 3: All predictors and absolute coefficients for depression outcomes 11
Figure 4: All predictors and absolute coefficients for anxiety outcomes 13
Figure 5: All predictors and absolute coefficients for functional impairment outcome 15
Figure 6: Bootstrap results for depression outcomes 16
Figure 7: Bootstrap results for anxiety outcomes 34
Figure 8: Bootstrap results for functional impairment outcome 52
Table 4: Prediction performance metrics for LASSO models of depression, anxiety, and functional impairment outcomes 58
Table 5: Prediction performance metrics for random forest models of depression, anxiety, and functional impairment outcomes 59
Table 6: Prediction performance metrics for gradient boost models of depression, anxiety, and functional impairment outcomes 60
Table 7: Prediction performance metrics for depression, anxiety, and functional impairment outcomes, excluding the COVID-19 period, developed using elastic net logistic regression (sensitivity analysis) 61
Table 8: TRIPOD-AI checklist 62
Table 9: Descriptive statistics for PHQ-9, GAD-7, and WSAS scores at baseline and last session for a) all patients and b) depression or anxiety cases 64
Table 10: Treatment outcomes of depression, anxiety, and functional impairment 65
**Supplementary Methods 66**
A) Bootstrap Validation Procedure 66
B) Calibration and Stability Analyses 67

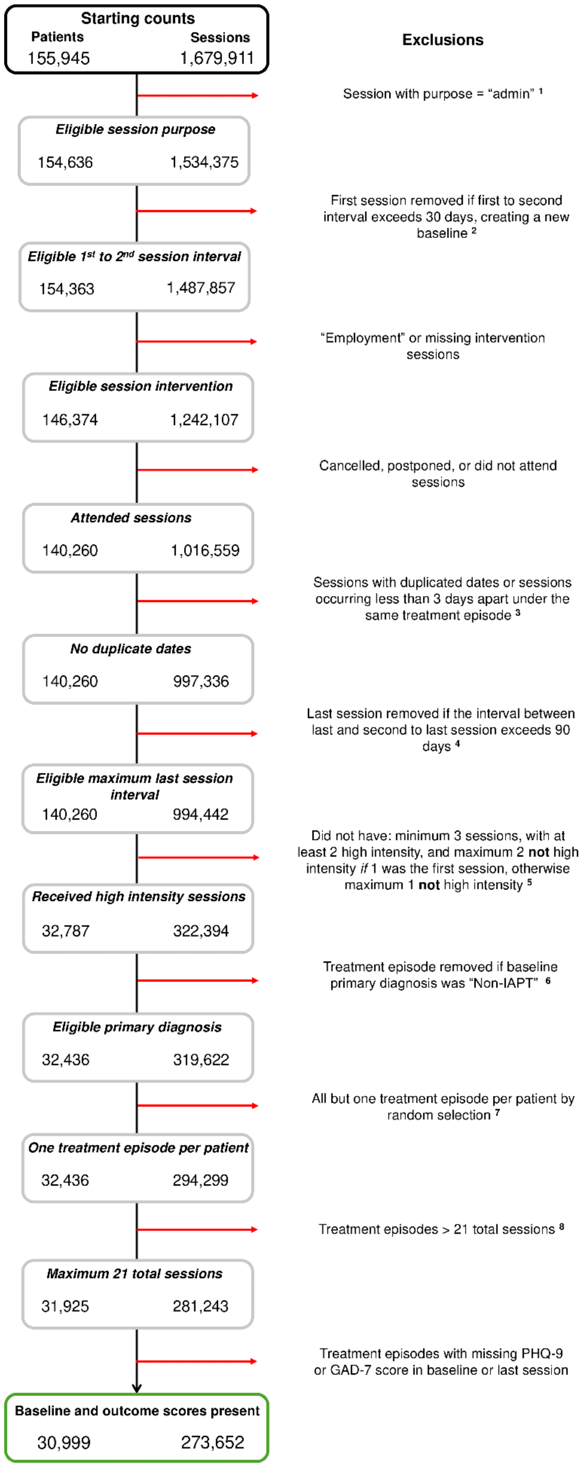


**Figure 1: Flowchart of data cleaning procedure. The sample includes all high-intensity patients who had between 3 and 21 sessions between the beginning of 2018–mid-2024 (N= 30,999)**

[1] Records of administrative sessions: focused on non-clinical tasks and often lacked measures of PHQ-9 and GAD-7.

[2] The initial session is generally an assessment or triage session, with treatment starting at the second session. If the gap between the first and second sessions exceeded 30 days, the second session was used as the baseline. This process was not repeated, as therapy with prolonged intervals between the 'new' baseline and the second session was deemed too disrupted for stable conclusions.

[3] Such sessions usually result from coding or data entry errors and were cleaned as follows:

- Removed if missing PHQ-9 and/or GAD-7 scores.
- If scores are identical, retain the most complete session; otherwise, keep the earliest session.
- If scores were different, calculate the mean score, and retain the most complete session; otherwise, keep the earliest session.

[4] A long interval between the last and second-to-last session may mean a miscoded new treatment episode or a follow-up. For this analysis, we removed such sessions based on clinical guidance, as we aimed to capture the outcome at the end of treatment rather than at follow-up.

[5] We focused the analysis on high-intensity interventions due to the differences in the average number of sessions and the level of structure between intervention intensities. However, because of the stepped-care format, most patients receive at least one low-intensity session, and the assessment/triage session may sometimes be classified as low-intensity when it is not coded as “non-intervention”.

[6] We removed treatment episodes where the baseline session’s primary diagnosis was “Non-IAPT” (often indicating employment-related topics not suitable for this dataset), which resulted in the exclusion of some patients.

[7] We kept only one treatment episode per patient to avoid bias and data leakage, ensuring no patient appeared in both the training and testing datasets. Since over 20% of patients were referred more than once initially, this approach was performed at this stage to maintain sample size and avoid randomly selecting episodes that did not eventually meet high-intensity criteria.

[8] The analysis was limited to patients with a maximum total number of 20 treatment sessions (i.e. 21 total sessions, using the first session as the baseline), aligning with the typical maximum duration of a treatment course. The initial skewed distribution may not have accurately reflected the typical treatment process and could distort the analysis; patients with an exceptionally high number of treatment sessions may have started another episode of care under the same treatment ID and were miscoded.

PHQ-9, Patient Health Questionnaire-9; GAD-7, Generalised Anxiety Disorder-7.

**Table 1: Proportion and cumulative proportion (%) of the total number of sessions per patient**

| Total no. of sessions | No. of patients | Proportion (%) | Cumulative proportion (%) |
|---|---|---|---|
| **3** | 1969 | 6.35 | 6.35 |
| **4** | 2152 | 6.94 | 13.29 |
| **5** | 2892 | 9.33 | 22.62 |
| **6** | 4744 | 15.30 | 37.93 |
| **7** | 3974 | 12.82 | 50.75 |
| **8** | 2118 | 6.83 | 57.58 |
| **9** | 1831 | 5.91 | 63.49 |
| **10** | 1457 | 4.70 | 68.19 |
| **11** | 1363 | 4.40 | 72.58 |
| **12** | 1576 | 5.08 | 77.67 |
| **13** | 1762 | 5.68 | 83.35 |
| **14** | 1361 | 4.39 | 87.74 |
| **15** | 1012 | 3.26 | 91.01 |
| **16** | 820 | 2.65 | 93.65 |
| **17** | 630 | 2.03 | 95.68 |
| **18** | 534 | 1.72 | 97.41 |
| **19** | 359 | 1.16 | 98.56 |
| **20** | 251 | 0.81 | 99.37 |
| **21** | 194 | 0.63 | 100.00 |

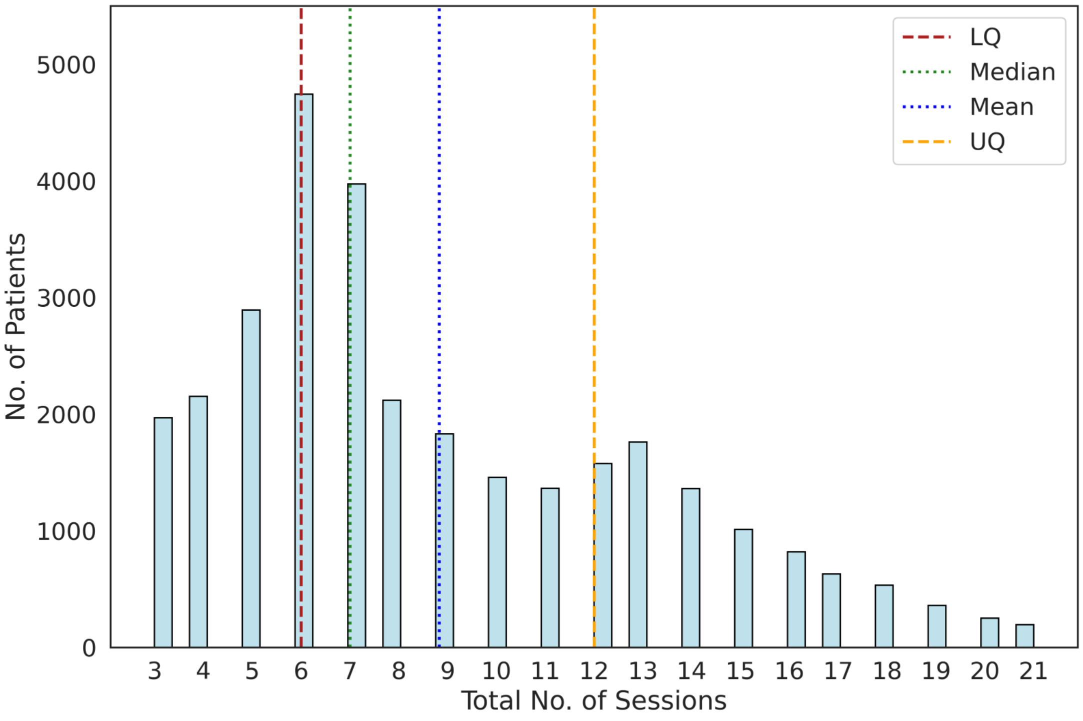


**Figure 2: Distribution of the total number of sessions per patient (N=30,999)**

Mean (SD): 8.83 (4.28), median (IQR): 7.0 (6-12), range: 3-21.

**Table 2: Specific psychological interventions received (N = 30,999)**

| Intervention | N | % |
|---|---|---|
| **Cognitive behavioural therapy (CBT)** | **15,144** | **48.85** |
| *Individual CBT* | *13,978* | *45.09* |
| *Group CBT* | *1,146* | *3.70* |
| *Computerised / guided self-help CBT* | *20* | *0.06* |
| **Counselling** | **14,295** | **46.11** |
| *Counselling (unspecified)* | *11,769* | *37.97* |
| *Counselling for Depression* | *1,711* | *5.52* |
| *Integrative counselling* | *694* | *2.24* |
| *Humanistic / person-centred* | *121* | *0.39* |
| **Psychodynamic or interpersonal therapy** | **553** | **1.78** |
| *Psychodynamic therapy* | *302* | *0.97* |
| *Dynamic interpersonal therapy* | *145* | *0.47* |
| *Interpersonal therapy* | *106* | *0.34* |
| **Eye movement desensitisation and reprocessing** | **513** | **1.65** |
| **Couples therapy** | **270** | **0.87** |
| **Other*** | **167** | **0.65** |

*Other includes interventions that did not map to a named modality, including mindfulness, residual/unspecified intervention codes and a small number psychoeducational entries.

**Table 3: Descriptions of sociodemographic variables**

| Variable | Description/ data collection | Categories/ coverage | Notes regarding data cleaning |
|---|---|---|---|
| **Age** | Collected as a continuous variable during baseline session. | Mean (SD), Median (IQR), Range | NA |
| **IMD Decile (2015)** | Index of Multiple Deprivation (IMD) decile based on patient postcode. | Deciles 1-10 (1 = most deprived, 10 = least deprived) | NA |
| **Ethnicity** | Self-reported ethnicity during baseline session. | White, Black/Black British/Caribbean/African, Mixed/Multiple, Asian/Asian British, Other | Followed the 2021 Census (https://www.ethnicity-facts-figures.service.gov.uk/style-guide/ethnic-groups/) |
| **Gender** | Self-reported gender identity during baseline session. | Woman (cisgender + transgender), Man (cisgender + transgender), Non-binary | There were inconsistencies in how data were collected. The dataset included multiple overlapping categories, such as: Female, Female (including transgender), Transgender female.<br><br>To address this, we consolidated these categories into broader, more inclusive groups: Woman: combines "female," "female (including transgender)," and "transgender female."<br><br>Similar steps were followed for men. |
| **Sexual Orientation** | Self-reported sexual orientation during baseline session. | Straight, Bisexual, Lesbian/Gay, Questioning, Queer, Asexual | There was some inconsistent reporting, e.g. overlapping categories such as: Lesbian, Gay, Lesbian or Gay.<br><br>So, we combined similar categories: Lesbian/Gay: combines "lesbian," "gay," and "lesbian or gay." |

| | | | |
|---|---|---|---|
| | | | Due to small numbers, pansexual was combined with bisexual. |
| **Employment status** | Self-reported employment status during baseline session. | Employed, Unemployed, Unemployed due to sickness/disability, Student, Self-employed, Retired, Other | “Other” includes smaller categories such as carers and homemakers. |
| **Religion** | Self-reported religious affiliation during baseline session. | Atheist/Agnostic, Christian, Muslim, Other | "Other" includes smaller self-reported religions, such as but not limited to Buddhist, Jewish, Sikh, and Jainist. |
| **Receive benefits** | Self-reported receipt of government benefits during baseline session. | Yes, No | NA |
| **Statutory Sick Pay (SSP)** | Self-reported receipt of SSP during baseline session. | Yes, No | NA |
| **Speak English** | Self-reported ability to speak English during baseline session. | Yes, No | NA |
| **Read English** | Self-reported ability to read English during baseline session. | Yes, No | NA |

**Table 4: Descriptions of clinical variables**

| Variable | Description/ data collection | Categories/ coverage | Notes regarding data cleaning |
|---|---|---|---|
| **PHQ-9** | Measures depression severity, collected during baseline session. Scale: Patient Health Questionnaire-9 | Mean (SD), Median (IQR), Range | NA |
| **GAD-7** | Measures anxiety severity, collected during baseline session. Scale: General Anxiety Disorder-7 | Mean (SD), Median (IQR), Range | NA |
| **WSAS** | Measures functional impairment, collected during baseline session. Scale: Work and Social Adjustment Scale | Mean (SD), Median (IQR), Range | NA |
| **Social Phobia** | Single-item measure of avoidance associated with social situations, recorded during the baseline assessment where completed. This item forms part of the NHS Talking Therapies minimum dataset. | Mean (SD), Median (IQR), Range | NA |
| **Agoraphobia** | Single-item measure of avoidance associated with agoraphobic situations, recorded during the baseline assessment where completed. This item forms part of the NHS Talking Therapies minimum dataset. | Mean (SD), Median (IQR), Range | NA |
| **Specific Phobia** | Single-item measure of avoidance associated with a personally identified feared object or situation, recorded during the baseline assessment where completed. This item forms part of the NHS Talking Therapies minimum dataset. | Mean (SD), Median (IQR), Range | NA |

| | | | |
|---|---|---|---|
| **Treatment service** | Indicates the service location where treatment was provided within SLaM. | Lambeth, Lewisham, Southwark, Croydon | NA |
| **Number of previous referrals** | This variable was derived by checking if the patient's ID appeared in any records prior to their baseline session date (including before 2018). It captures all previous referrals within the SLaM NHS-TT service. | Mean (SD), Median (IQR), Range | NA |
| **Primary diagnosis** | Captures the primary mental health diagnosis as input by the therapist or clinician during baseline session. | Depressive episode, Post-traumatic stress disorder, Generalised anxiety disorder, Panic/phobia, Reaction to severe stress and adjustment disorders, Obsessive-compulsive disorder, Mixed anxiety and depressive disorder, Other anxiety disorder, Other mood/personality disorder, Personal life events | Detailed table included below ***** |
| **Medication** | Captures baseline medication usage, including adherence. | Not prescribed, prescribed and taking, prescribed but not taking | NA |

| | | | |
|---|---|---|---|
| **>1 Long-term condition** | Indicates the presence of one or more long-term conditions, measured either at the baseline session or collected prior to | Yes, No | Multiple long-term conditions could have been reported. Data were mapped from a separate table to the baseline session to determine if the patient reported having a long-term condition on or before this date. Due to complications and unreliability in the data, specific conditions were not used as predictors; only the presence of any long-term condition was included. |
| **>1 Disability** | Indicates the presence of one or more disability, measured either at the baseline session or collected prior to | Yes, No | Multiple disabilities could have been reported. Data were mapped from a separate table to the baseline session to determine if the patient reported having a disability on or before this date. Due to complications and unreliability in the data, specific disabilities were not used as predictors; only the presence of any disability was included. |

#### * Primary diagnosis categories

| **Primary diagnosis** | **Notes** |
|---|---|
| Depressive episode | Includes depressive disorder (e.g., mild, moderate, recurrent, and severe episodes without psychotic symptoms) |
| Post-traumatic stress disorder | Includes PTSD |
| Generalised anxiety disorder | Includes GAD |
| Panic/phobia | Includes any panic disorder, social phobia, specific phobia, and agoraphobia |

| | |
|---|---|
| Reaction to severe stress and adjustment disorders | Consists of any adjustment disorders and acute stress reactions |
| Obsessive-compulsive disorder | Includes OCD |
| Mixed anxiety and depressive disorder | Co-occurrence of anxiety and depressive symptoms |
| Other anxiety disorders | Includes health anxiety, somatoform disorders, hypochondriacal disorder, hair-pulling/skin-picking, and unspecified anxiety disorders |
| Other mood/personality disorder | Includes bipolar affective disorder, schizophrenia, sleep disorders, body dysmorphic disorder, persistent mood disorders, and emotionally unstable personality disorder |
| Personal life events | Includes alcohol-related issues, drug use, family disruption (e.g., separation, divorce), death of family members, childhood neglect, and exposure to traumatic events like war or terrorism |

This table outlines the primary diagnostic categories used and the specific conditions included within each category.

### Figure 3: All predictors and absolute coefficients for depression outcomes

#### a) Reliable improvement

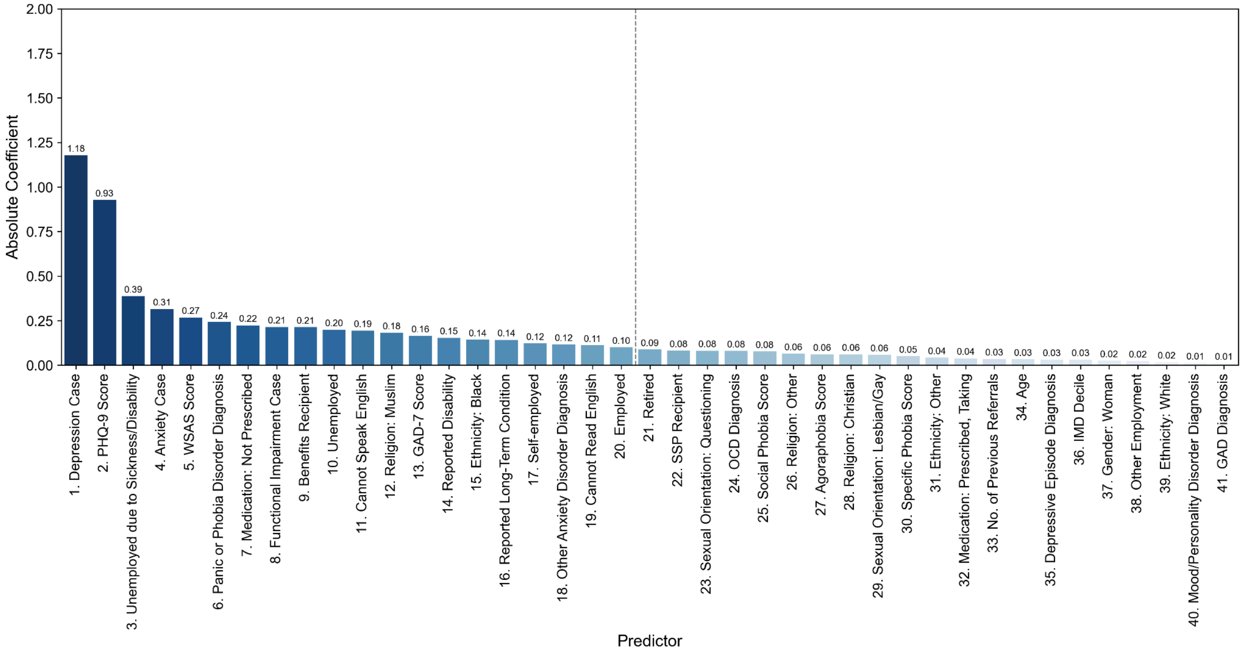


The dashed line after the first 20 predictors indicates the selected top predictors for this outcome, as highlighted in the heatmap presented in the main text.

#### b) Recovery

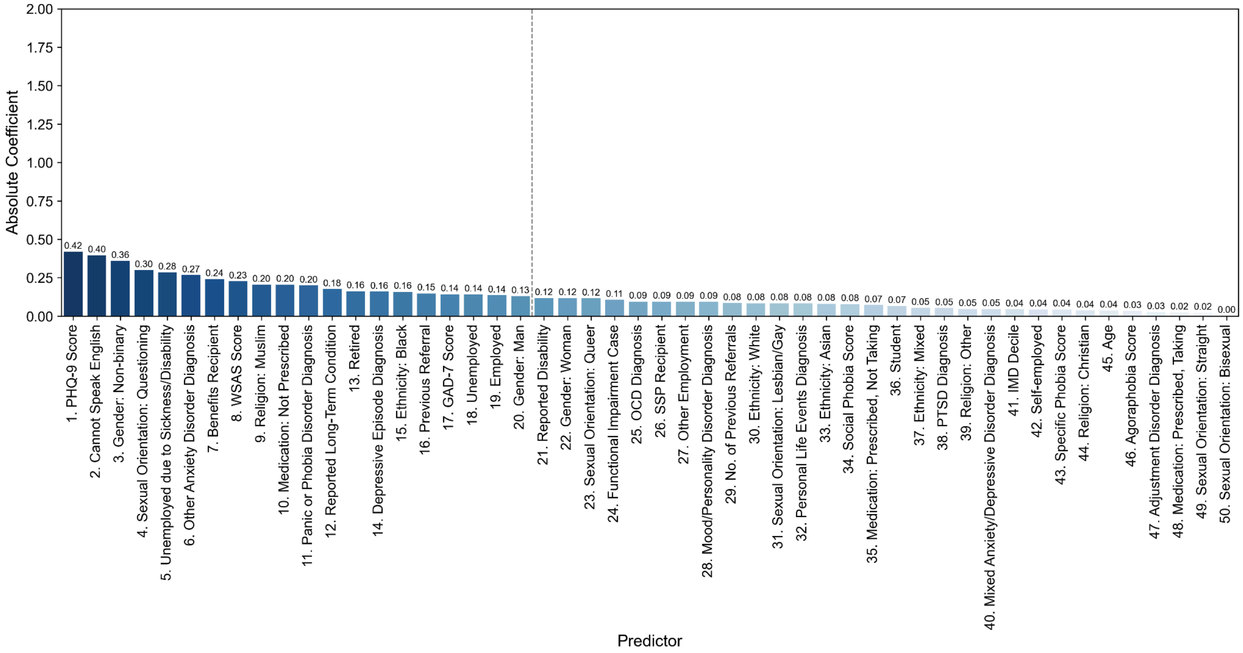

### c) Reliable recovery

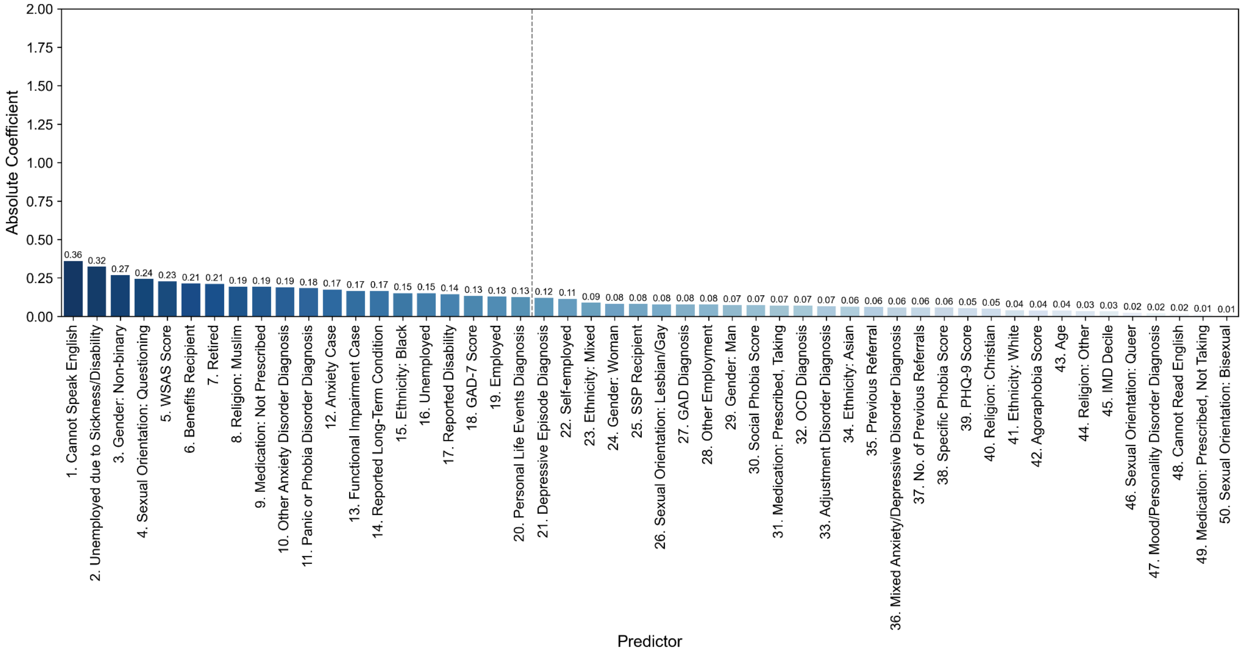

#### Figure 4: All predictors and absolute coefficients for anxiety outcomes

##### a) Reliable improvement

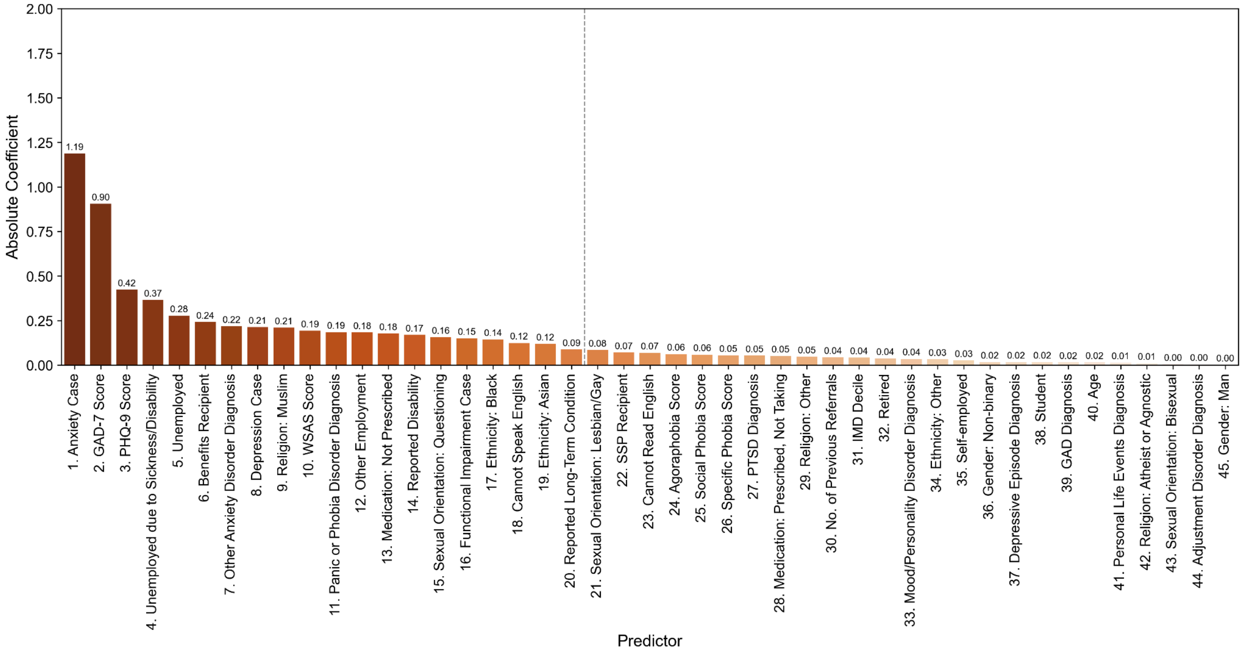


##### b) Recovery

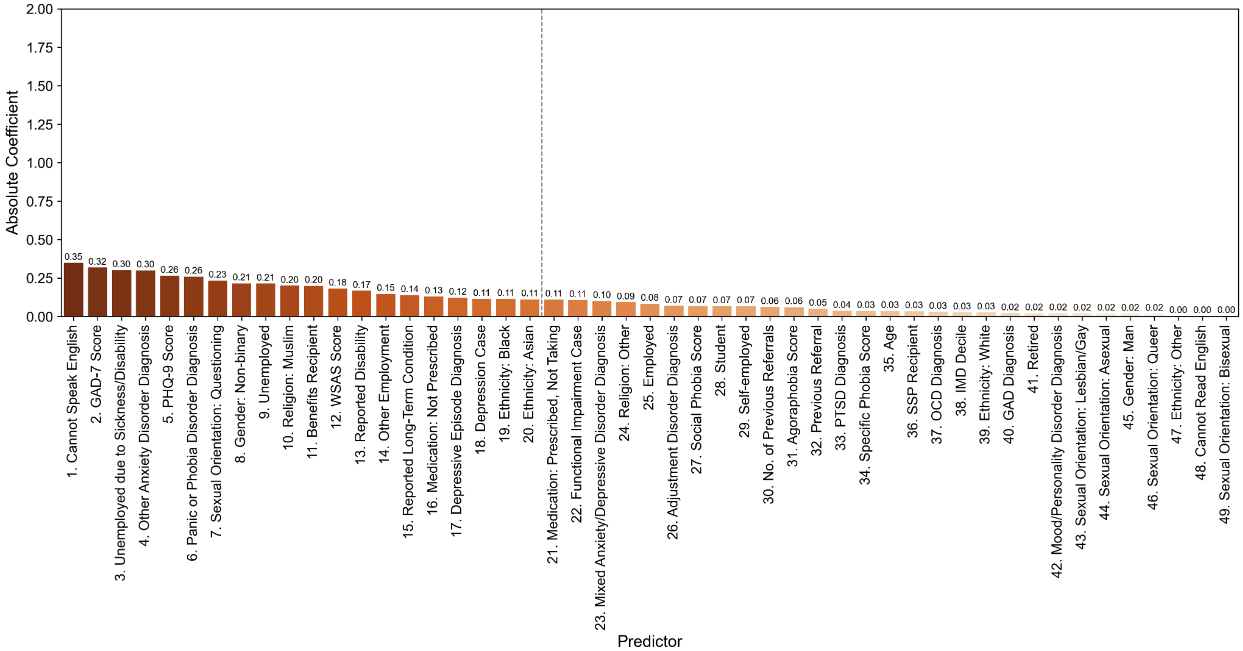

### c) Reliable recovery

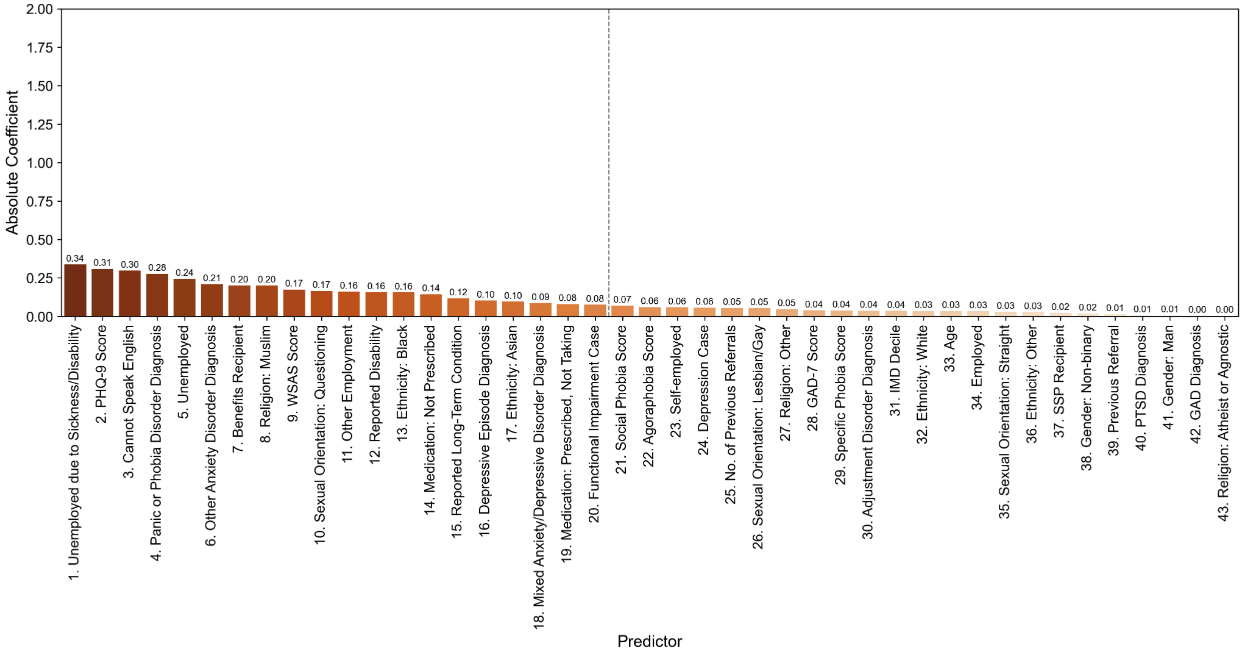

### Figure 5: All predictors and absolute coefficients for functional impairment outcome

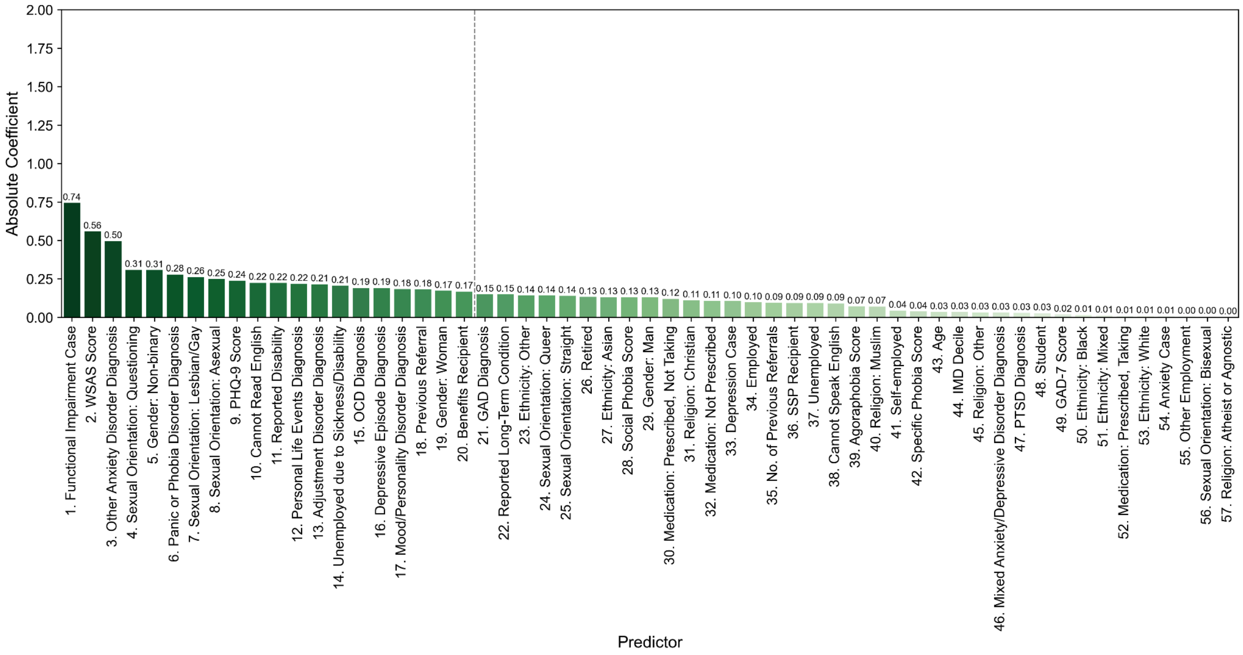

**Figure 6: Bootstrap results for depression outcomes**

**a) Reliable improvement**

- Calibration curve

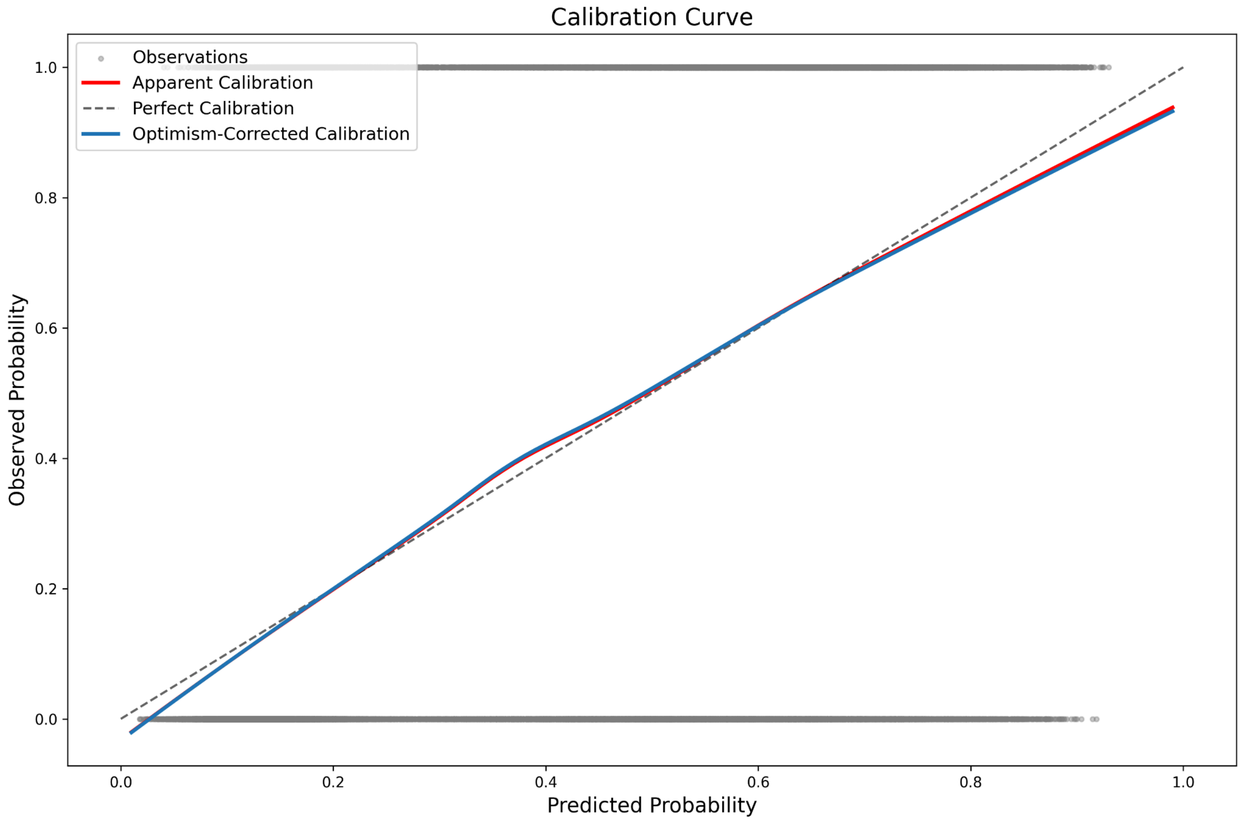

- Prediction instability

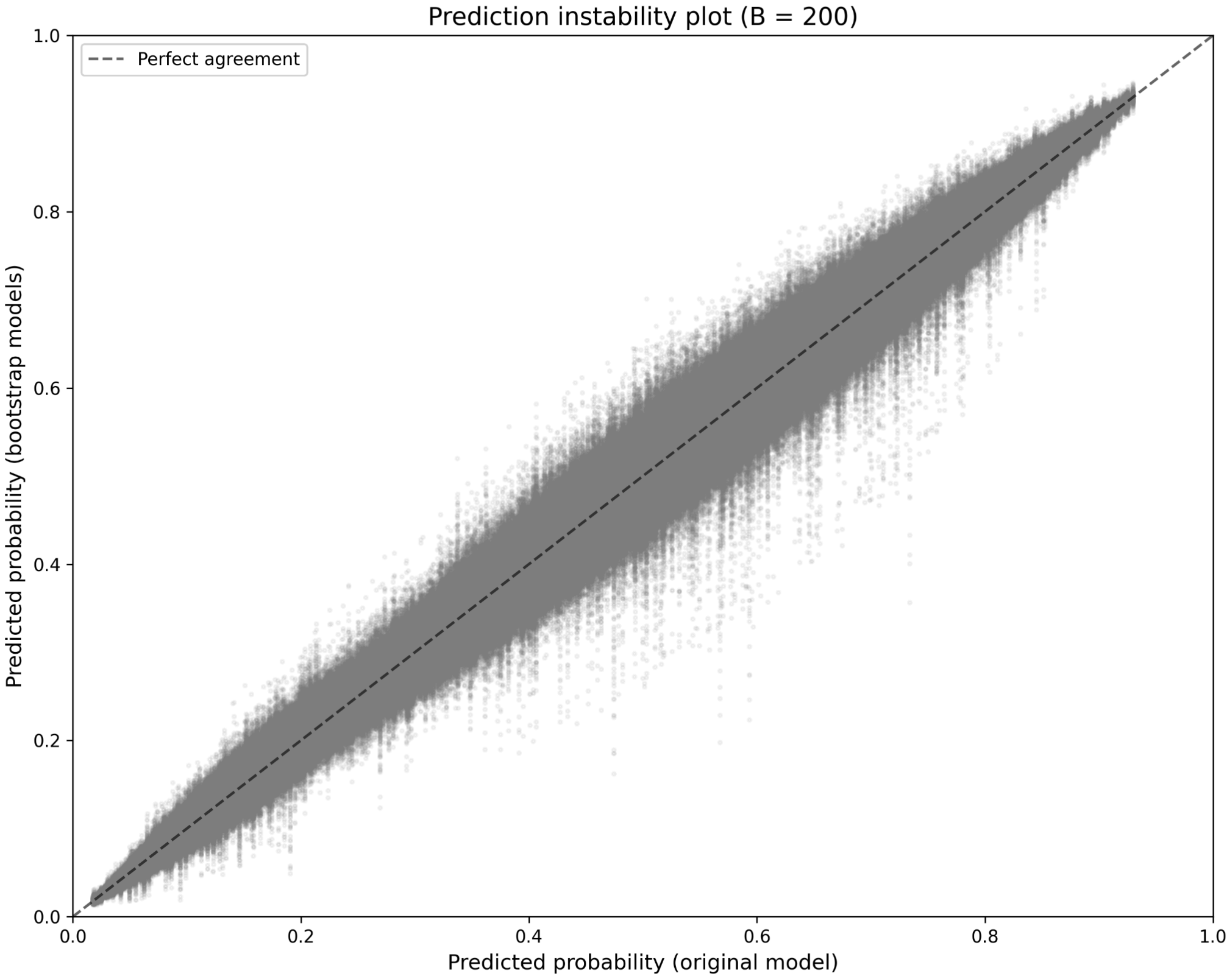

- Calibration instability

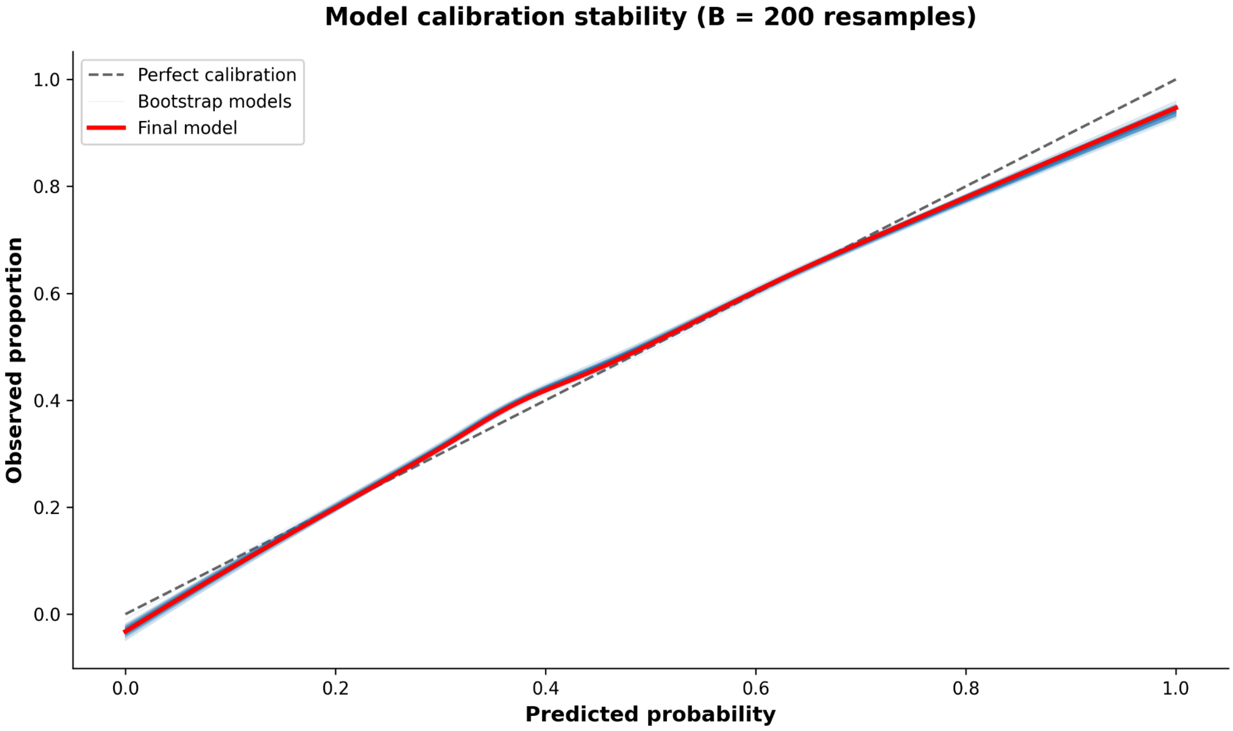

- MAPE

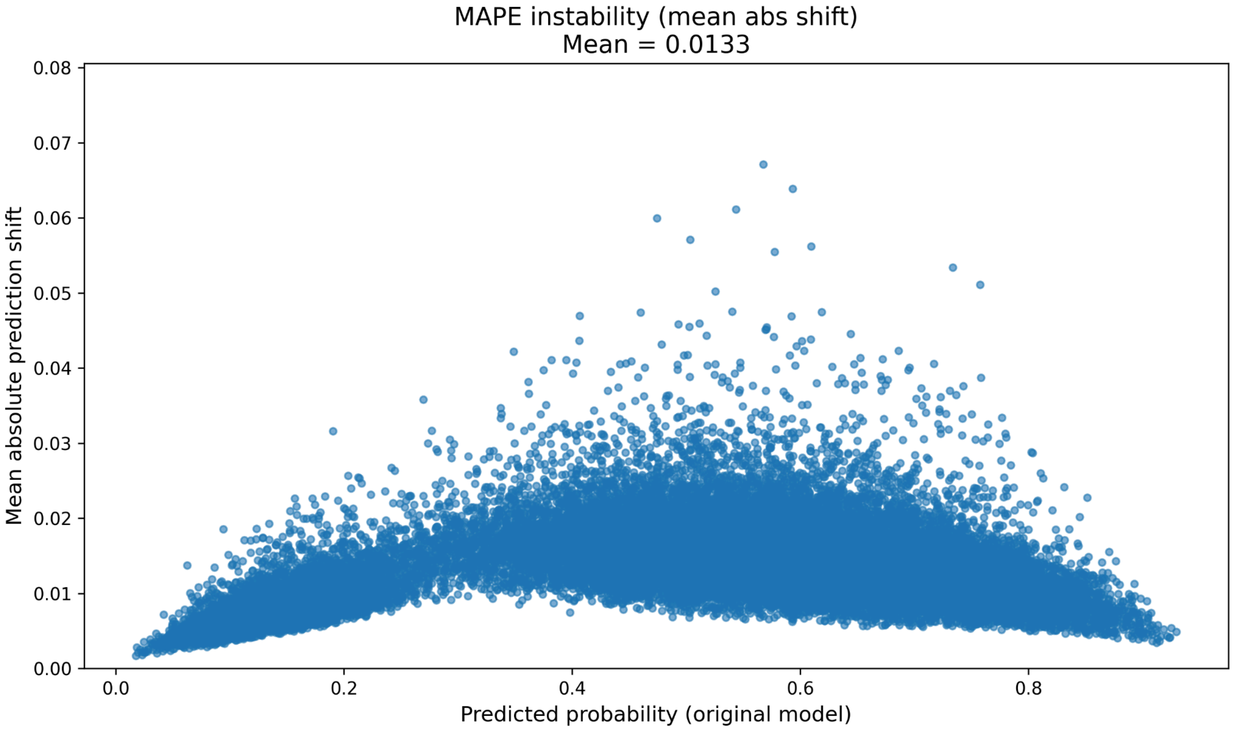


- Optimism

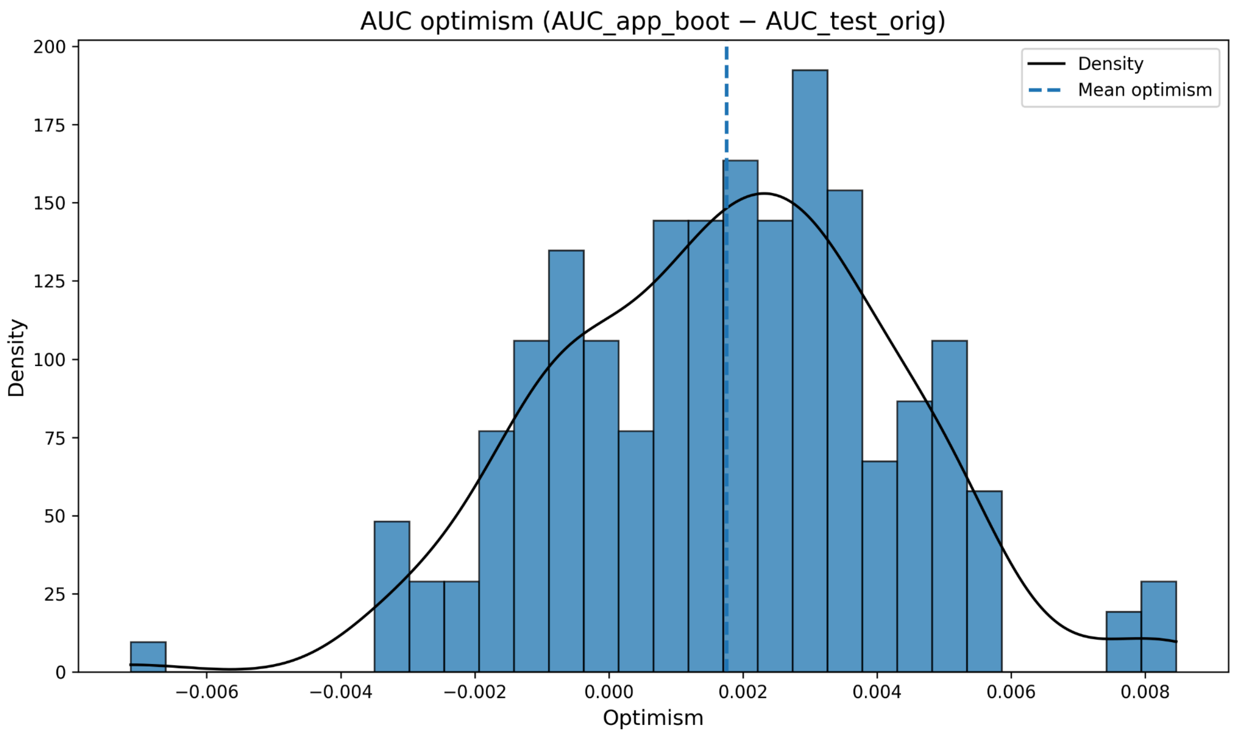


- AUC

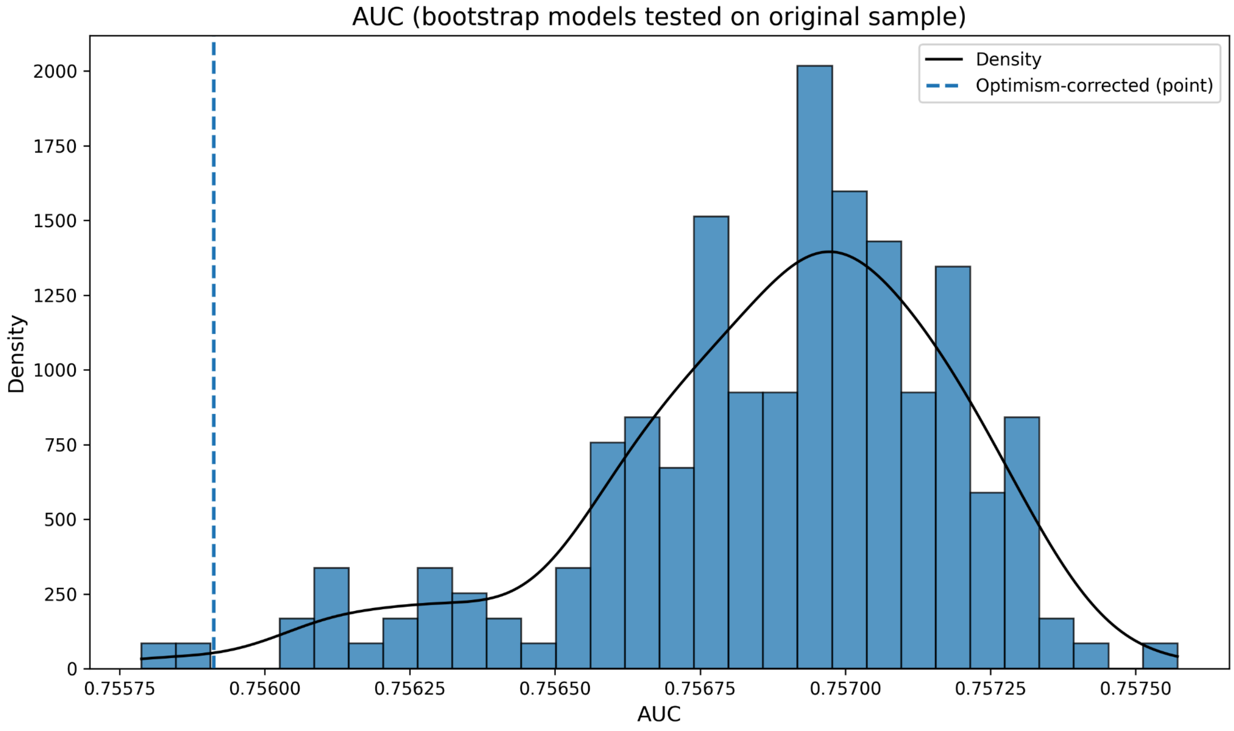


- Sensitivity

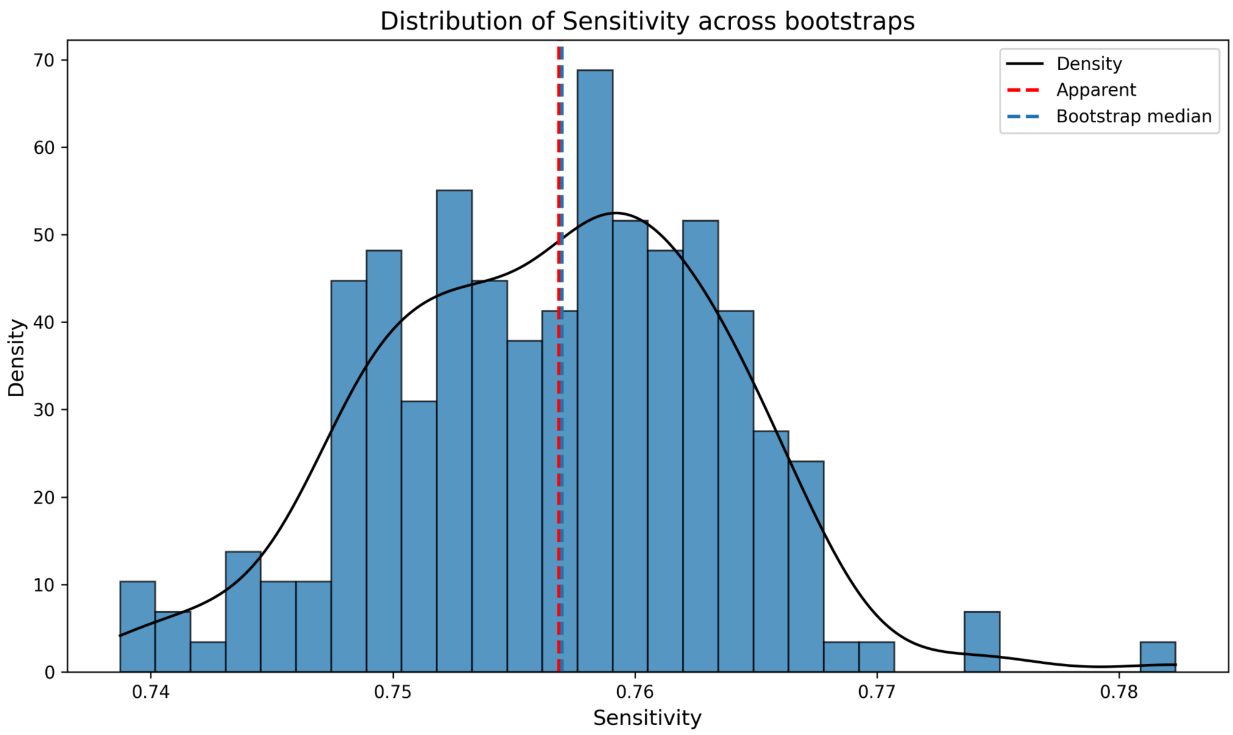


- Specificity

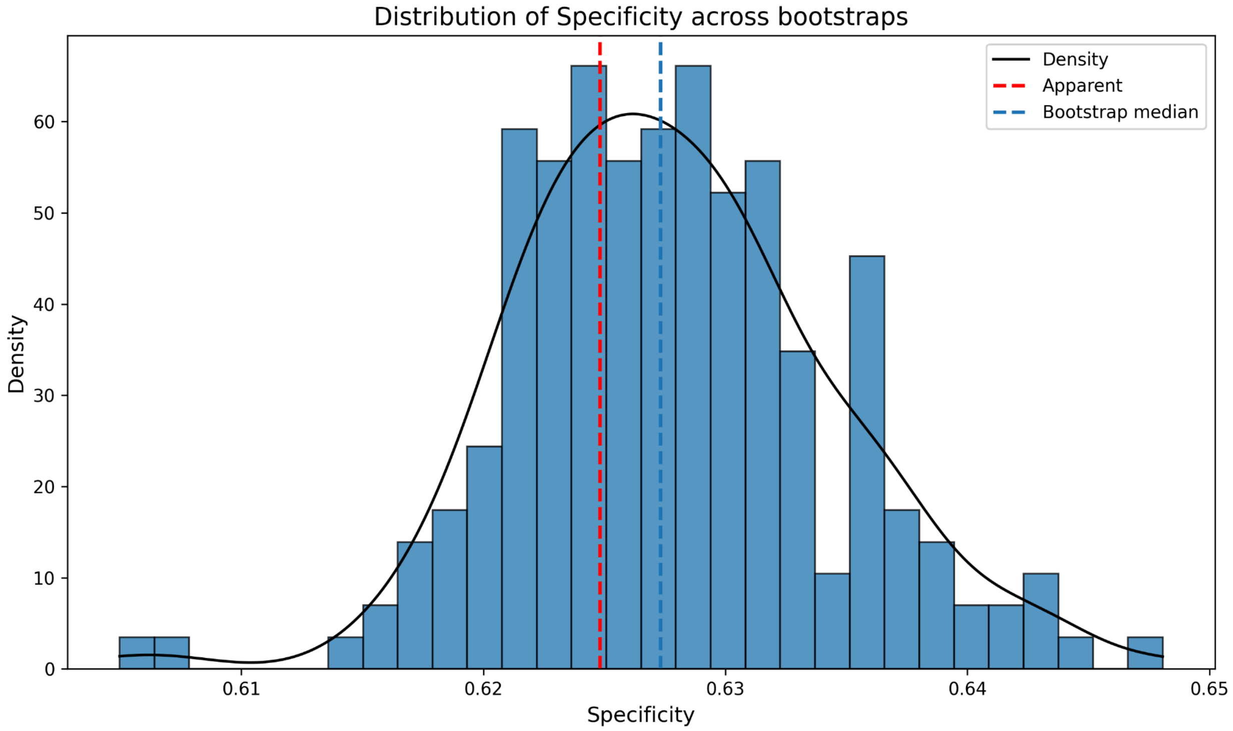

- Brier score

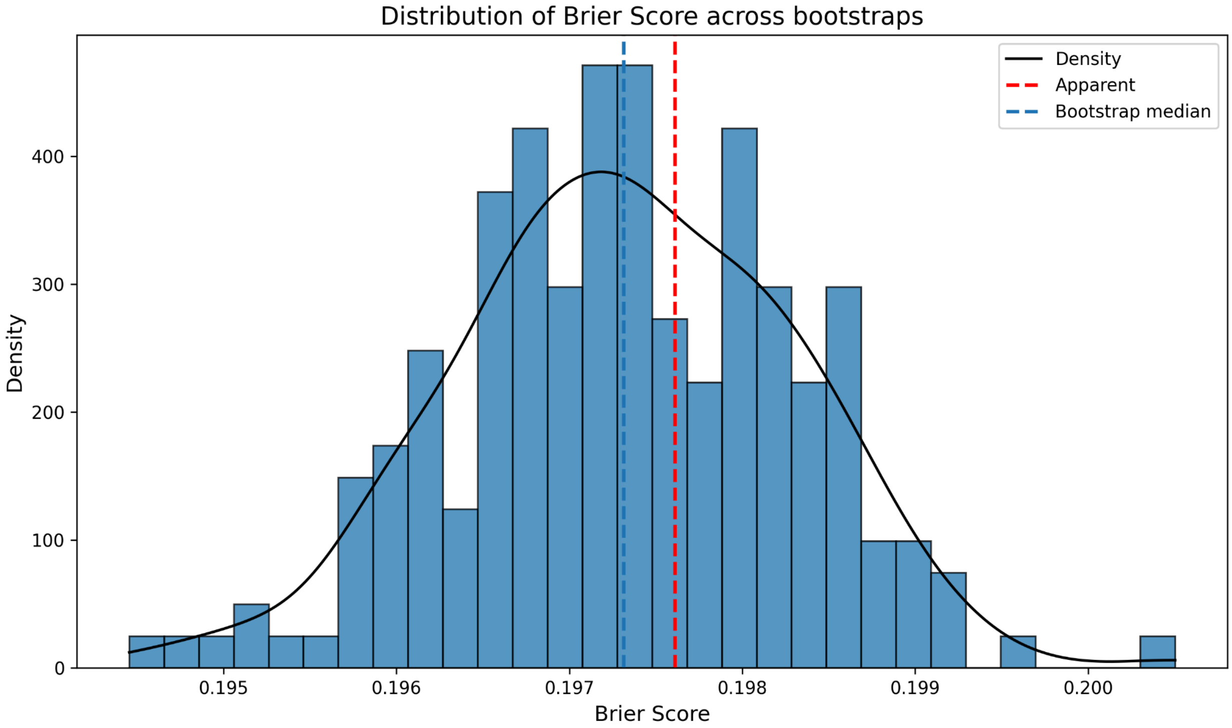


**b) Recovery**

- Calibration curve

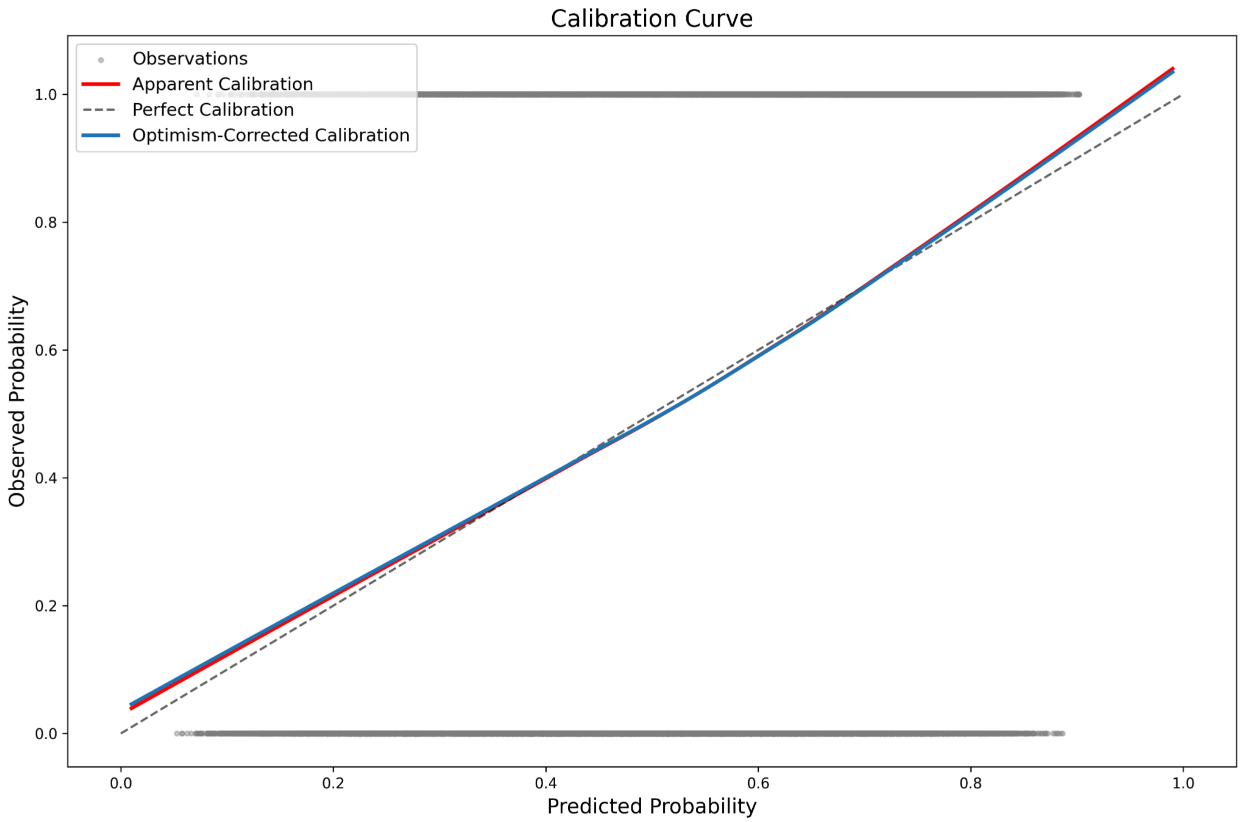


- Prediction instability

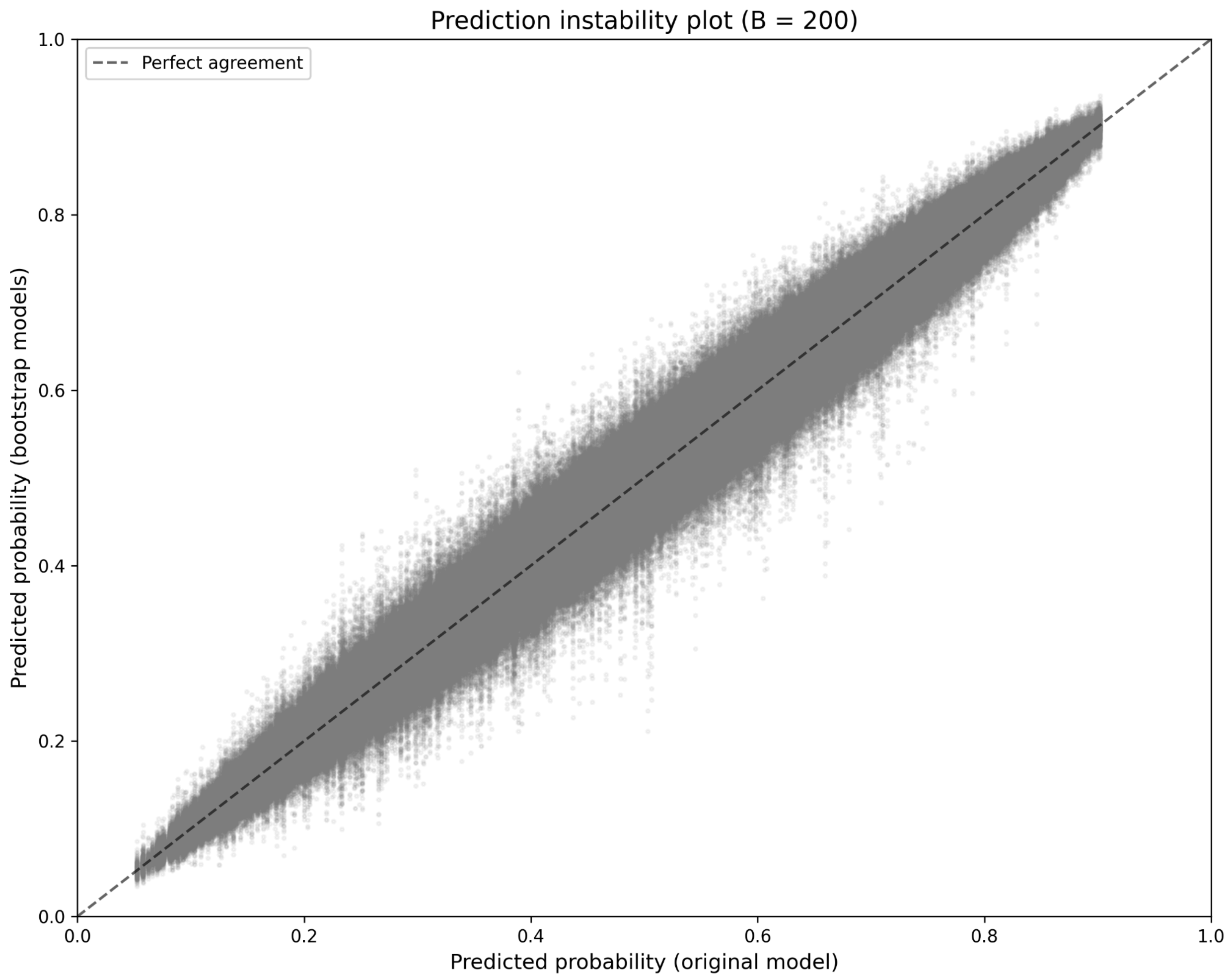


- Calibration instability

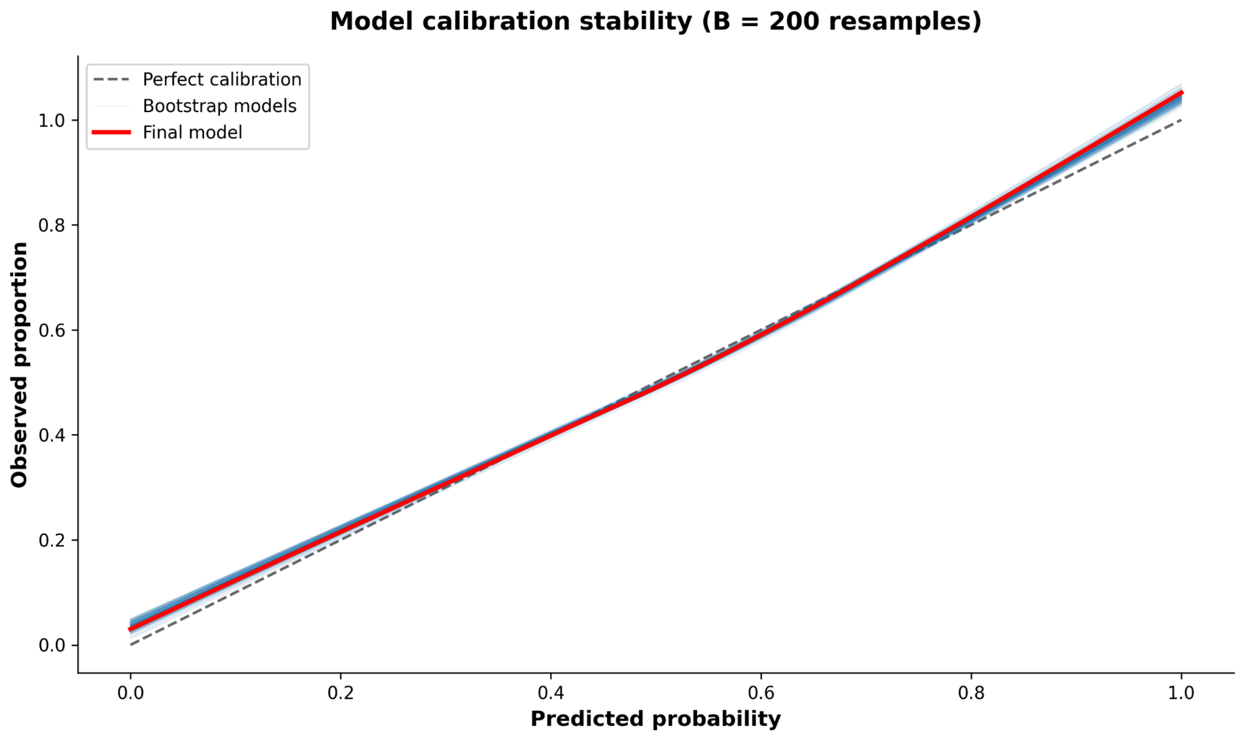

Model calibration stability (B = 200 resamples)
Perfect calibration
Bootstrap models
Final model
Observed proportion
Predicted probability
0.0
0.2
0.4
0.6
0.8
1.0


- MAPE

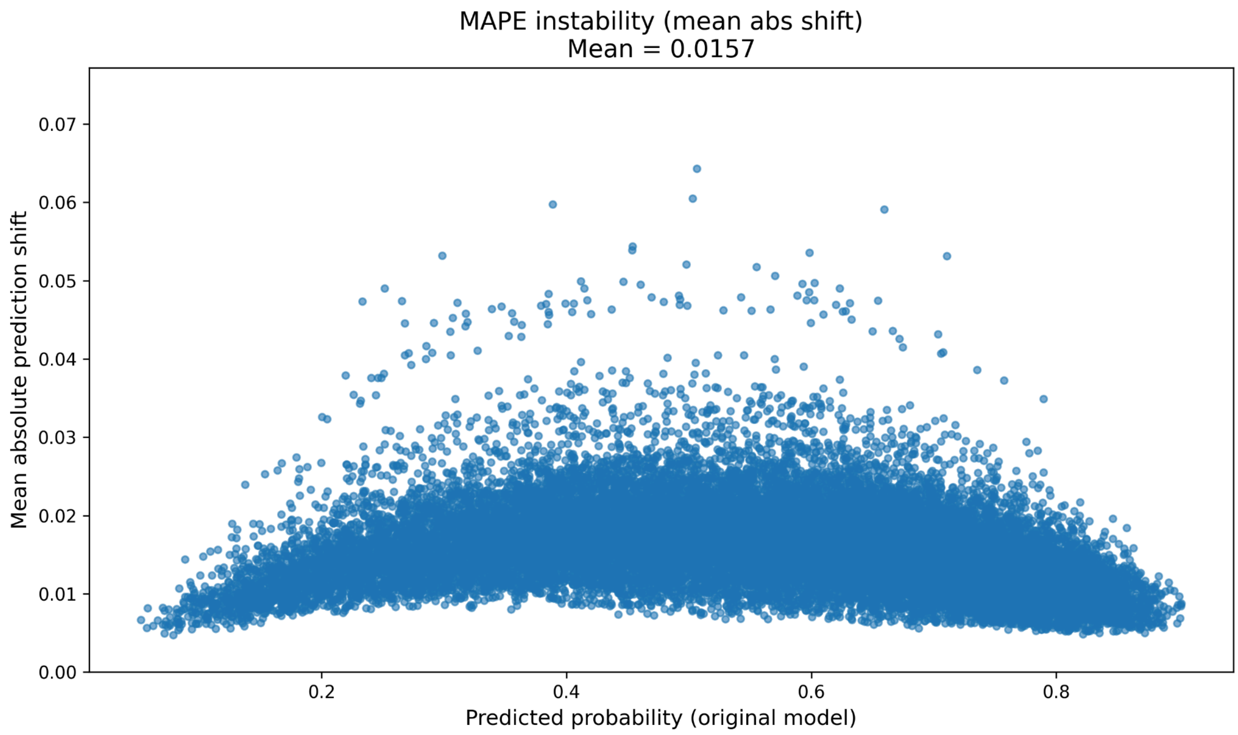


- Optimism

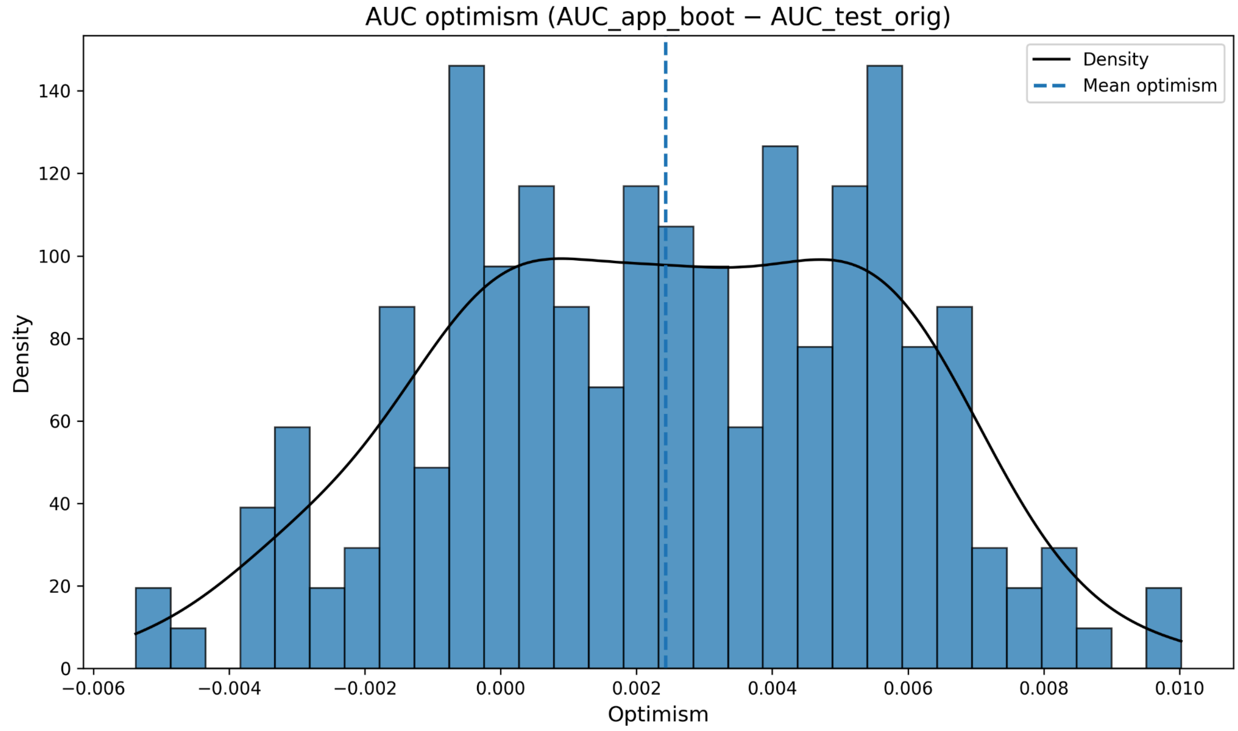


- AUC

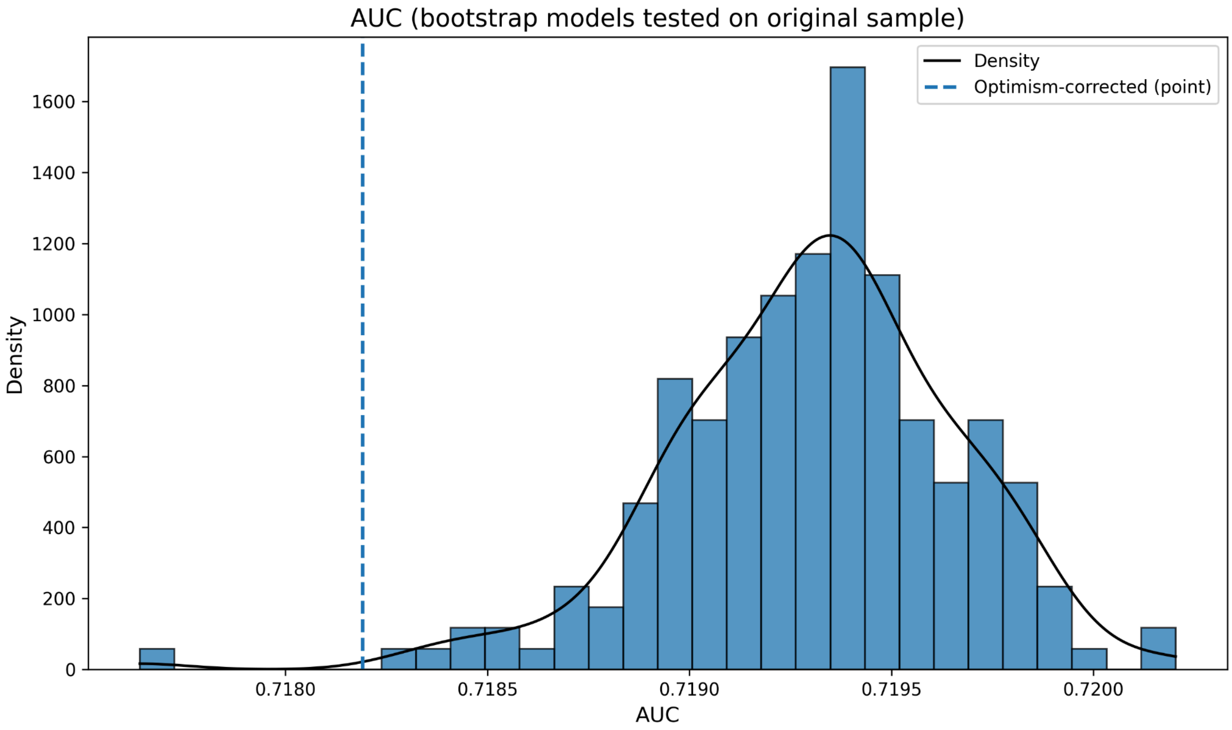


- Sensitivity

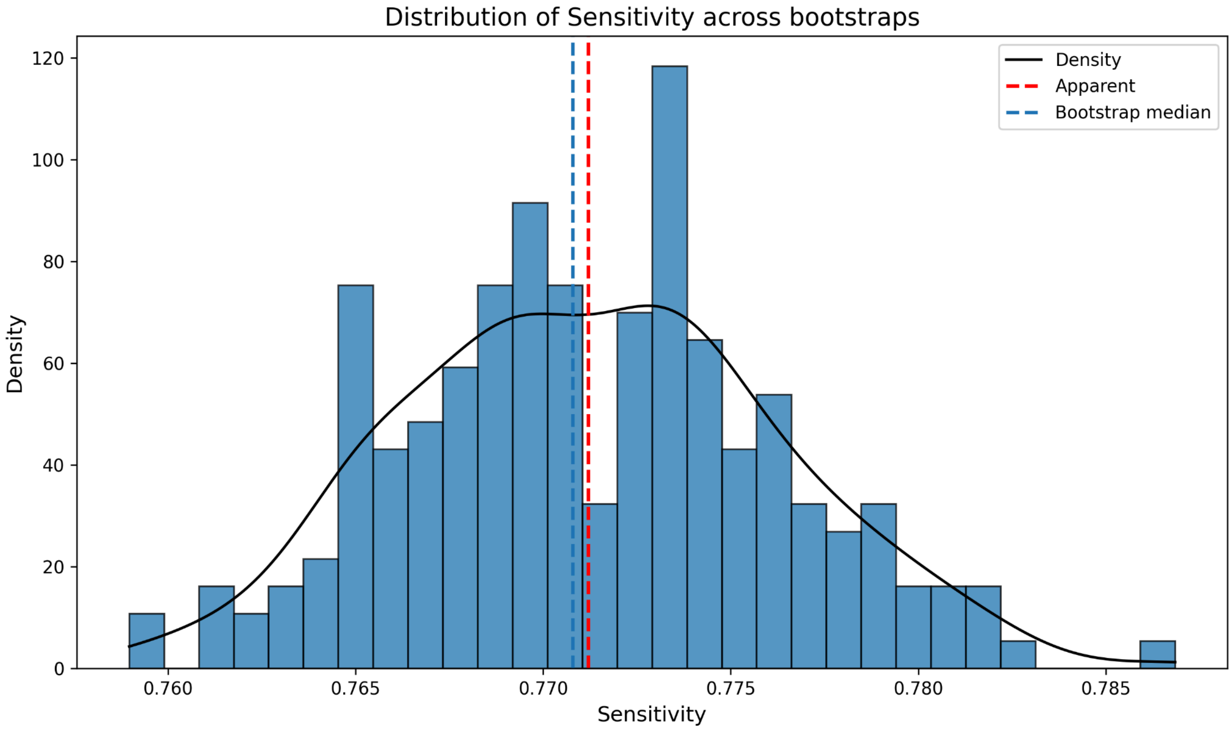


- Specificity

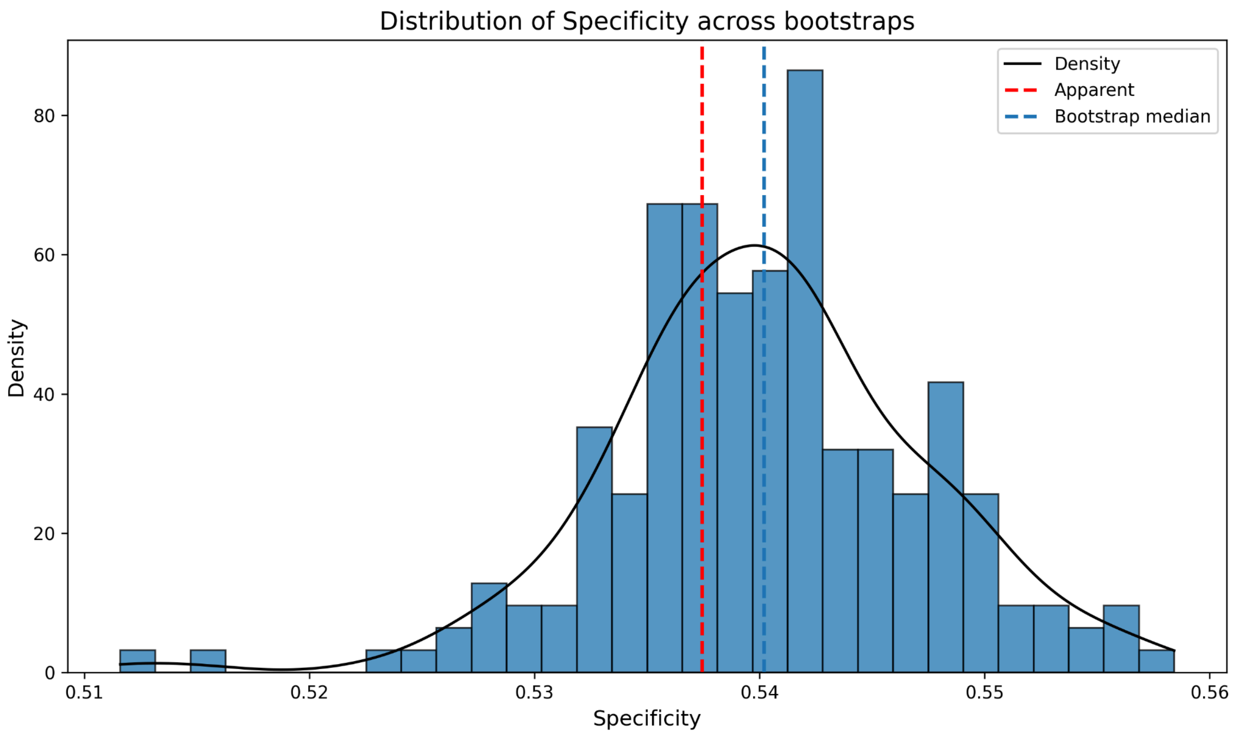


- Brier score

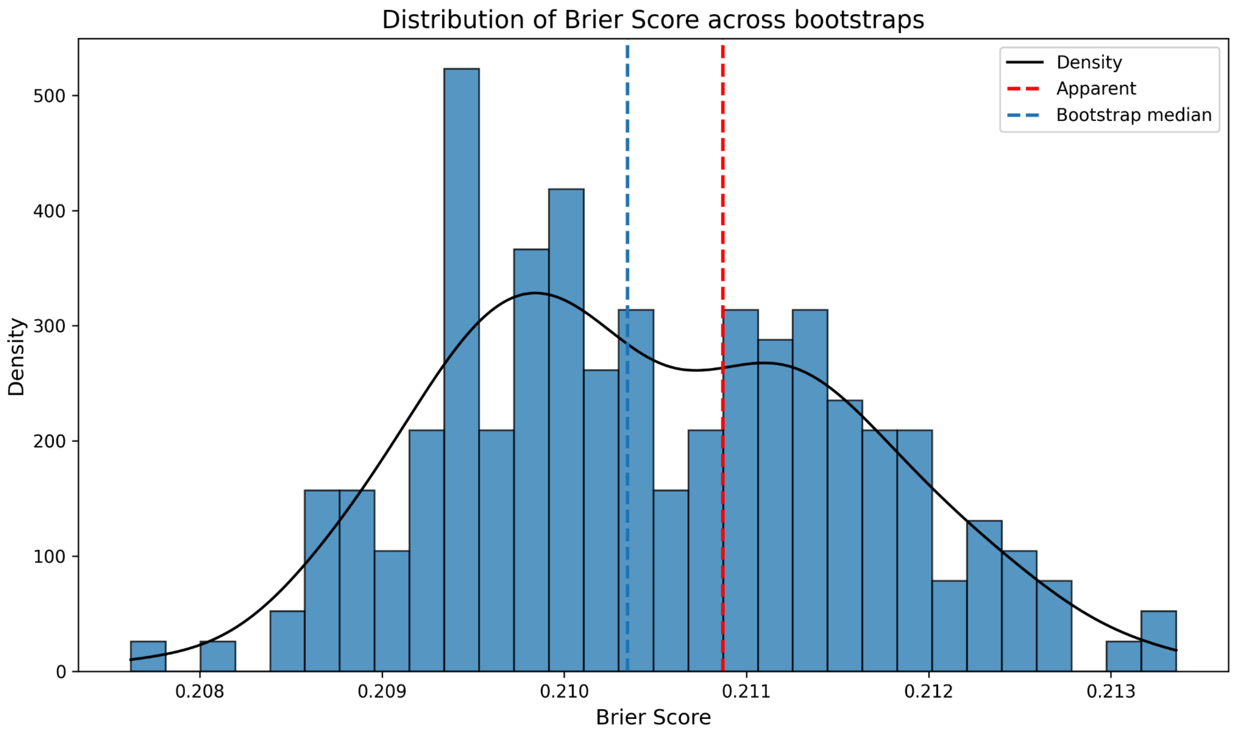


**c) Reliable recovery**

- Calibration curve

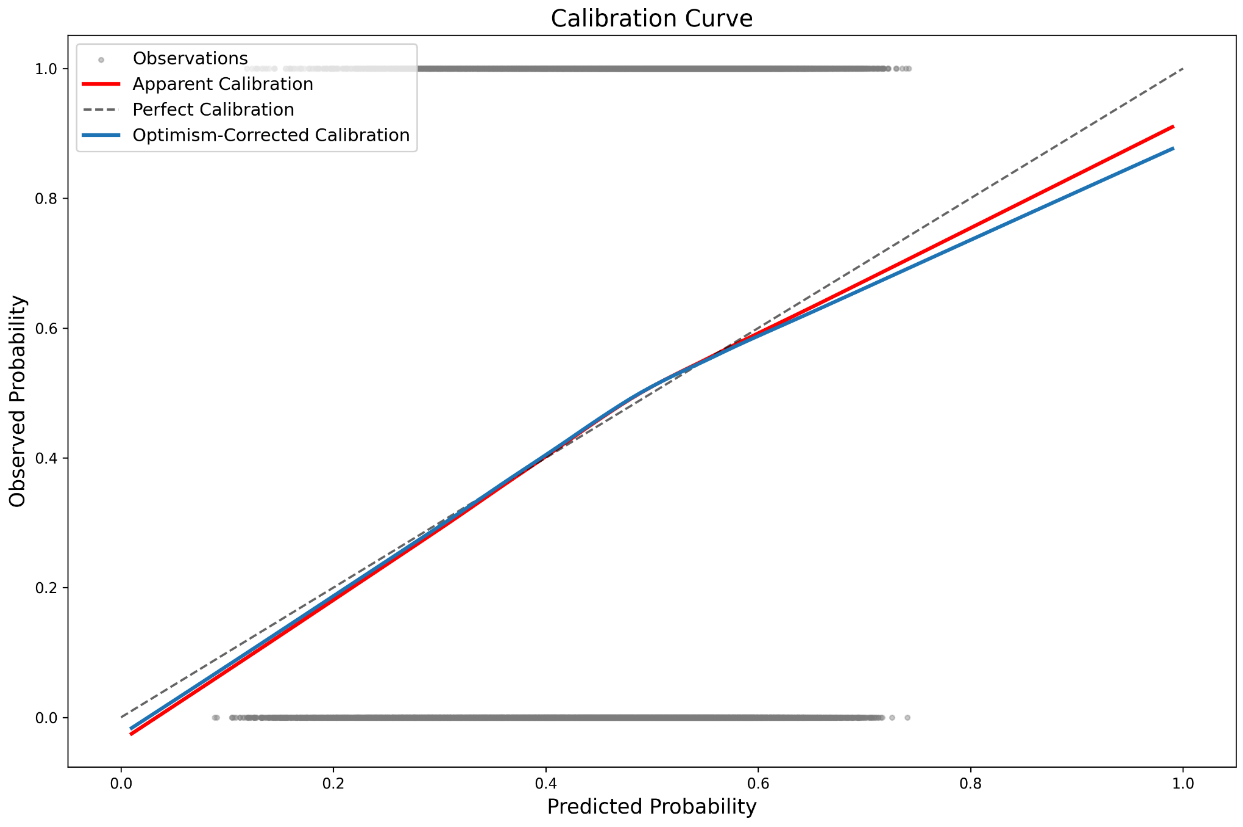


- Prediction instability

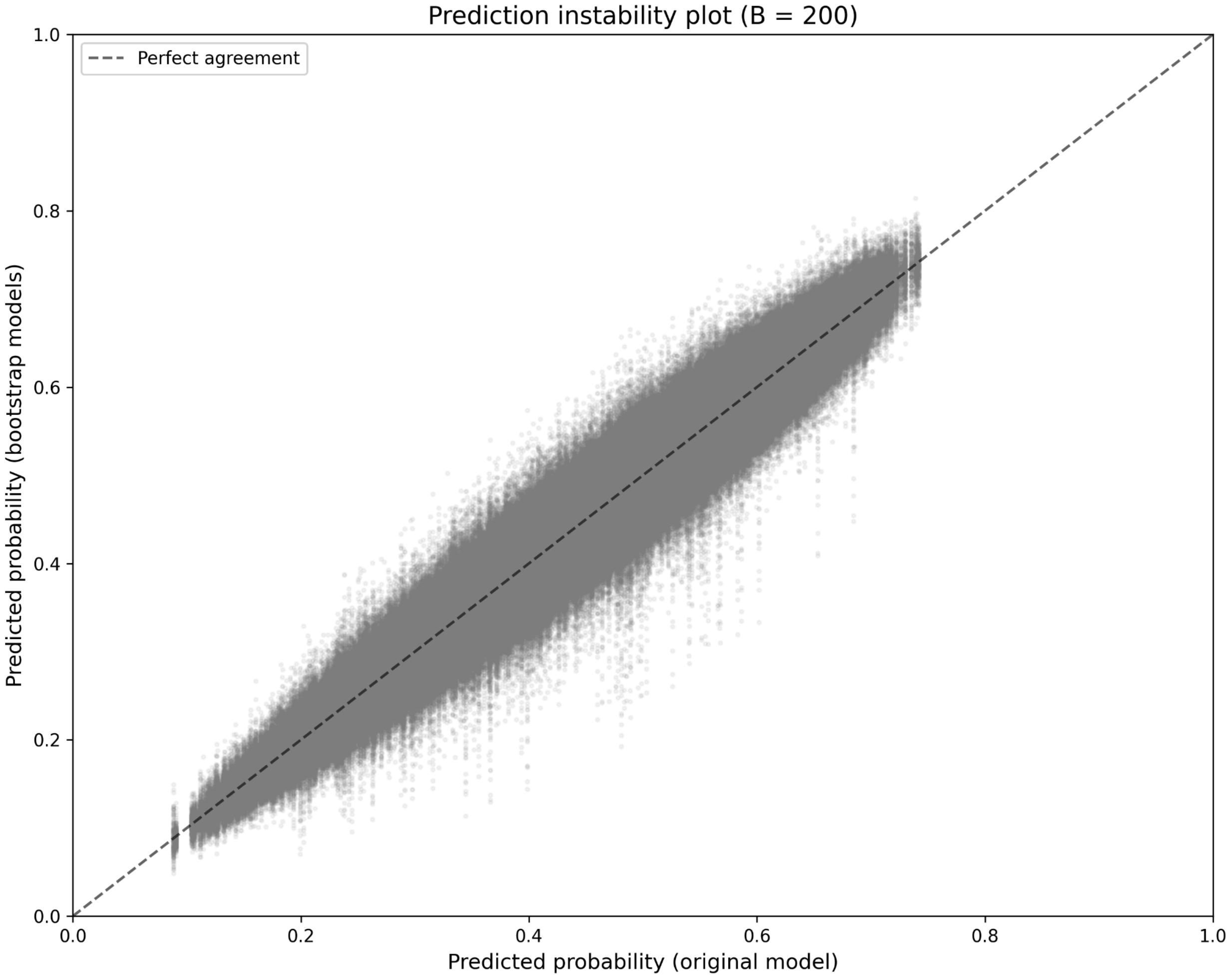


- Calibration instability

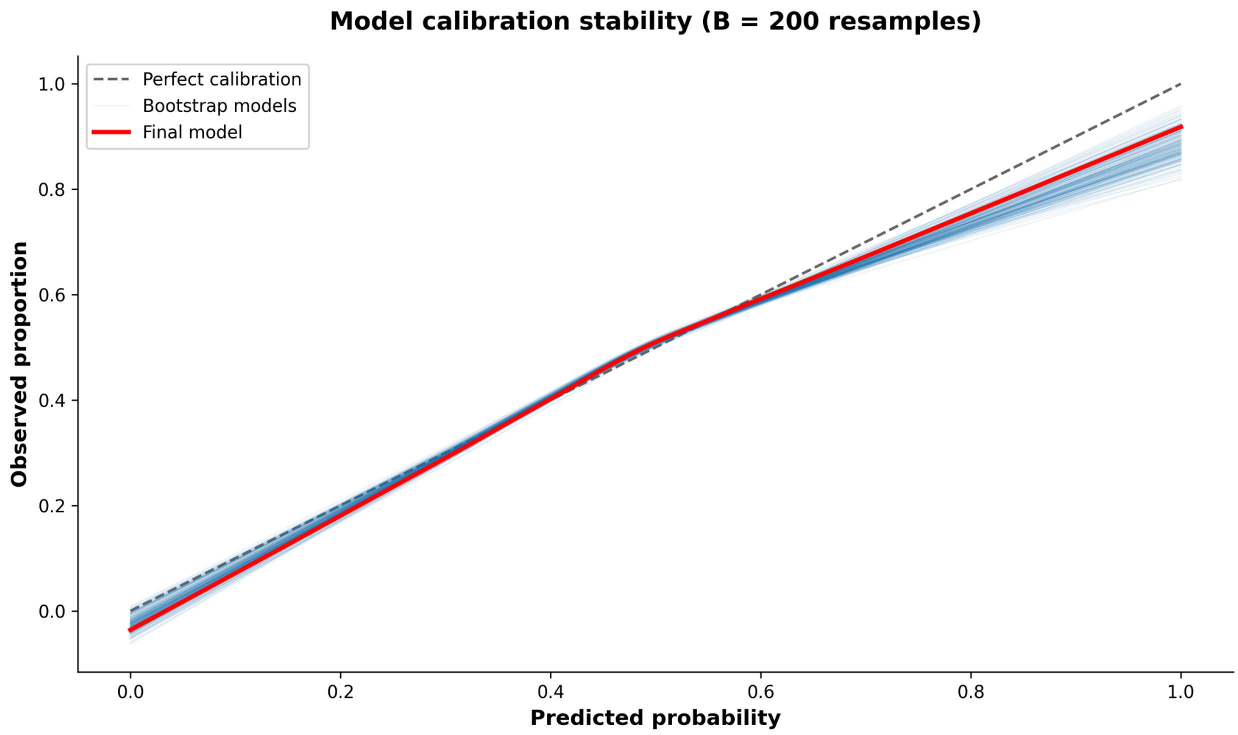

Model calibration stability (B = 200 resamples)
Perfect calibration
Bootstrap models
Final model
Observed proportion
Predicted probability
0.0
0.2
0.4
0.6
0.8
1.0


- MAPE

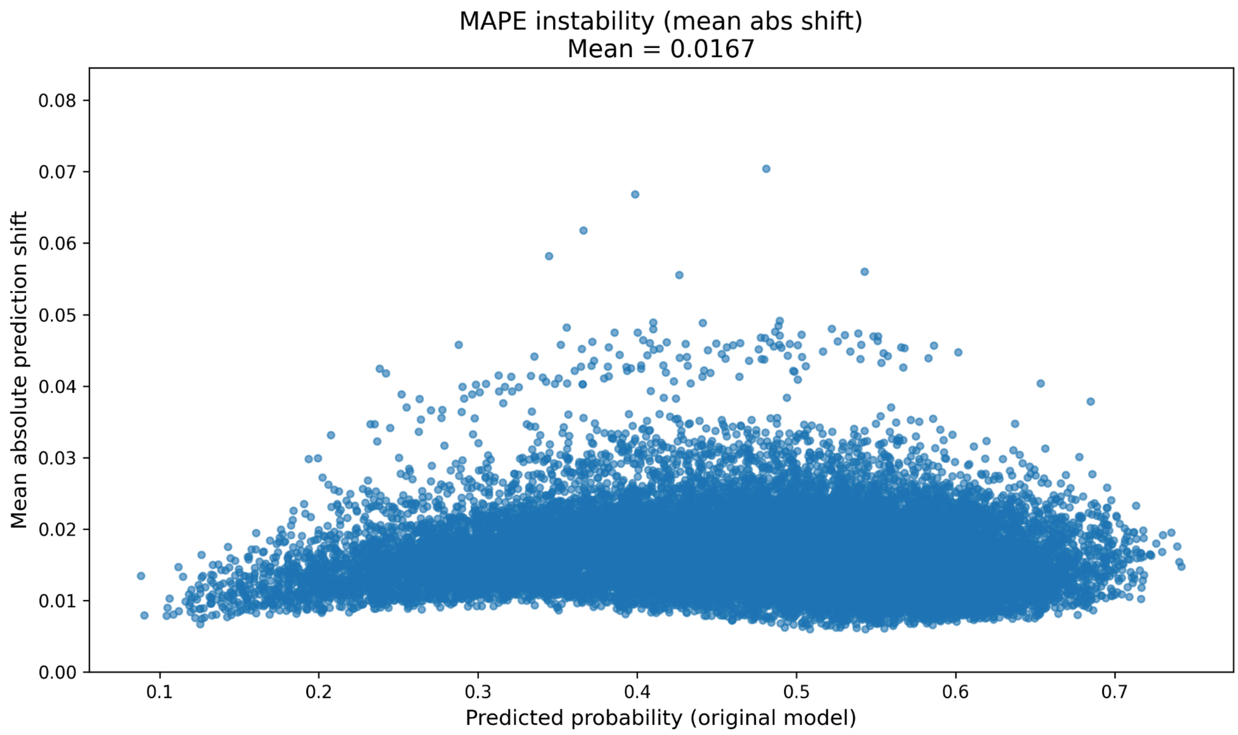


- Optimism

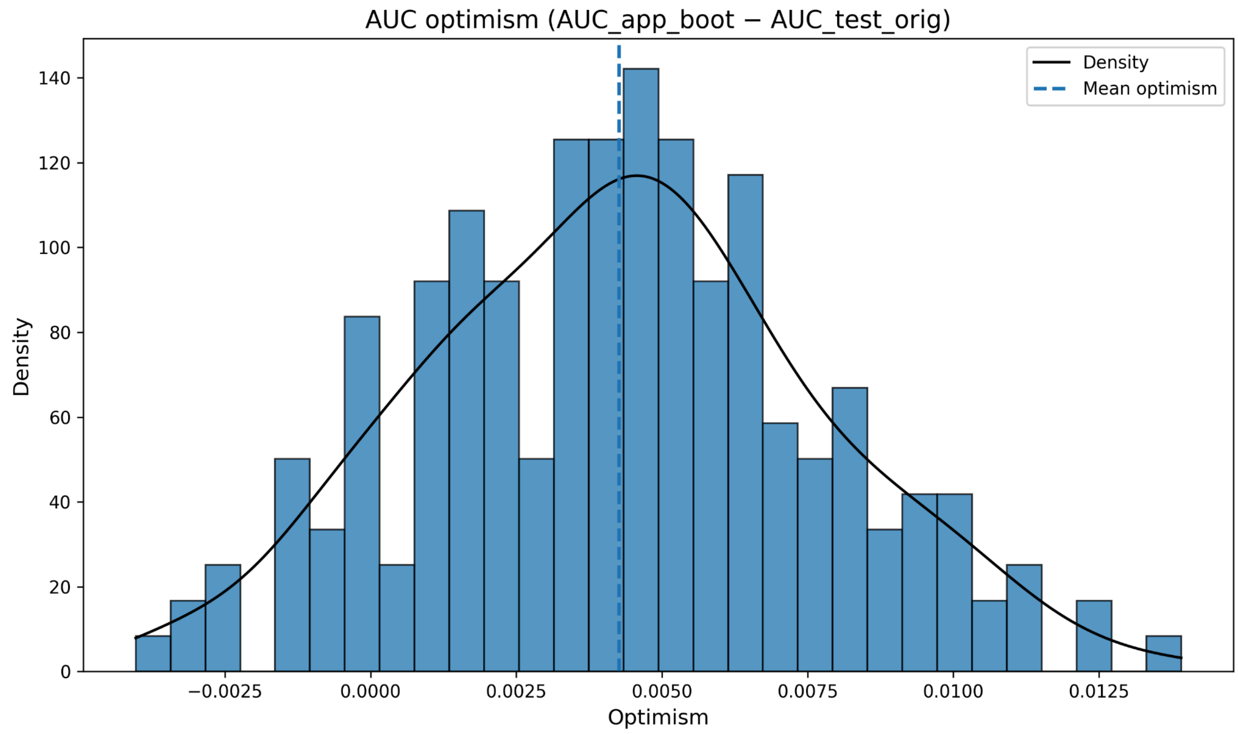


- AUC

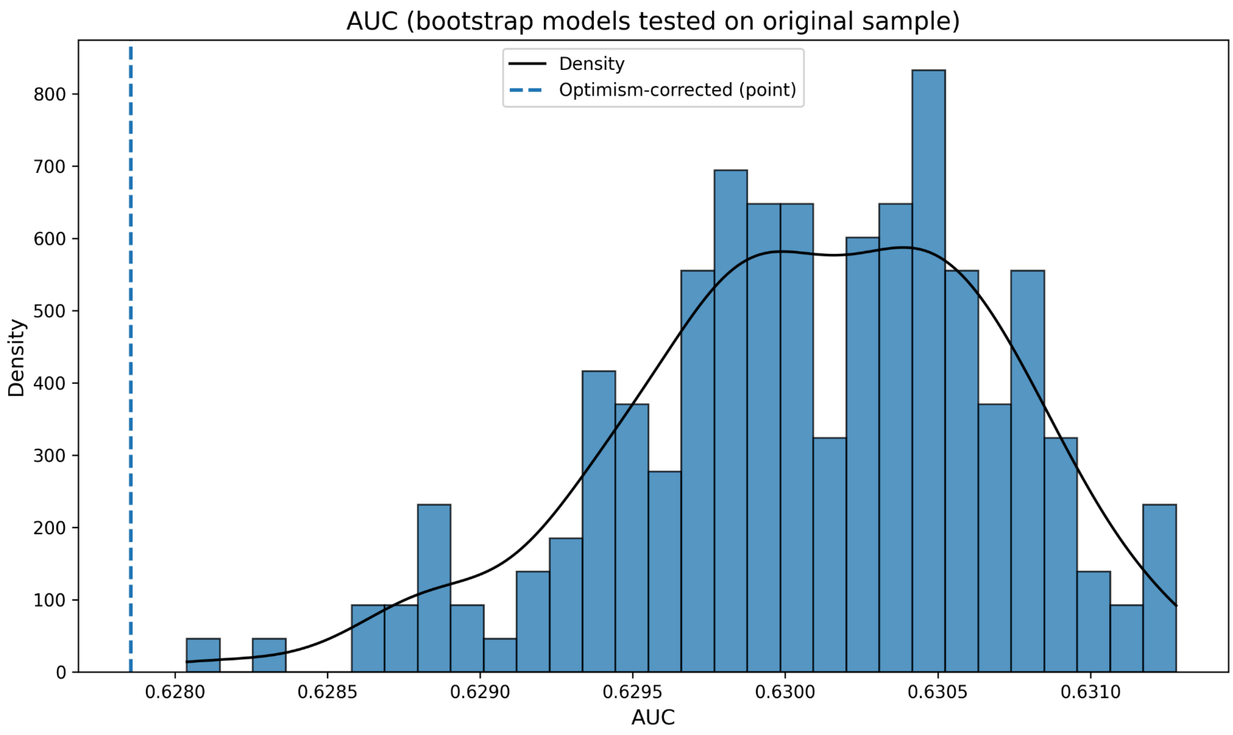


- Sensitivity

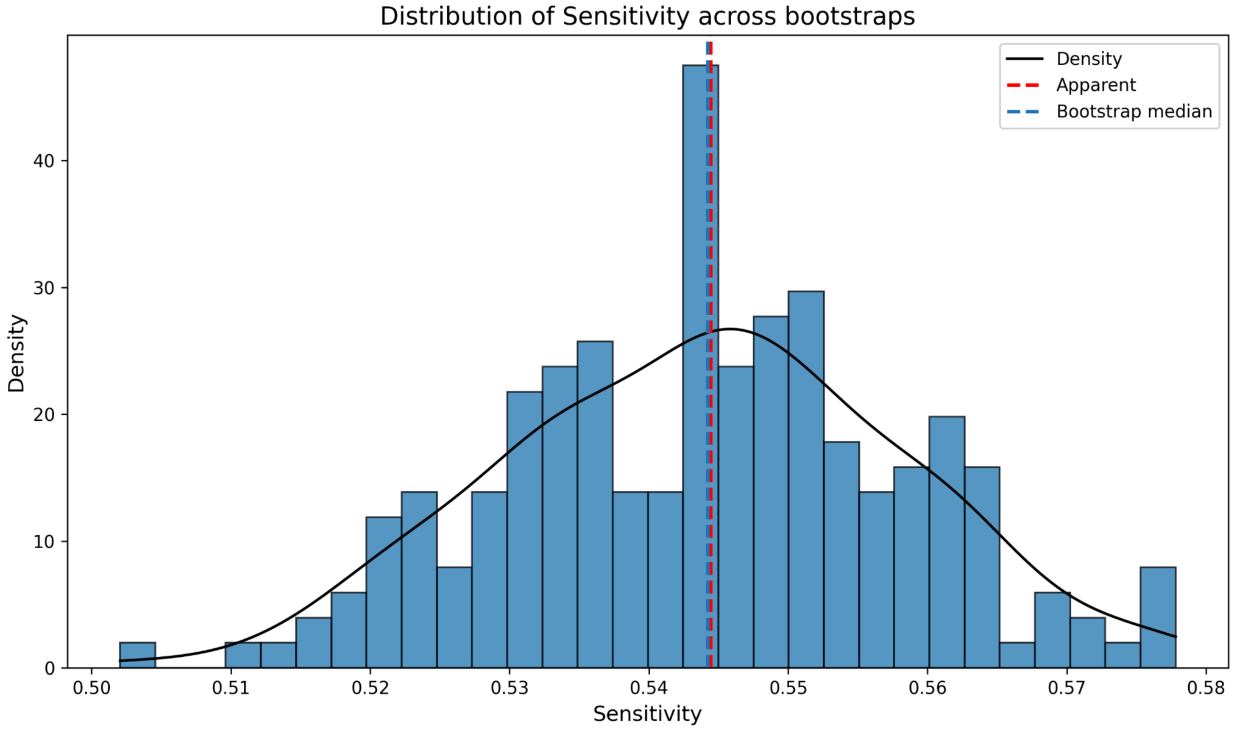


- Specificity

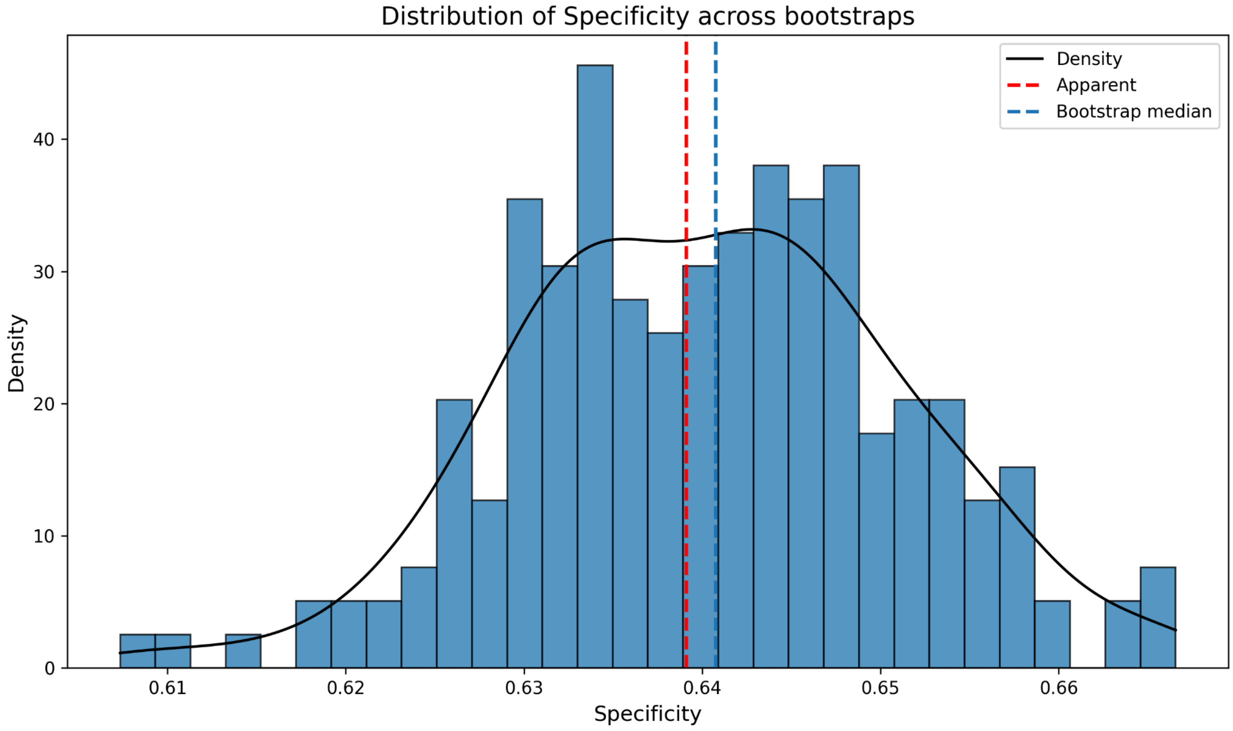
Distribution of Specificity across bootstraps
Density
Apparent
Bootstrap median
0
10
20
30
40
Density
0.61
0.62
0.63
0.64
0.65
0.66
Specificity

- Brier score

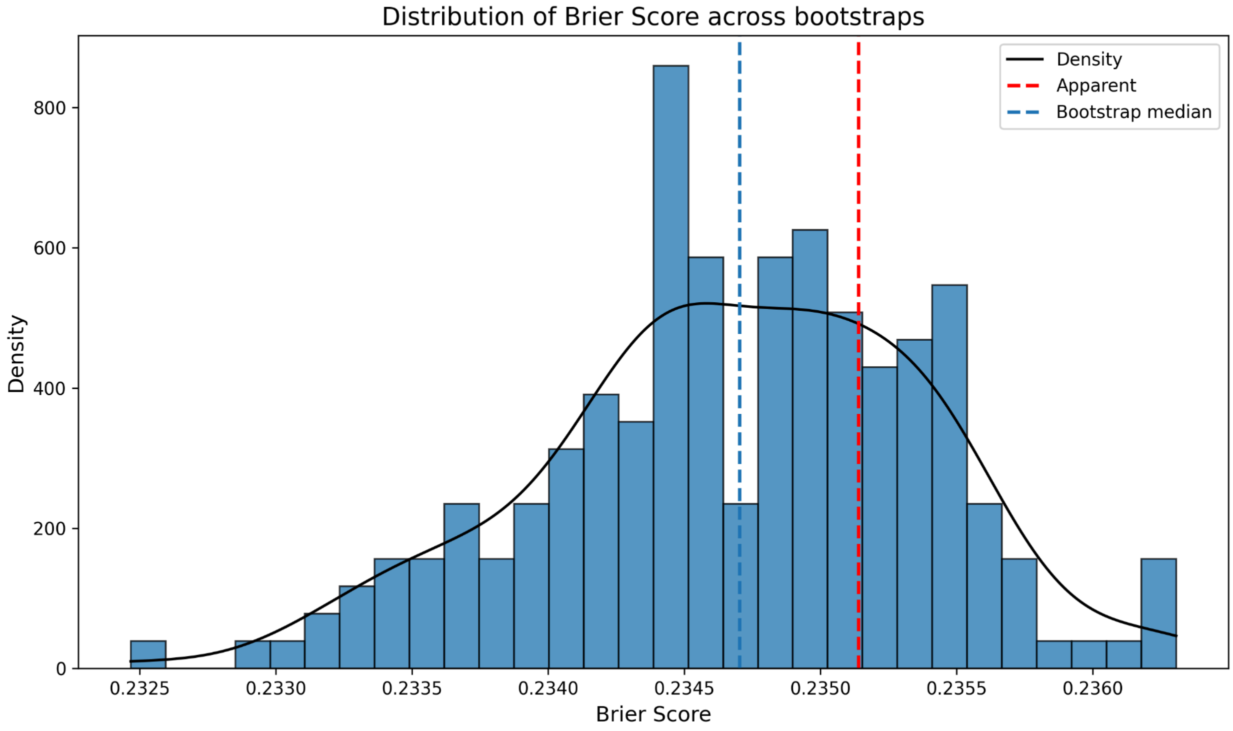
Distribution of Brier Score across bootstraps
Density
Apparent
Bootstrap median
0
200
400
600
800
Density
0.2325
0.2330
0.2335
0.2340
0.2345
0.2350
0.2355
0.2360
Brier Score

**Figure 7: Bootstrap results for anxiety outcomes**

a) **Reliable improvement**

- Calibration curve

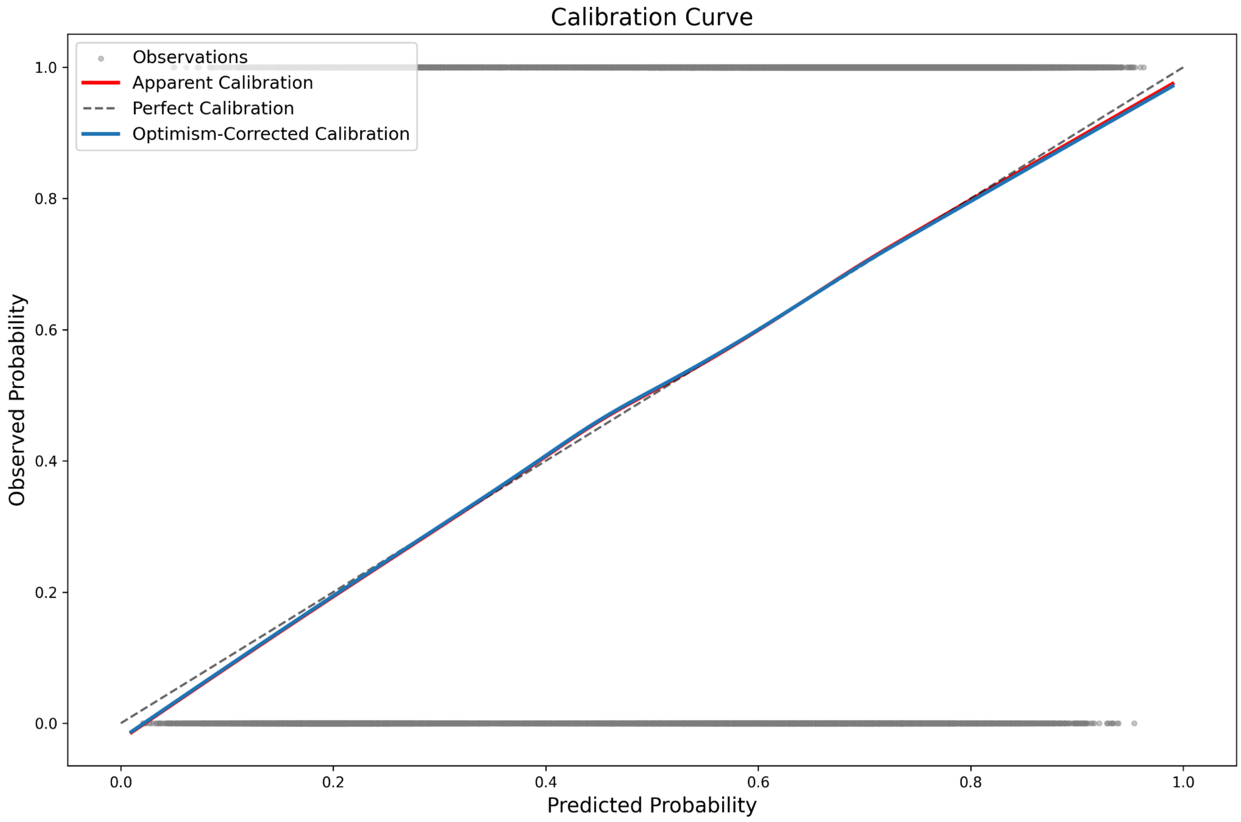

- Prediction instability

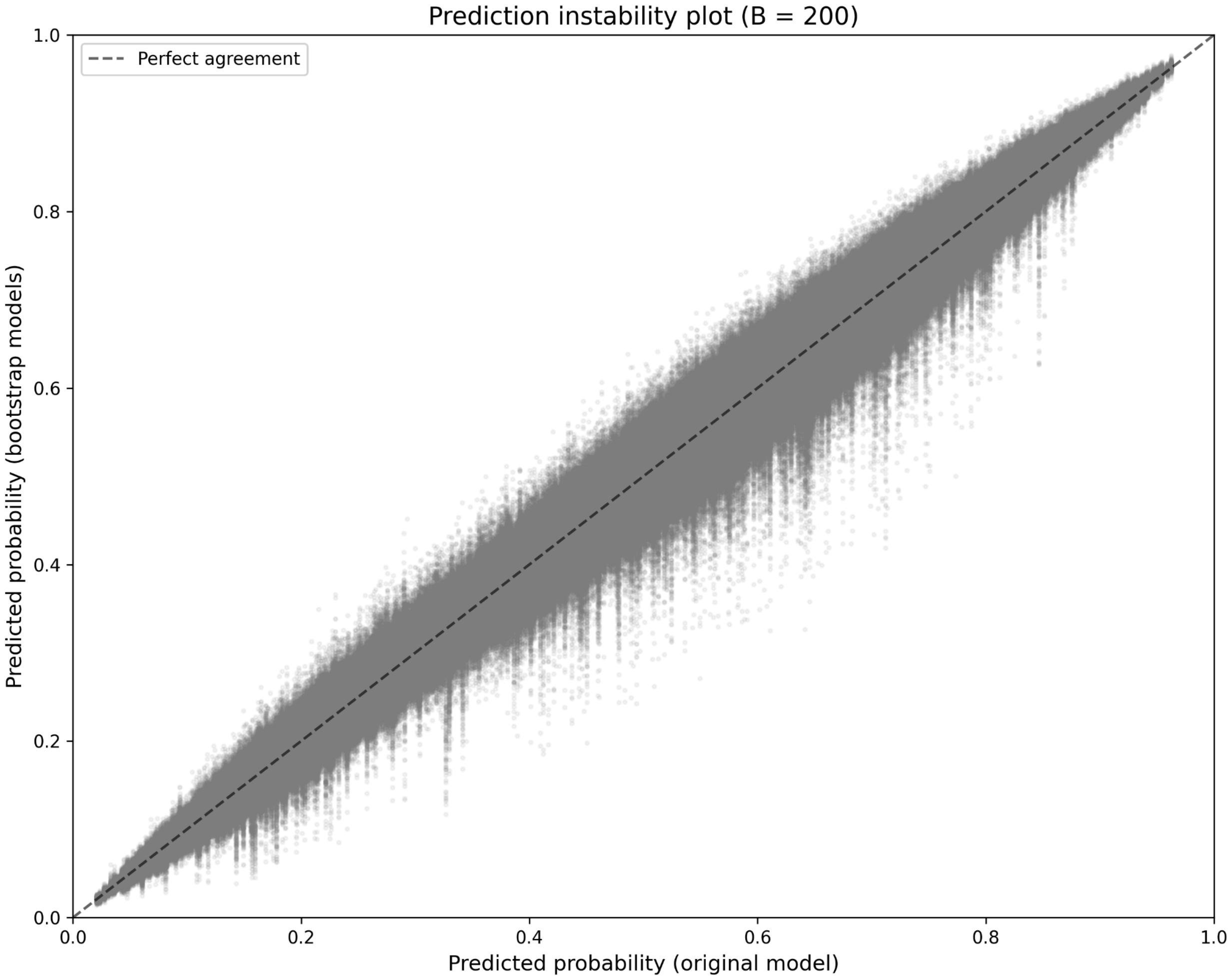

- Calibration instability

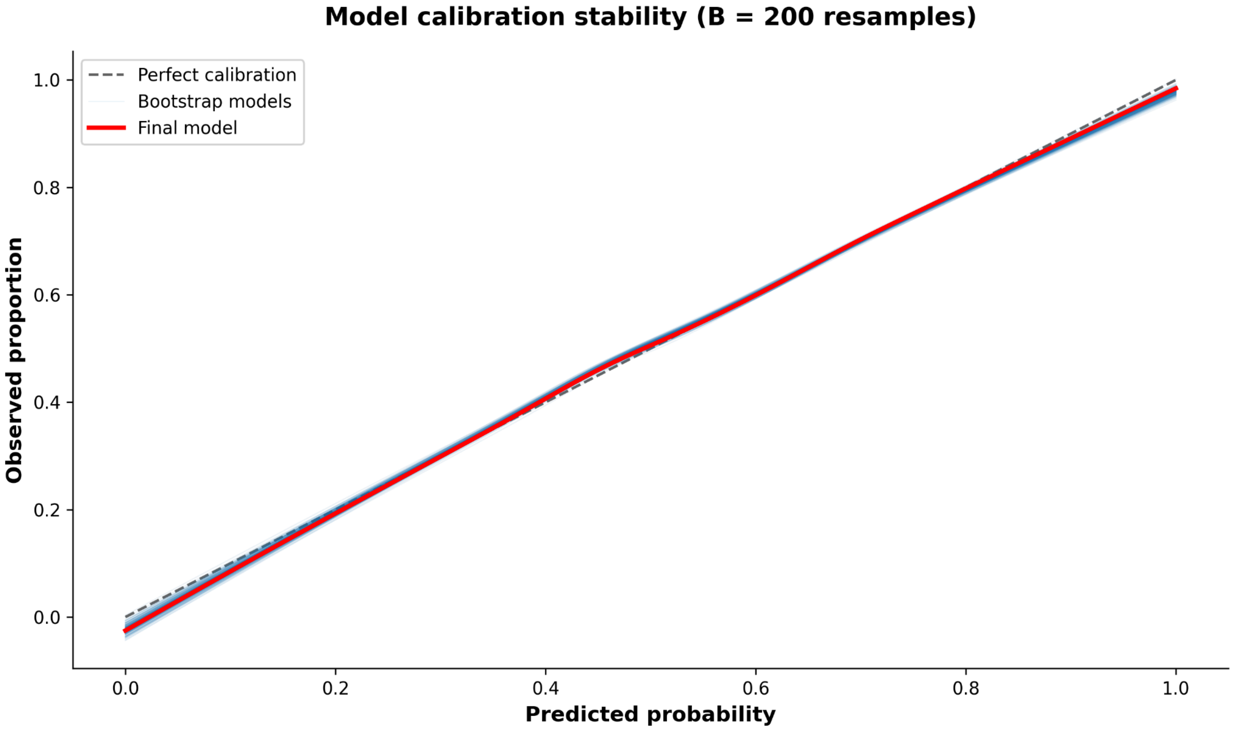

- MAPE

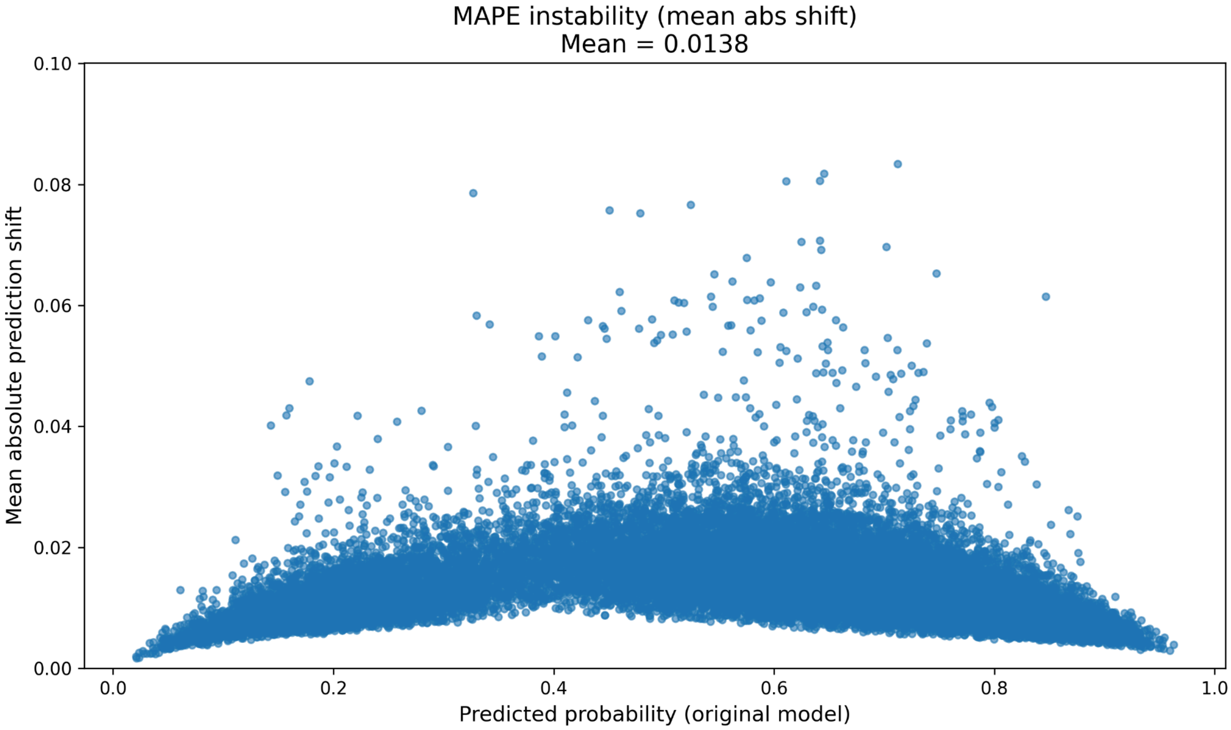


- Optimism

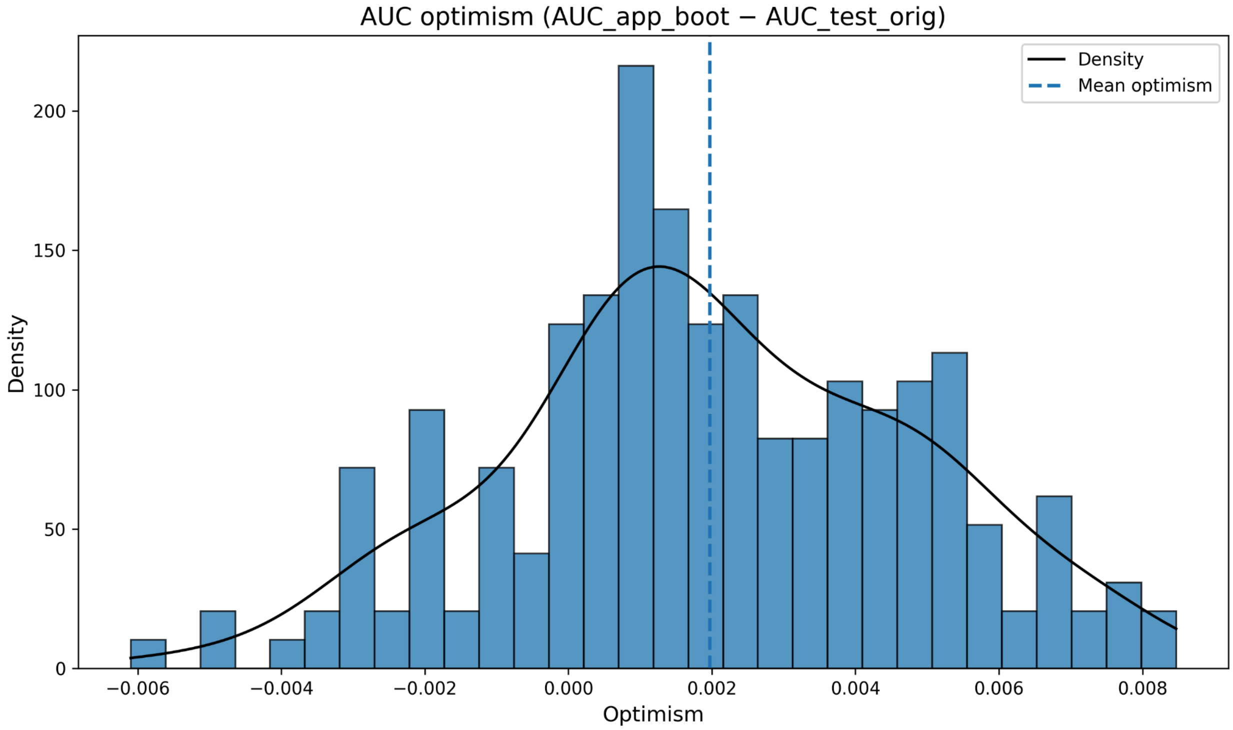

- AUC

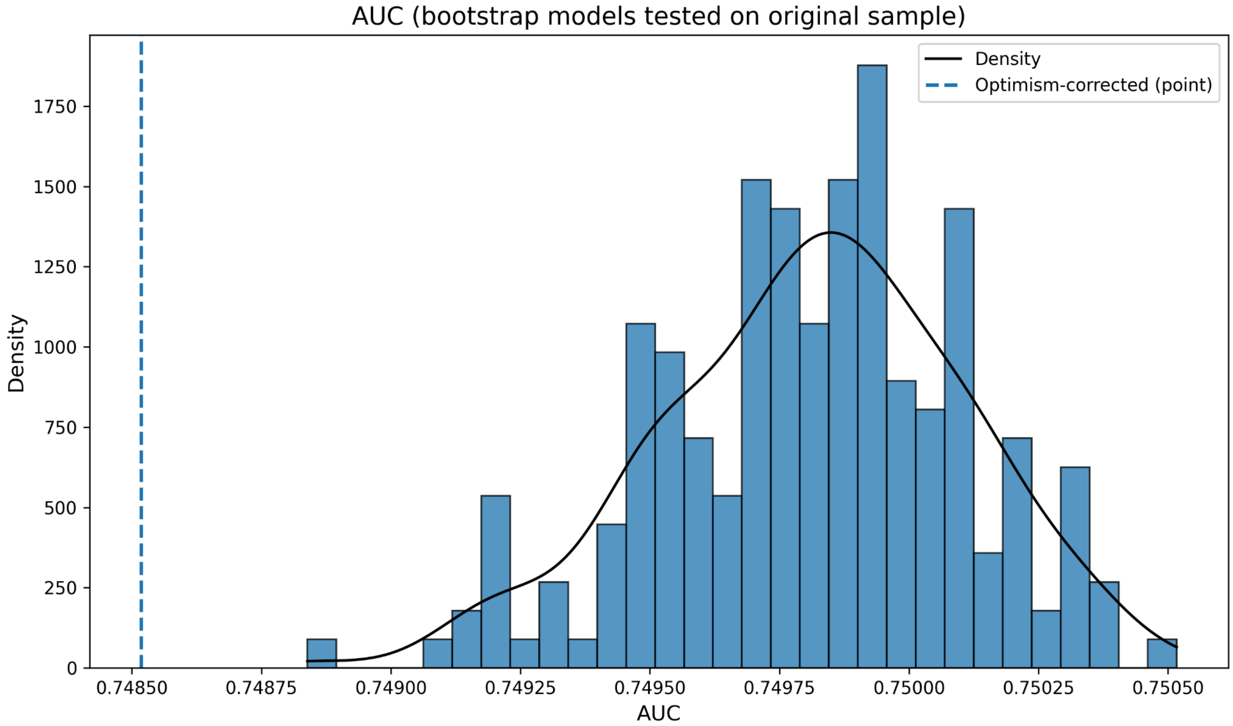


- Sensitivity

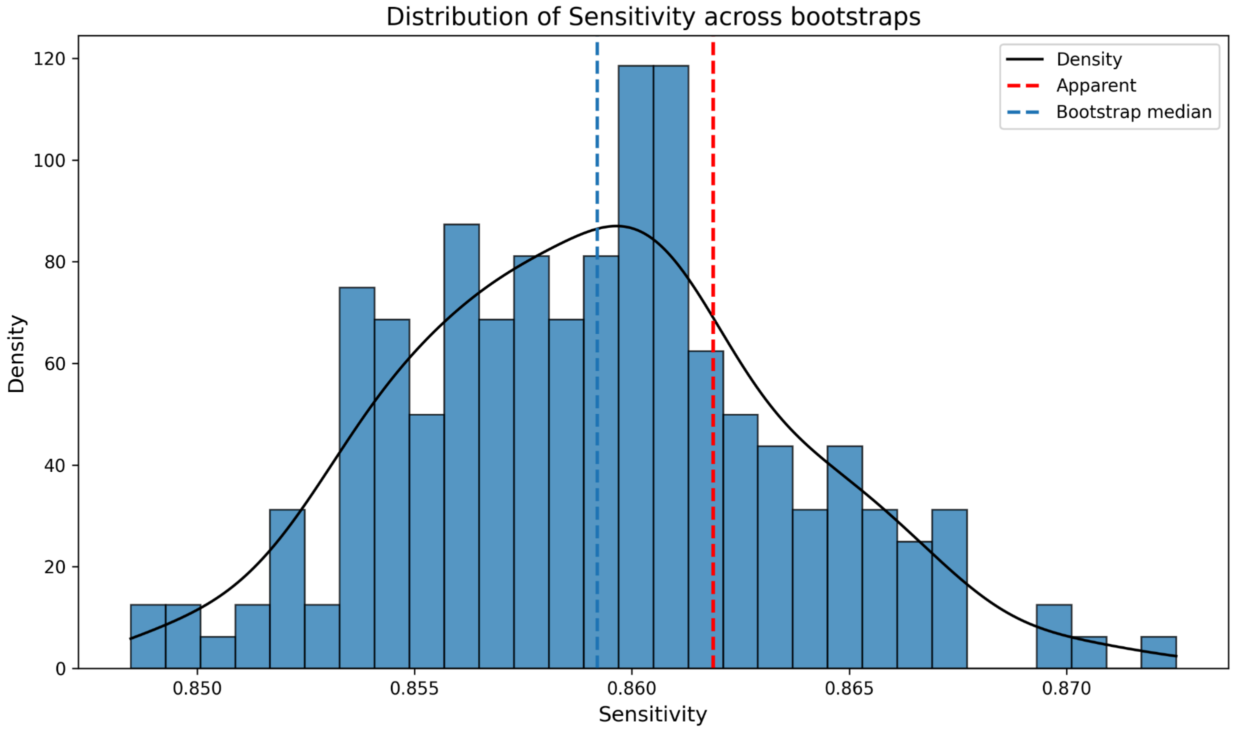

- Specificity

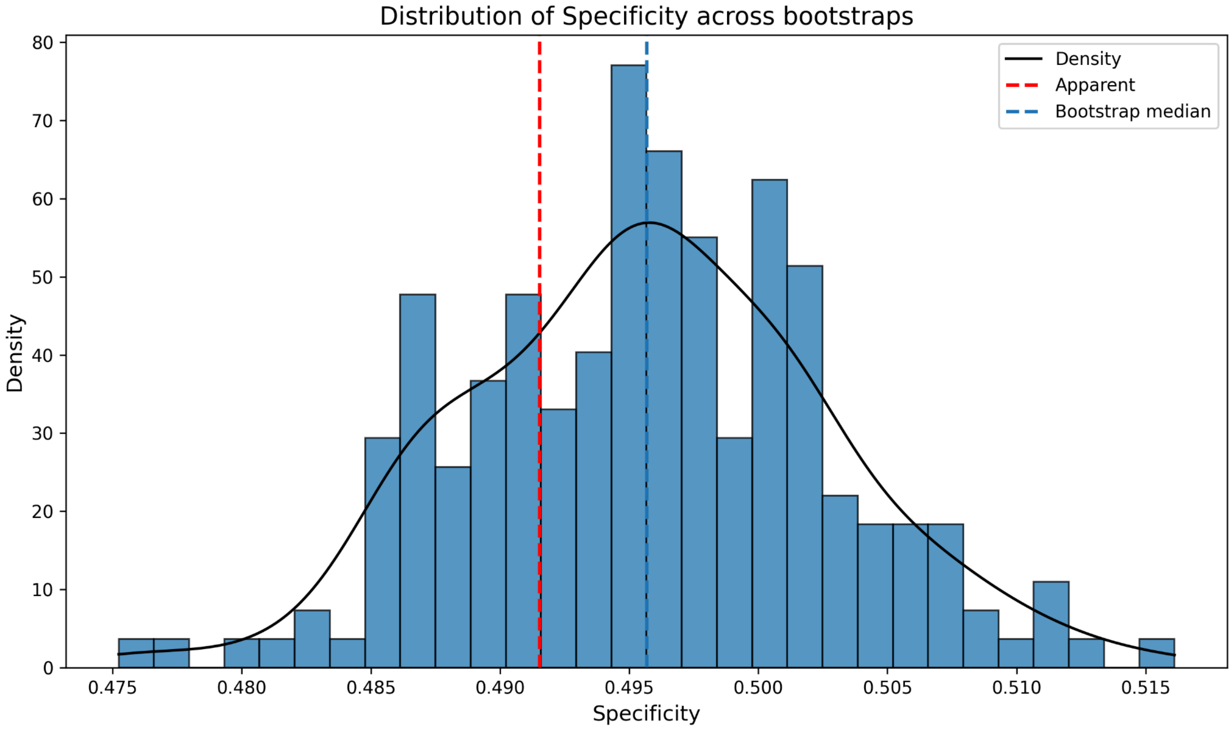


- Brier score

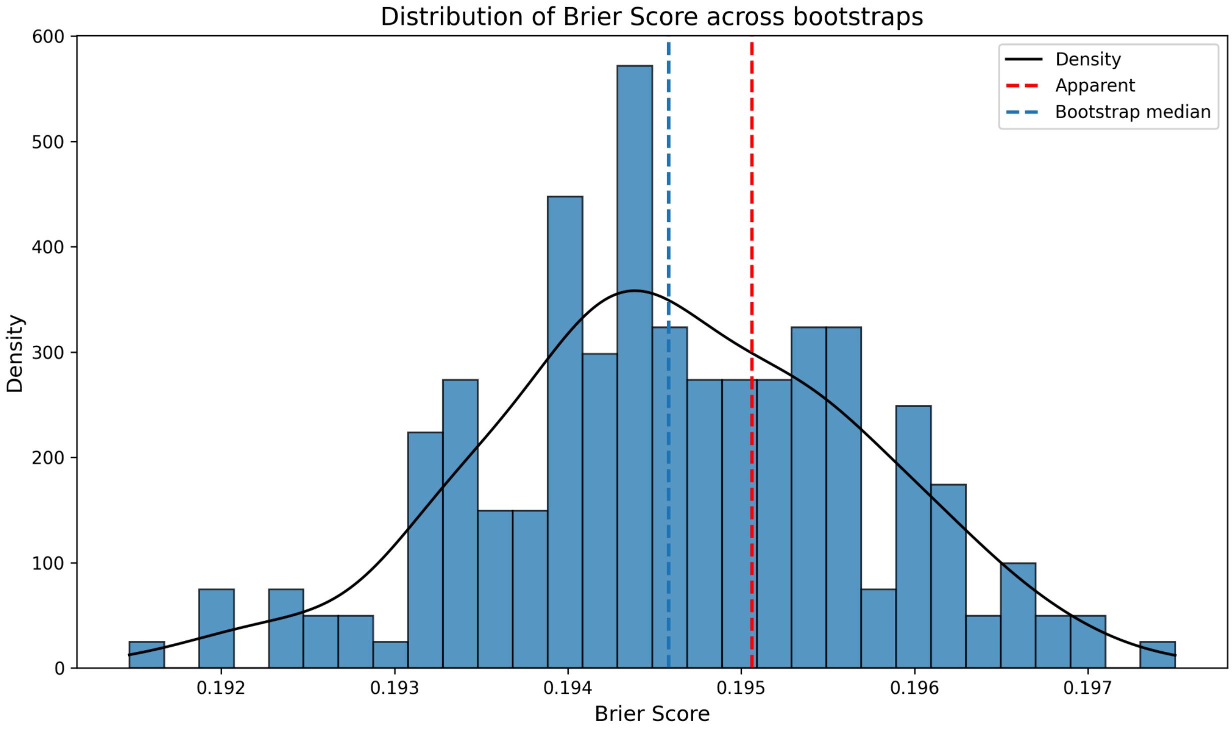

**b) Recovery**

- Calibration curve

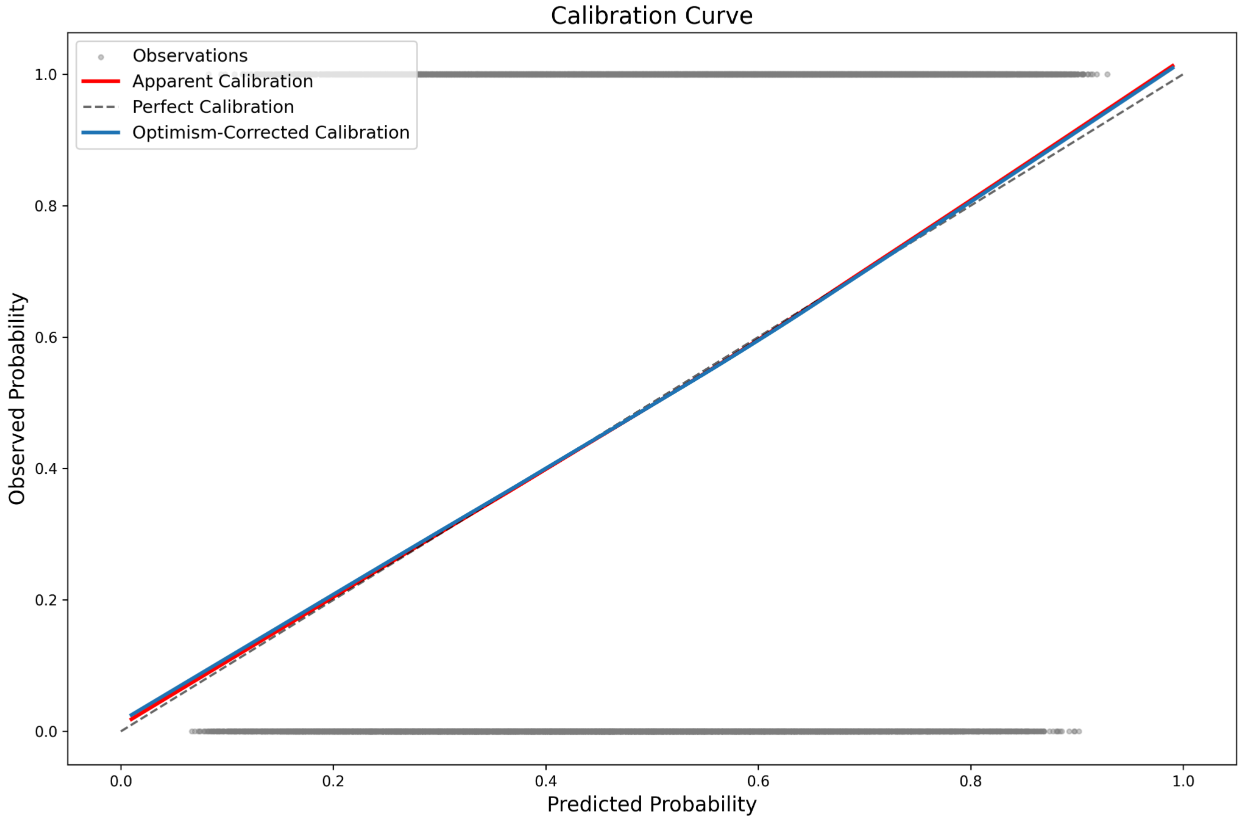

- Prediction instability

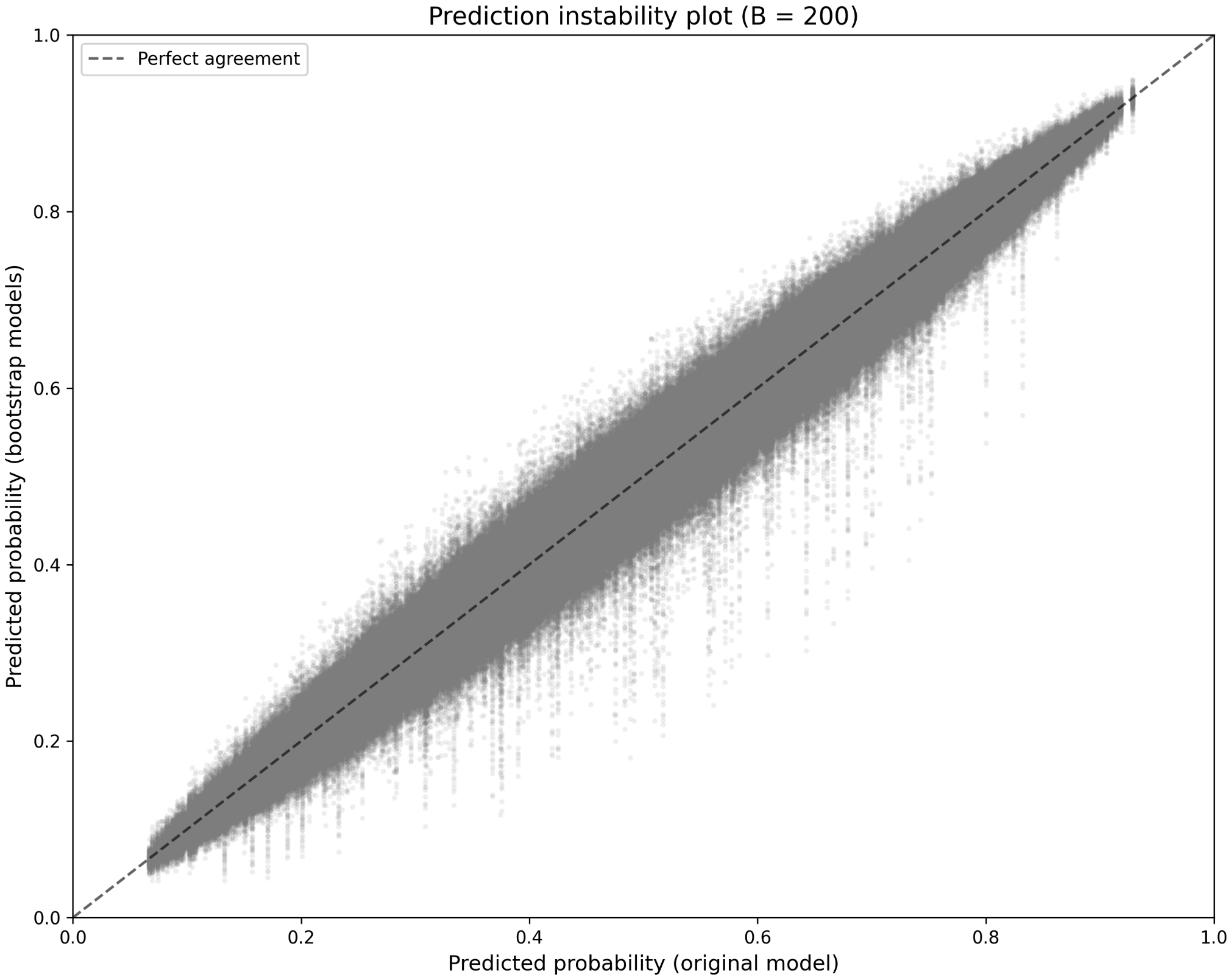

- Calibration instability

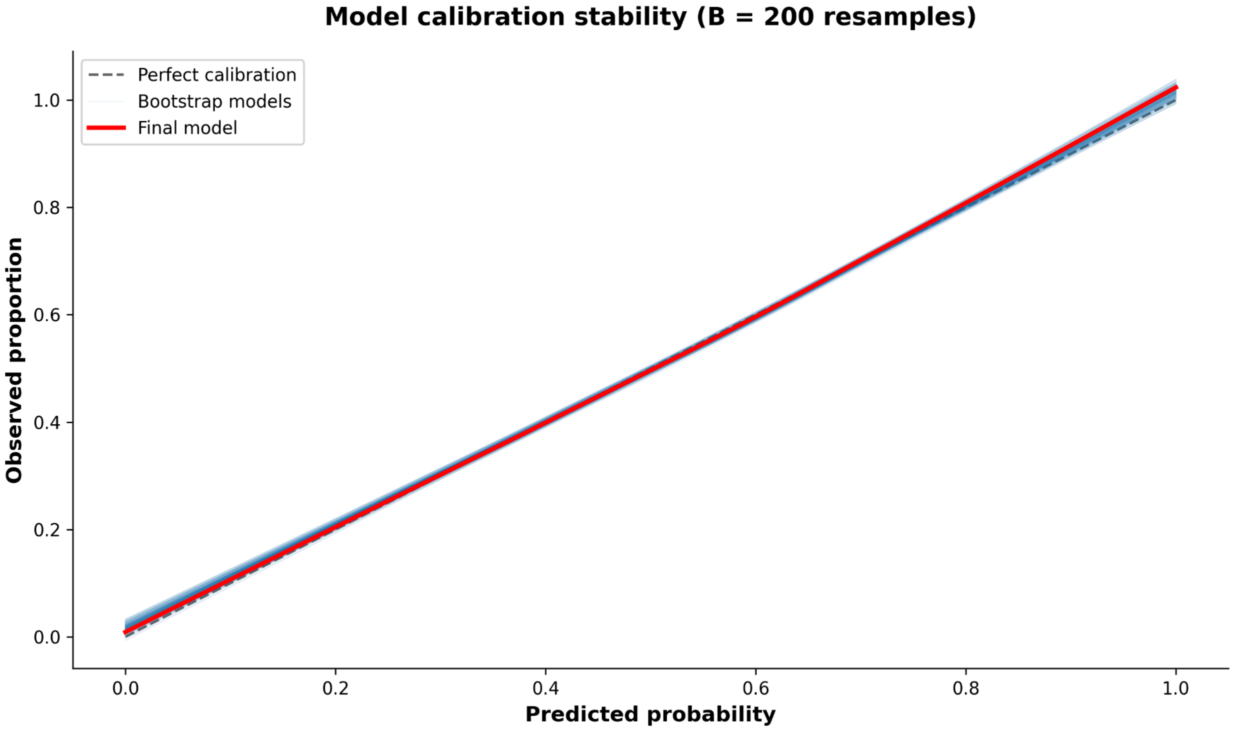

- MAPE

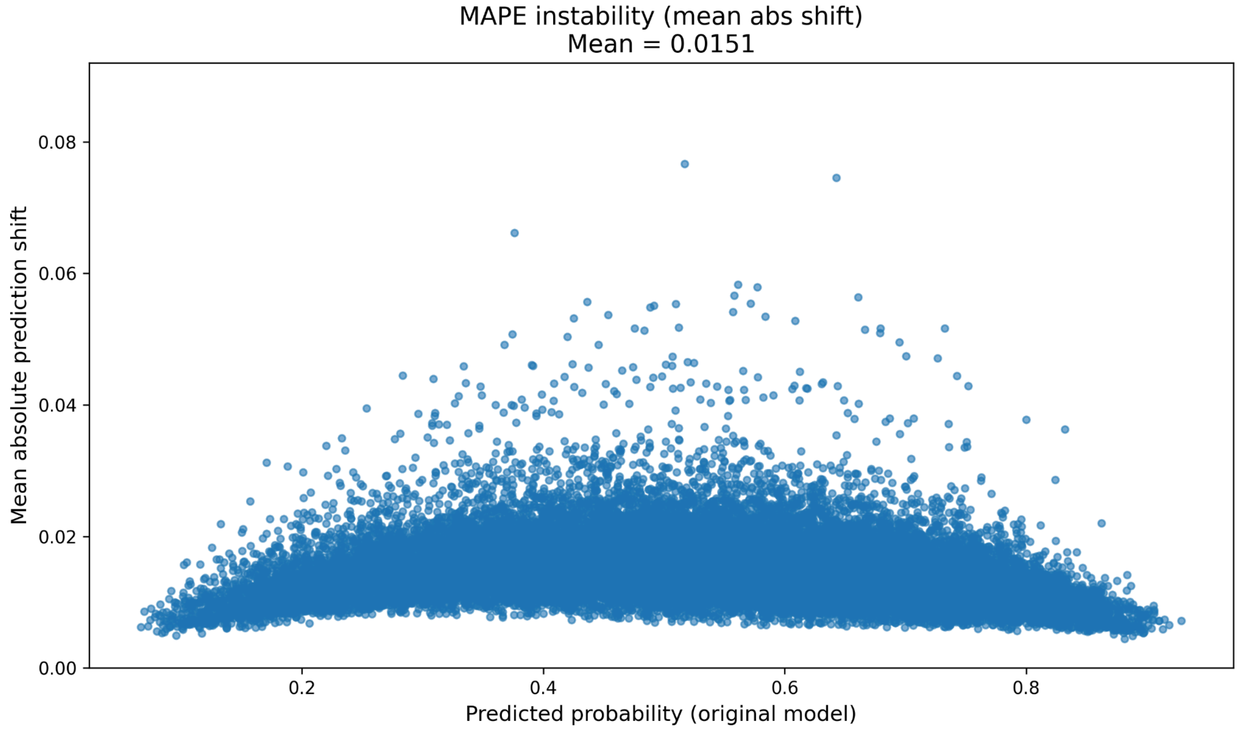


- Optimism

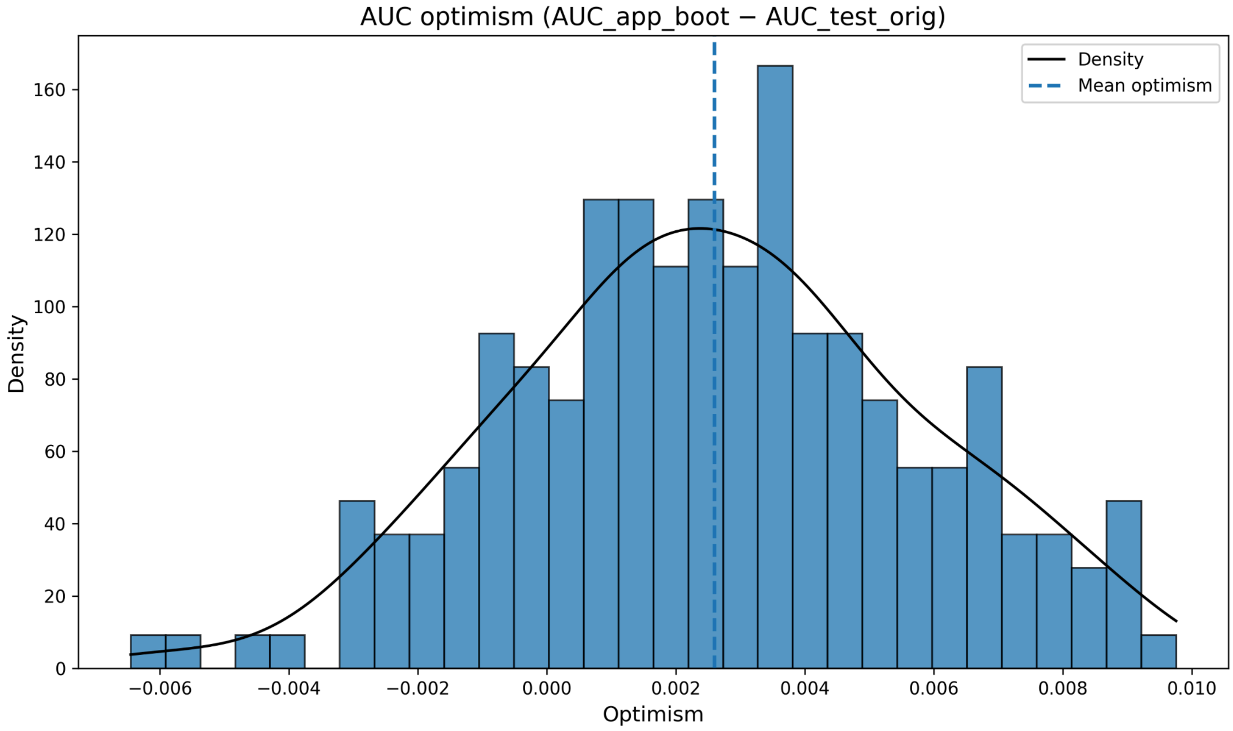

- AUC

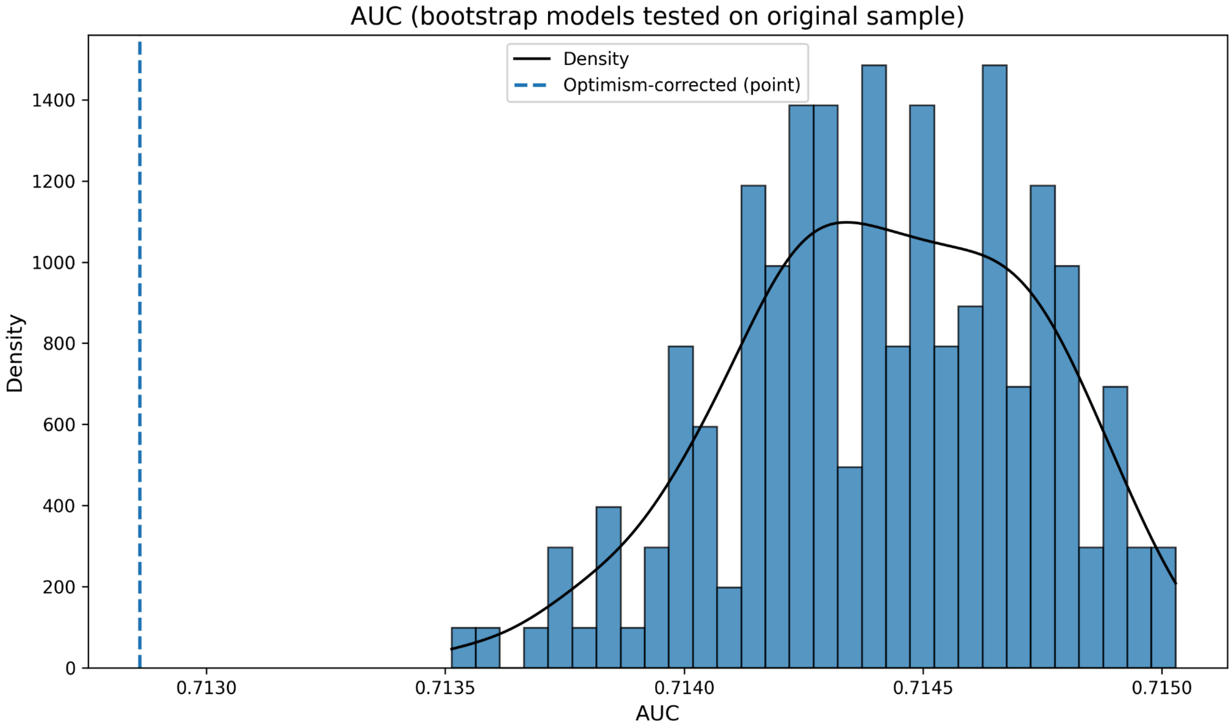


- Sensitivity

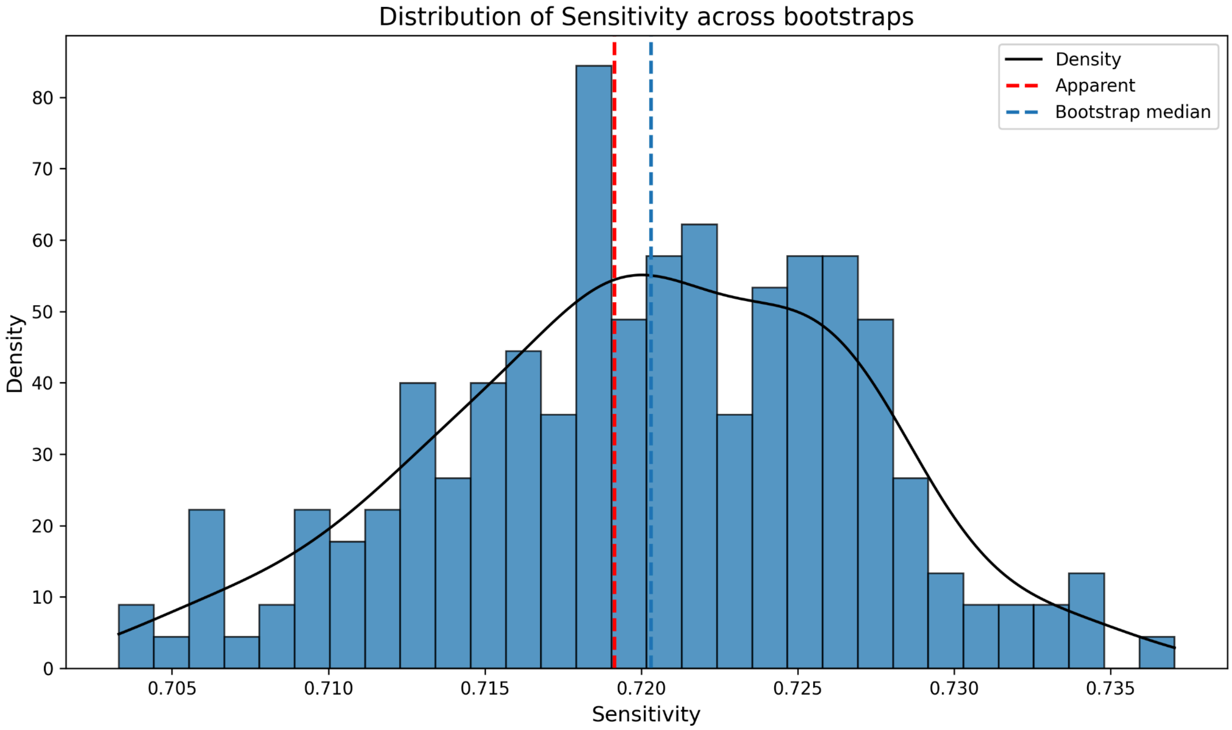

- Specificity

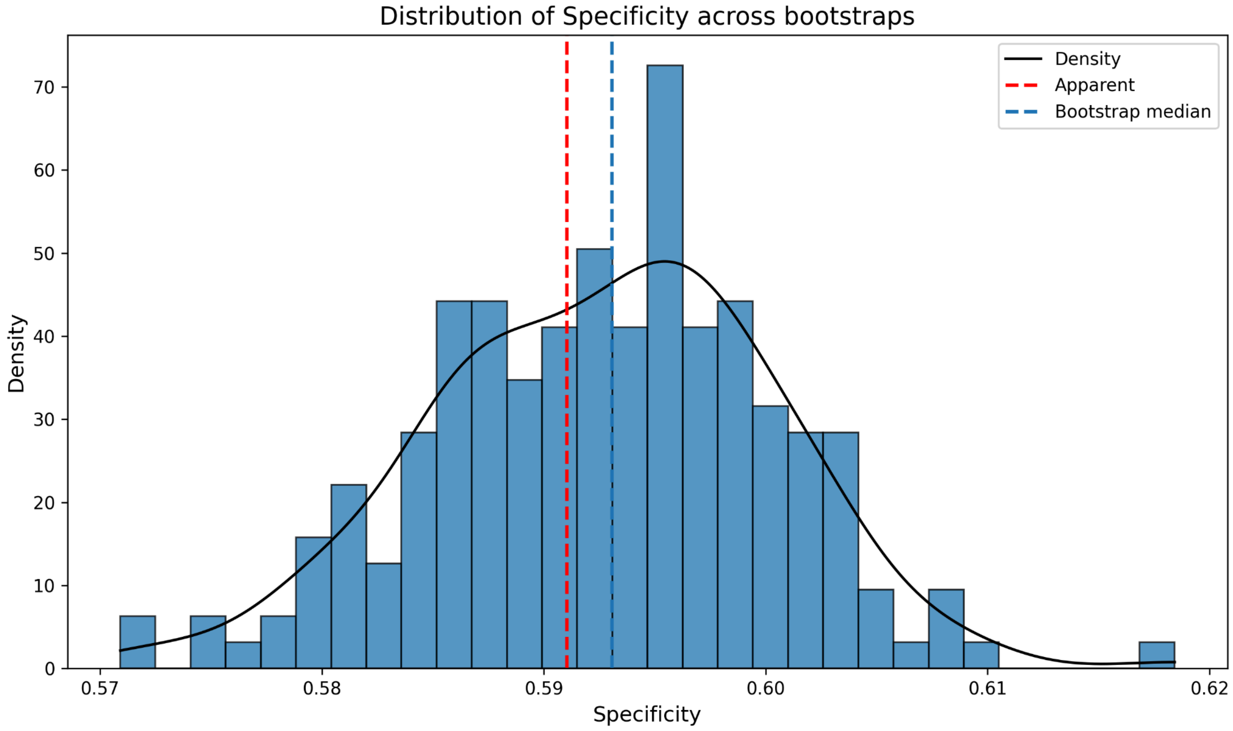


- Brier score

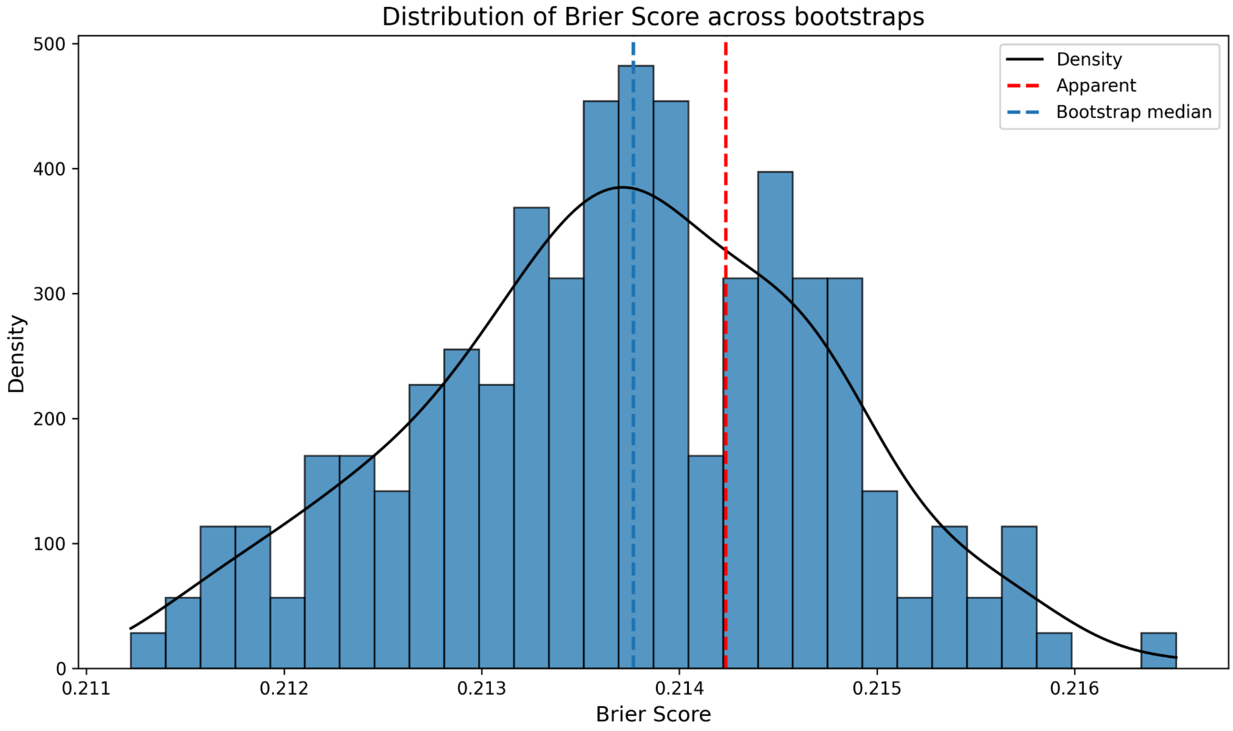

**c) Reliable recovery**

- Calibration curve

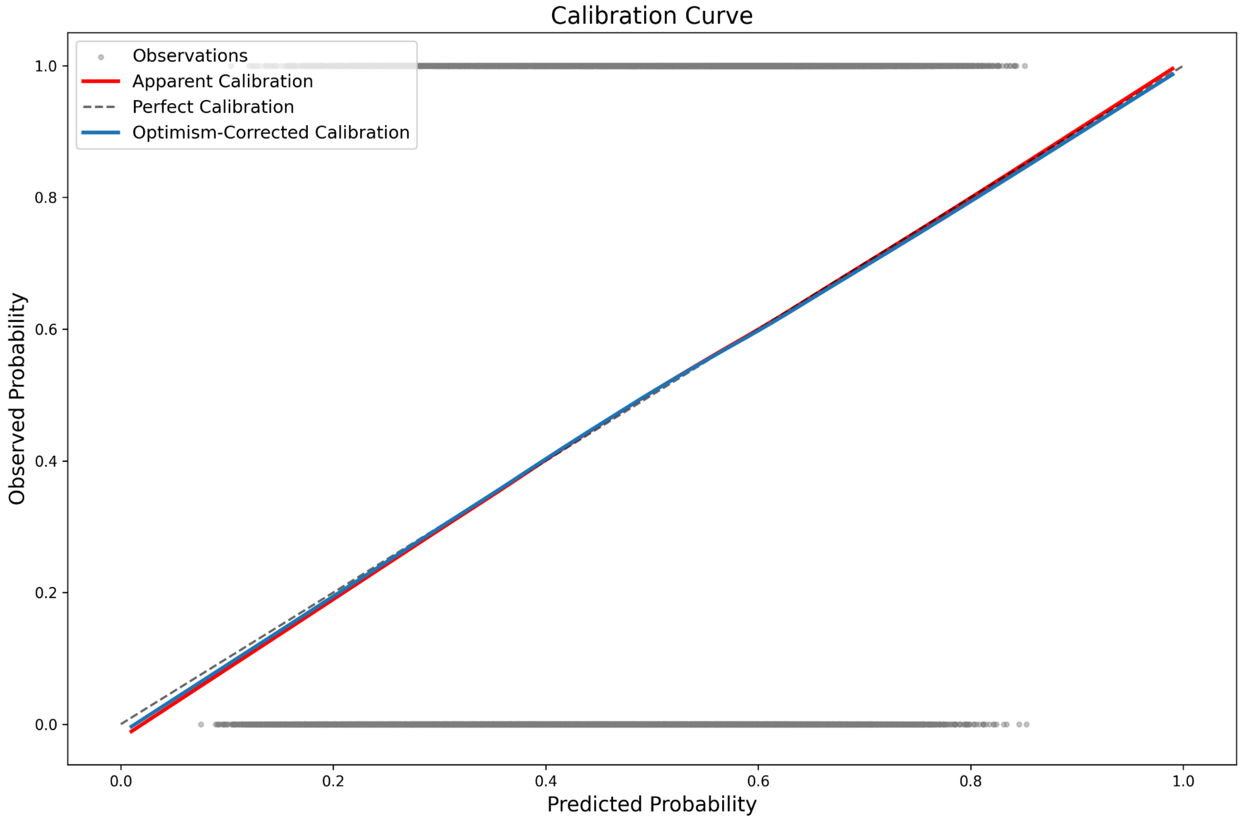

- Prediction instability

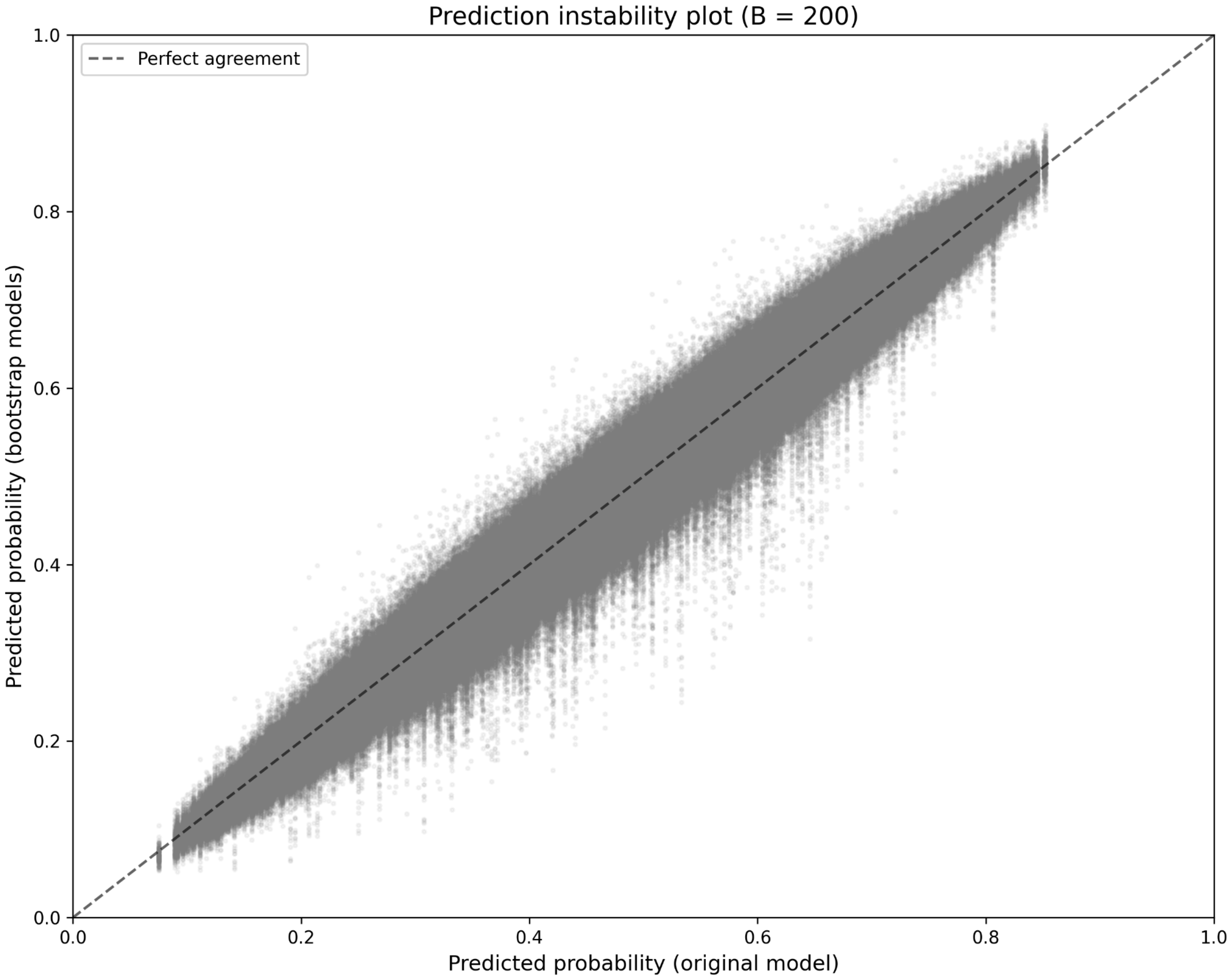

- Calibration instability

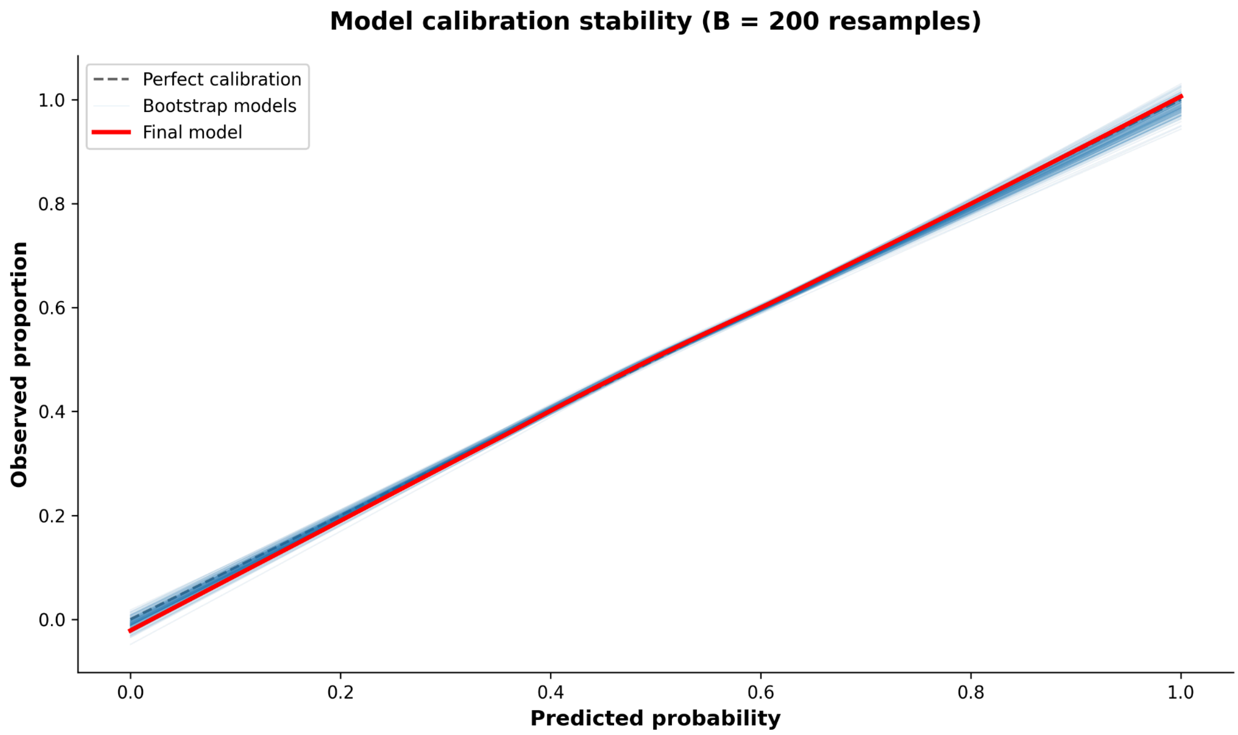

- MAPE

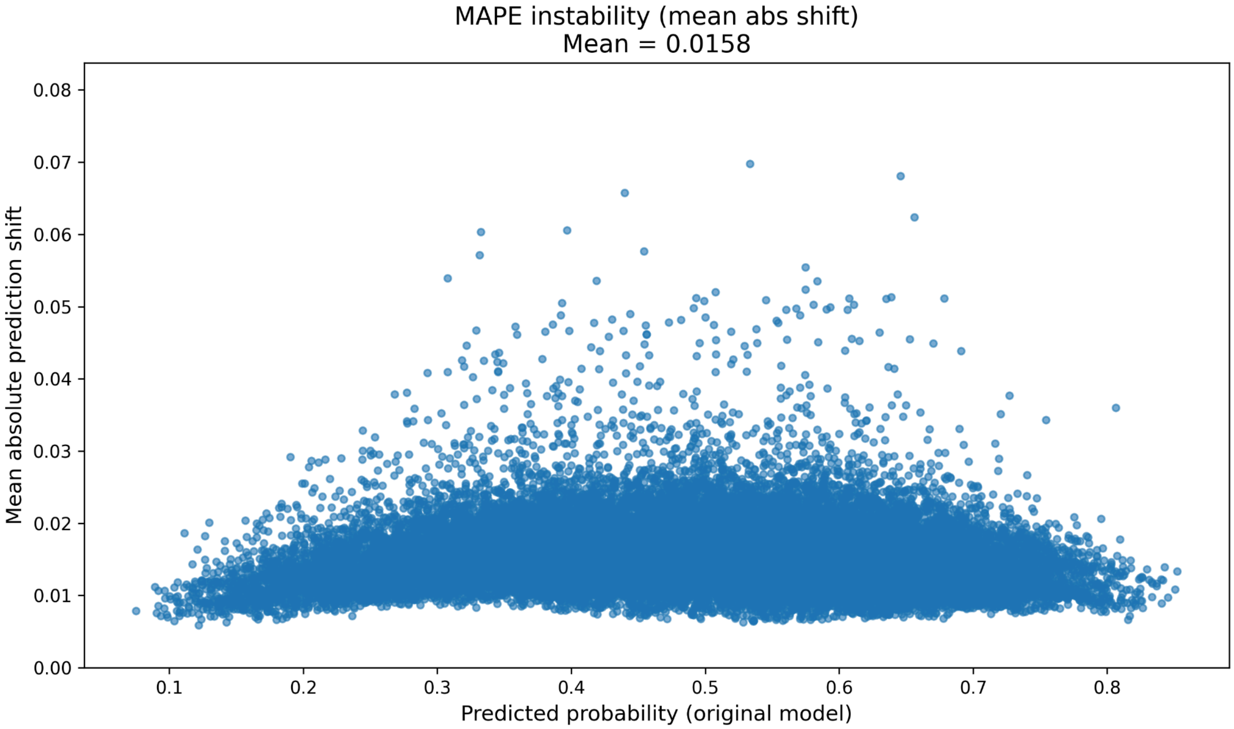


- Optimism

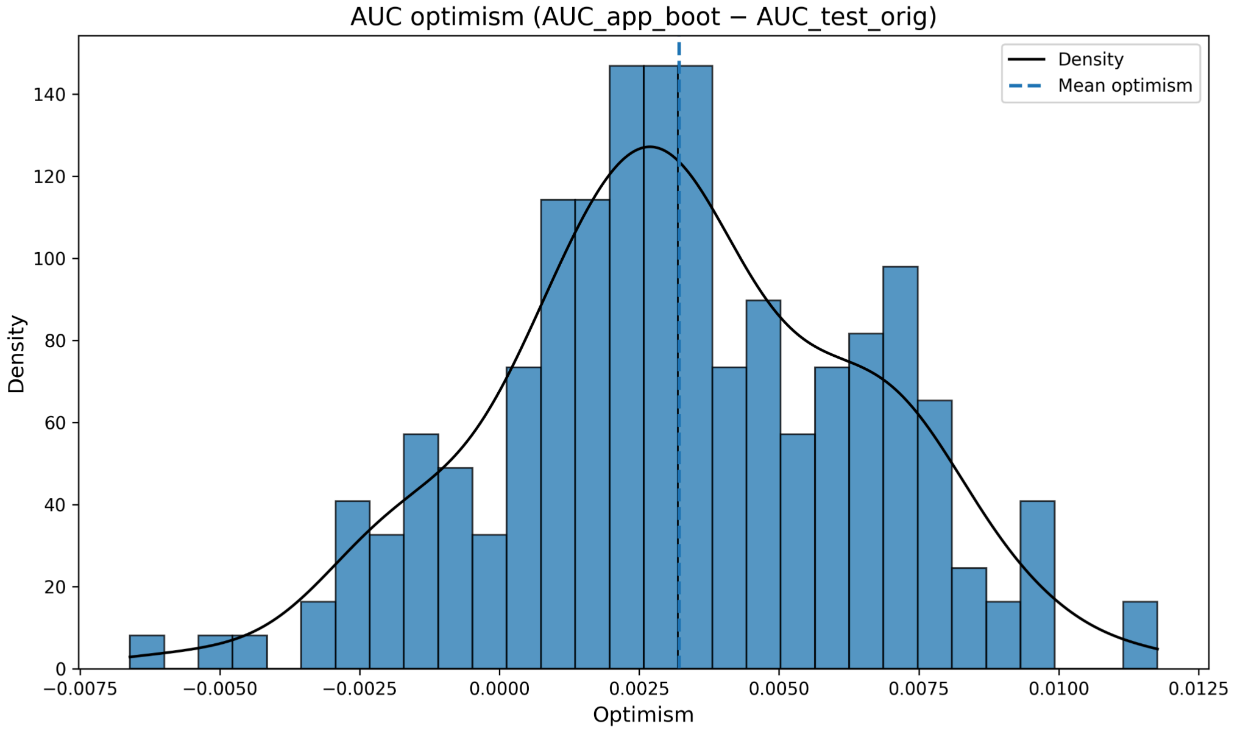

- AUC

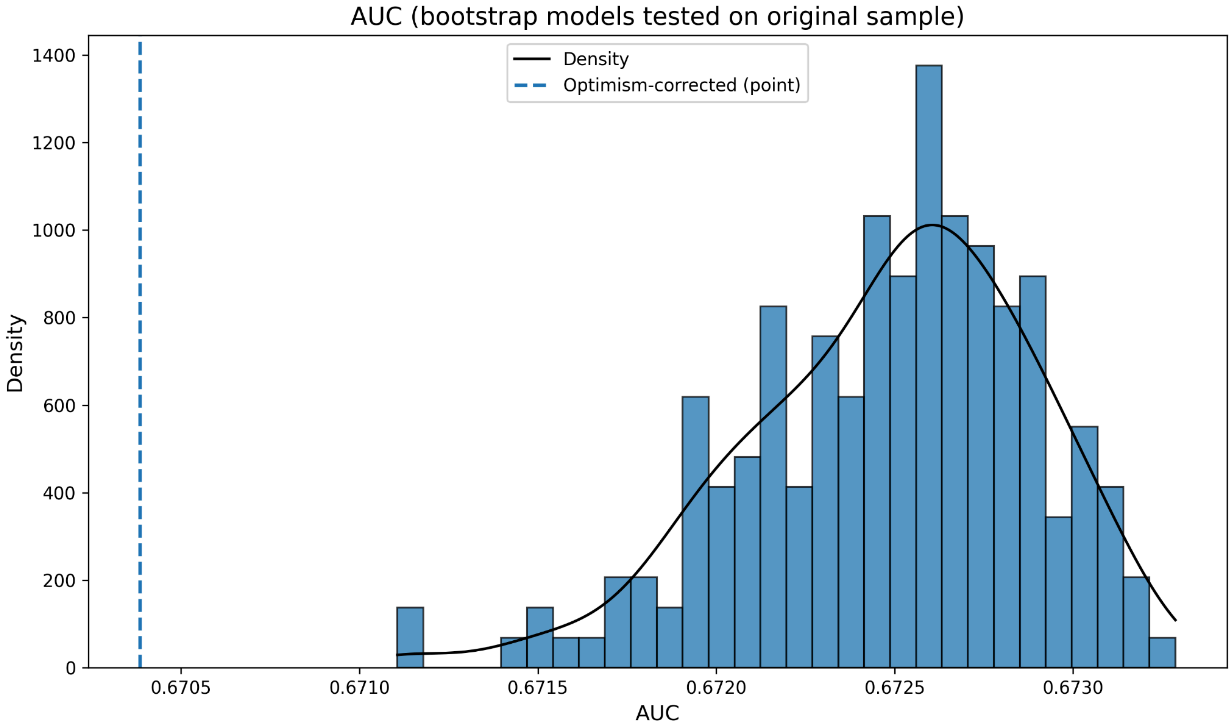


- Sensitivity

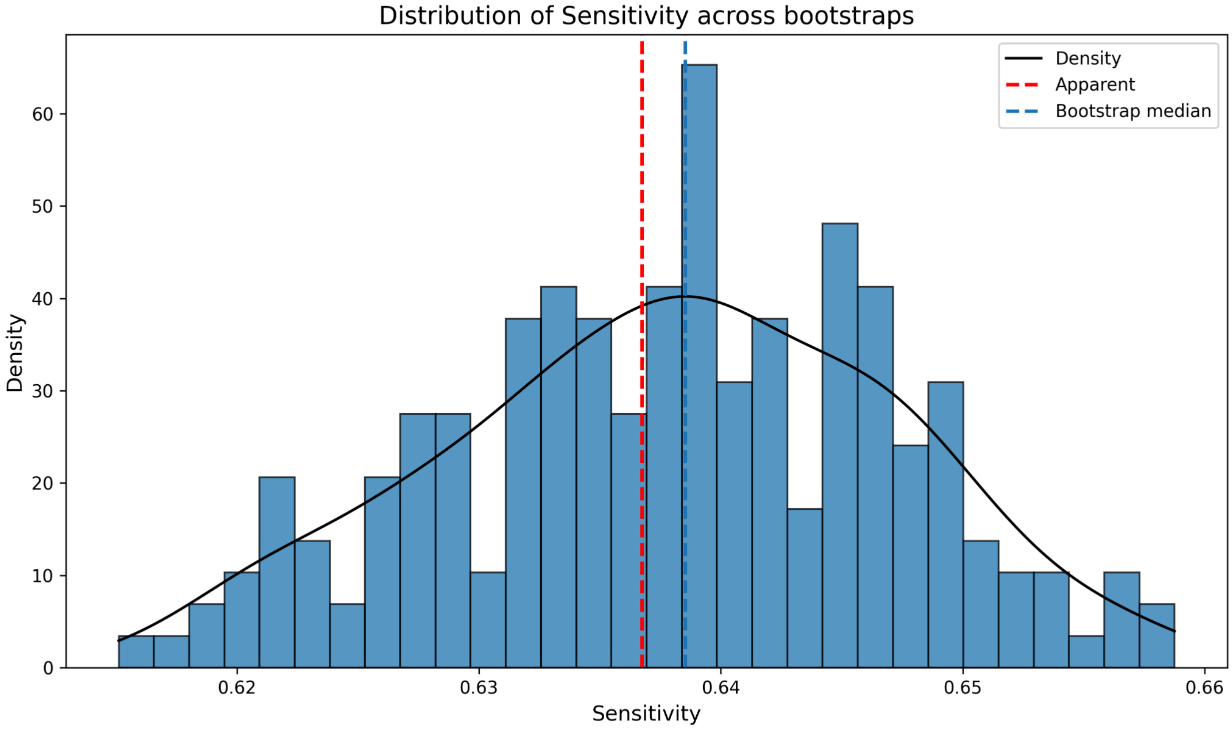

- Specificity

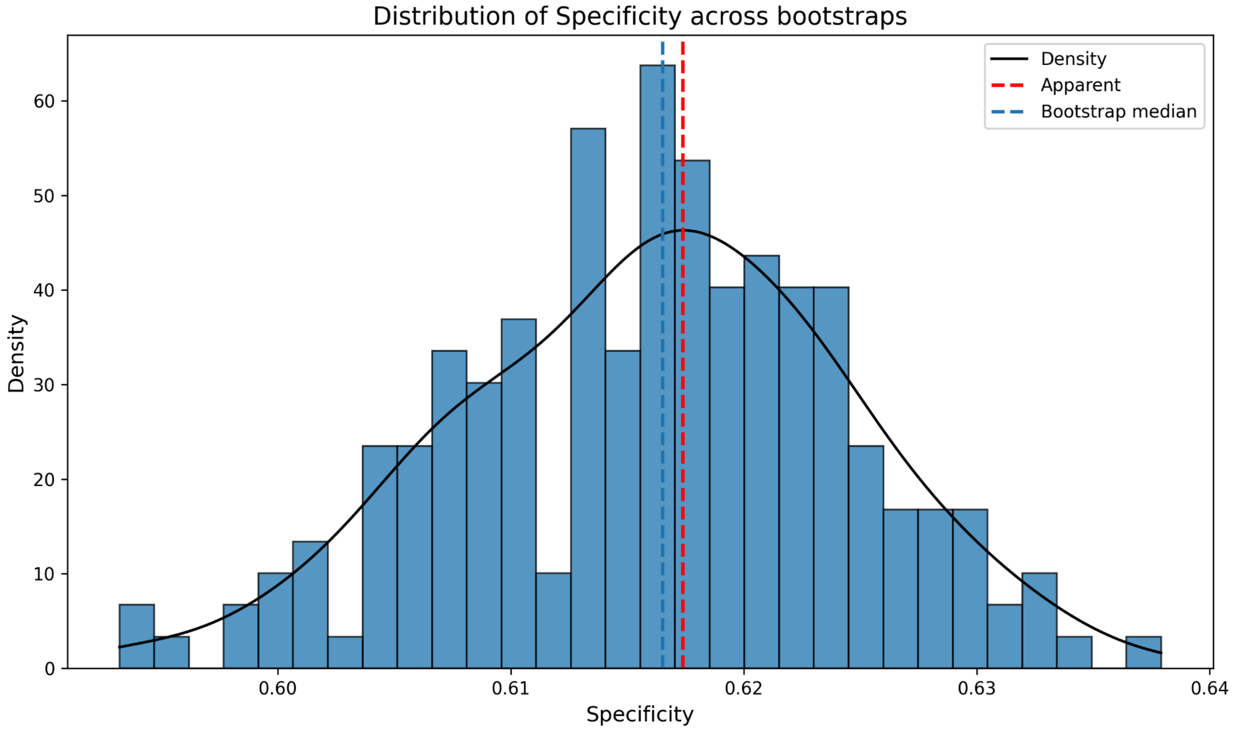


- Brier score

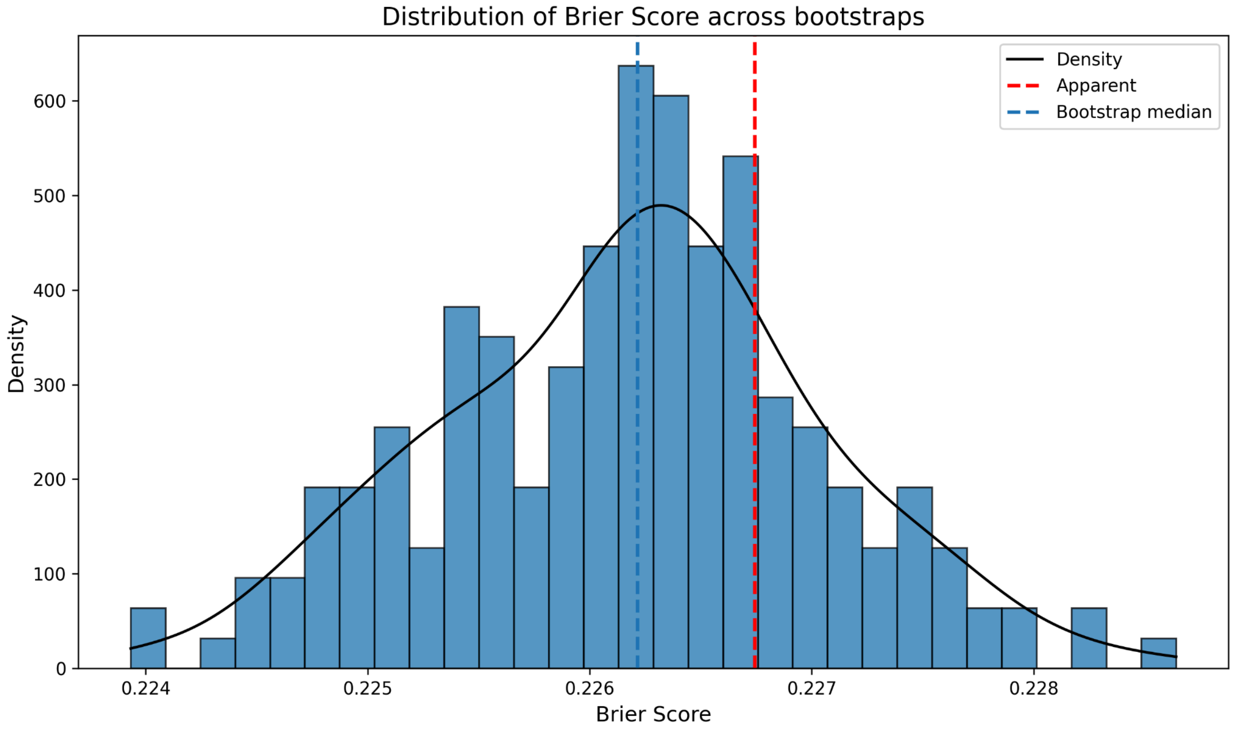

**Figure 8: Bootstrap results for functional impairment outcome**

- Calibration curve

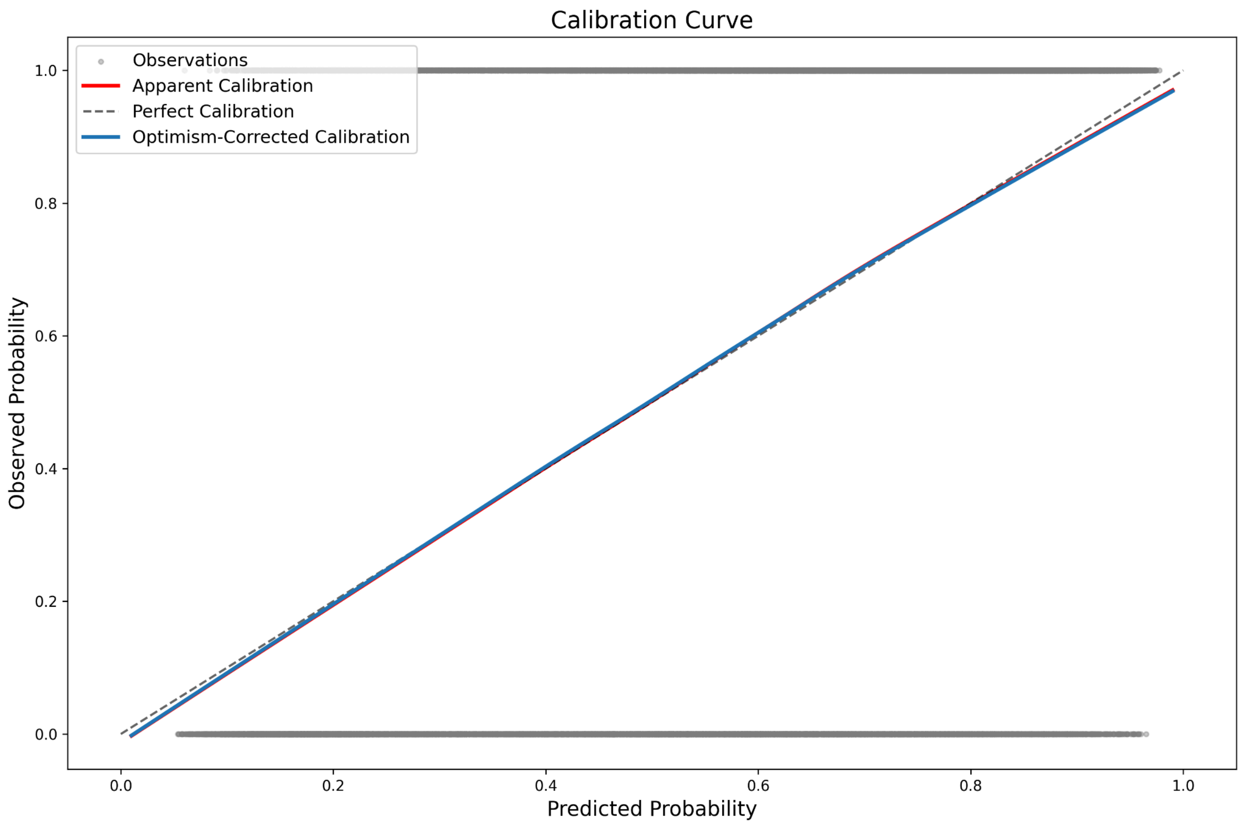

- Prediction instability

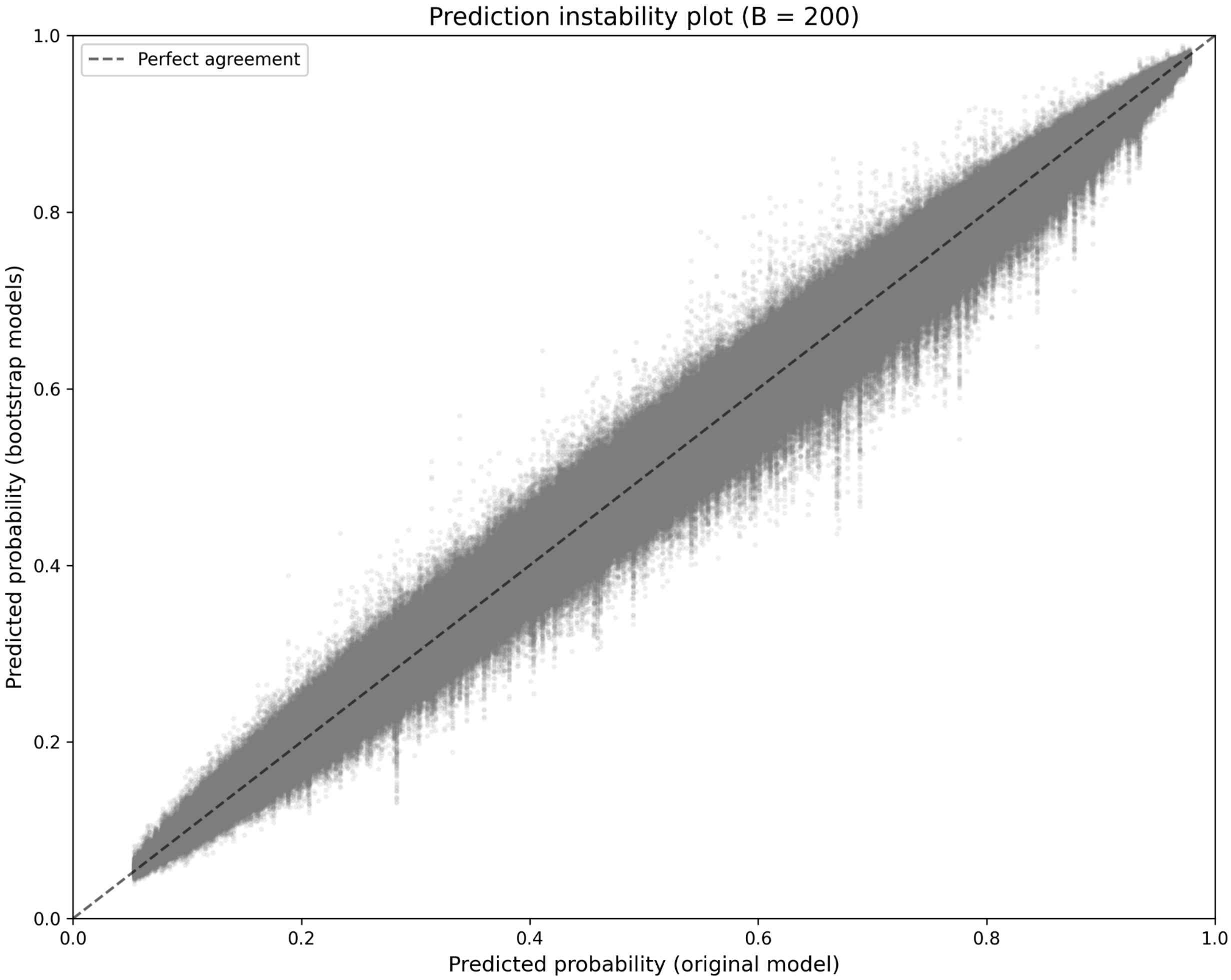

- Calibration instability

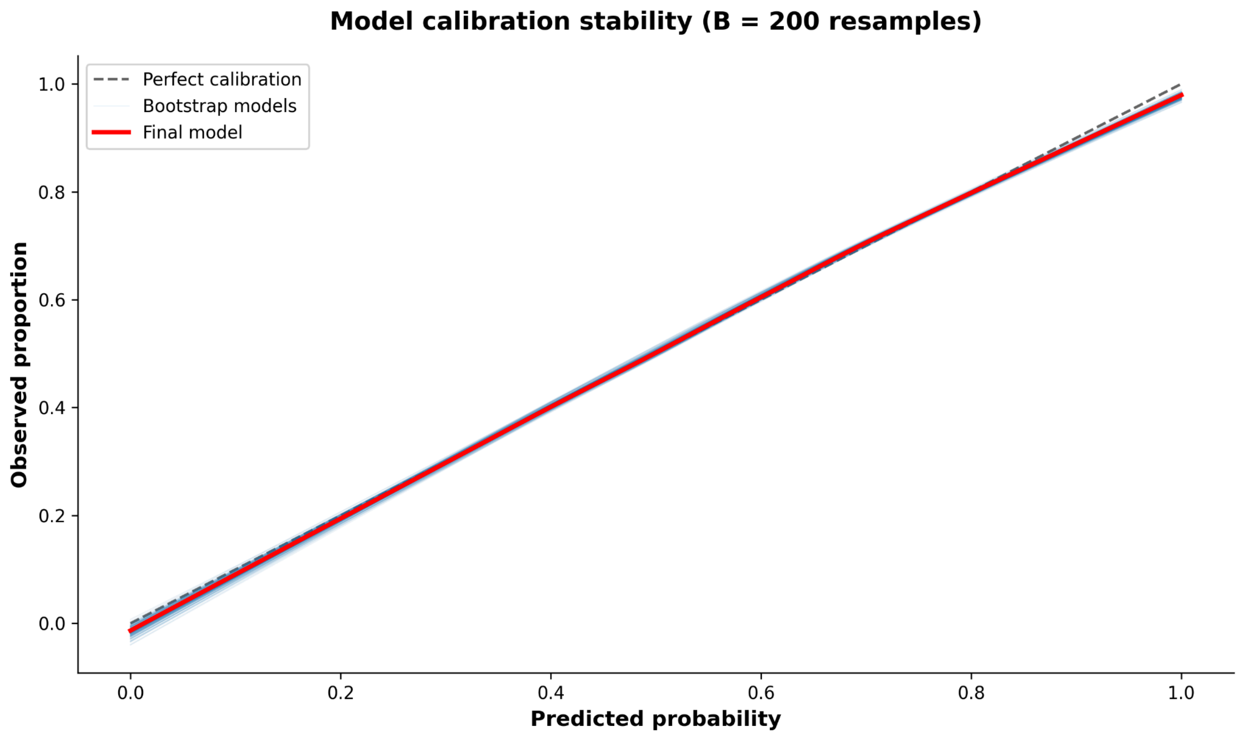

- MAPE

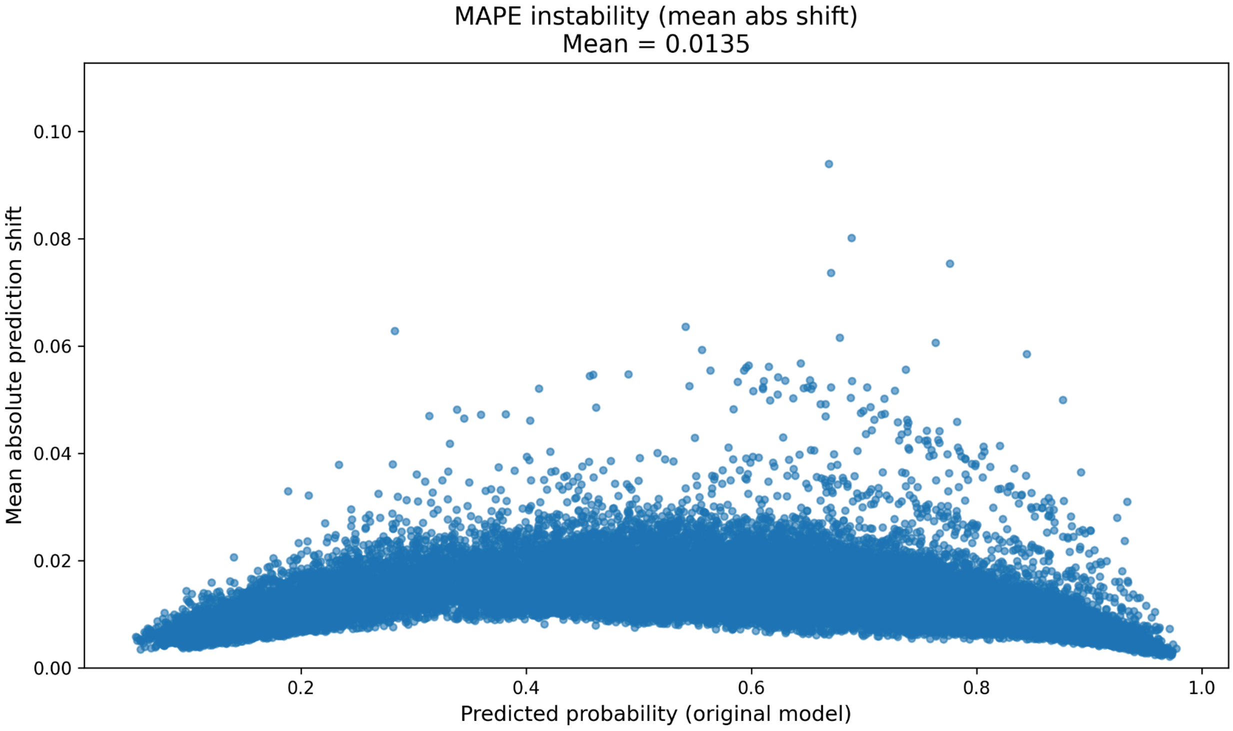


- Optimism

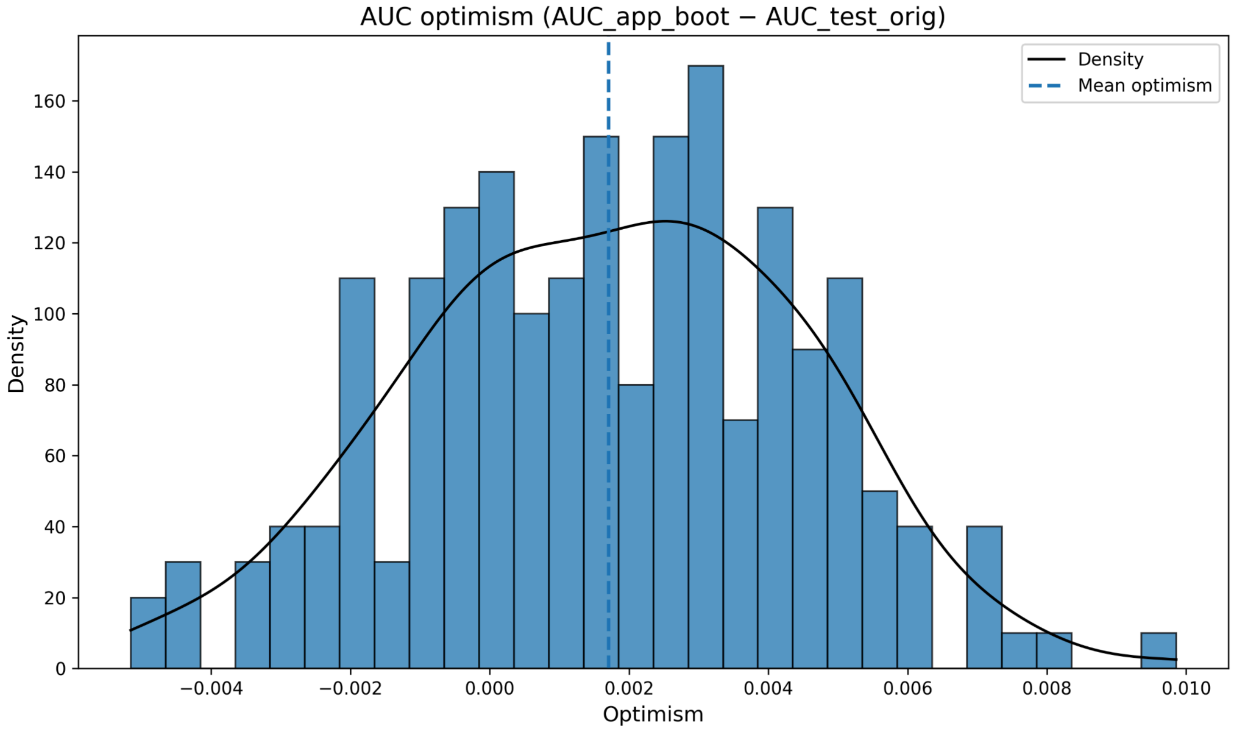

- AUC

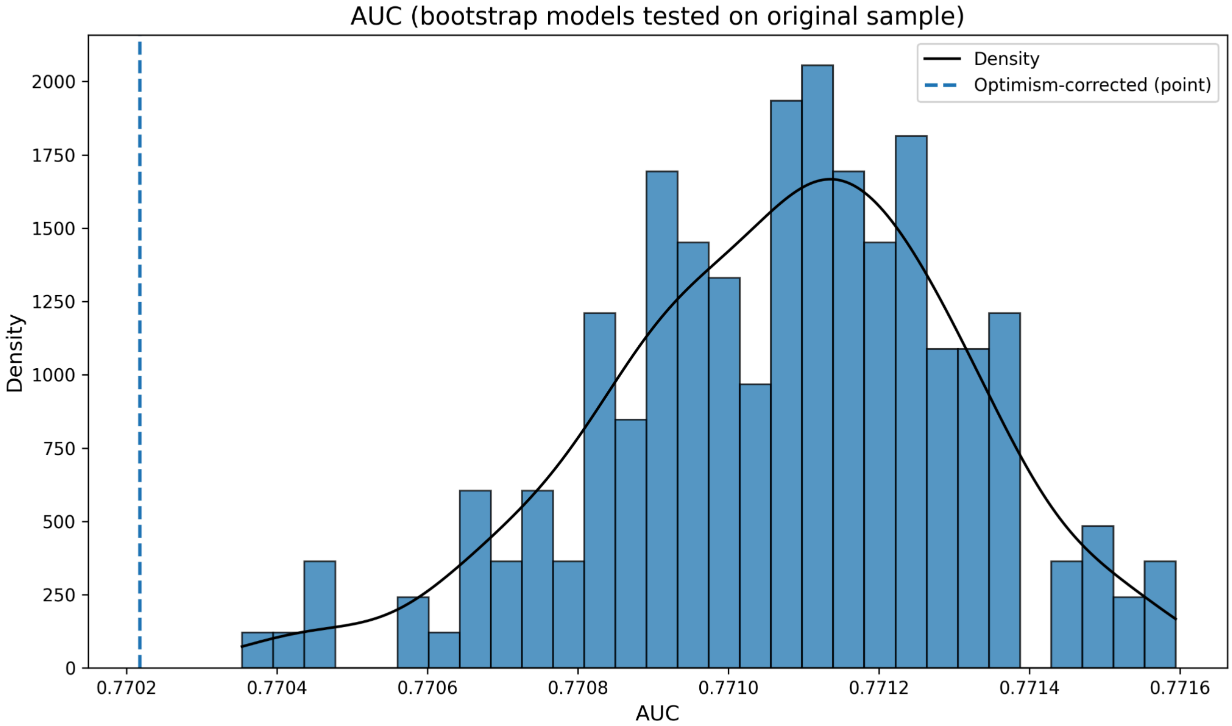


- Sensitivity

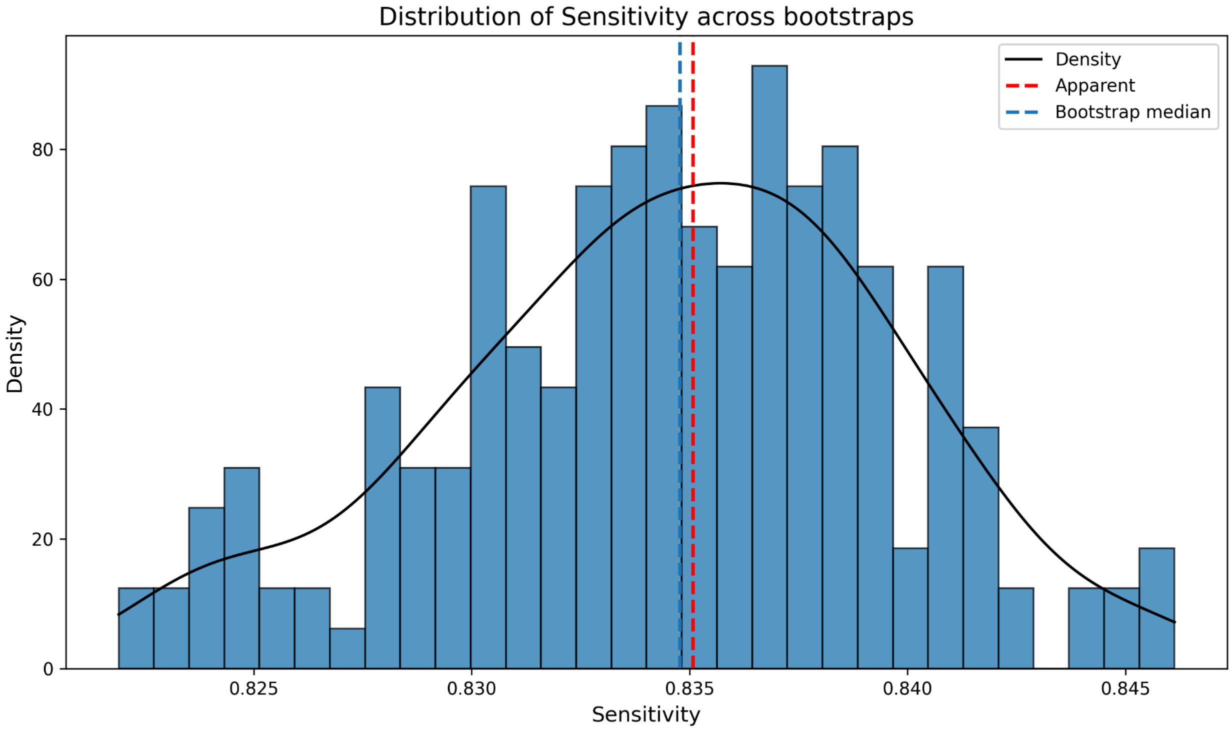

- Specificity

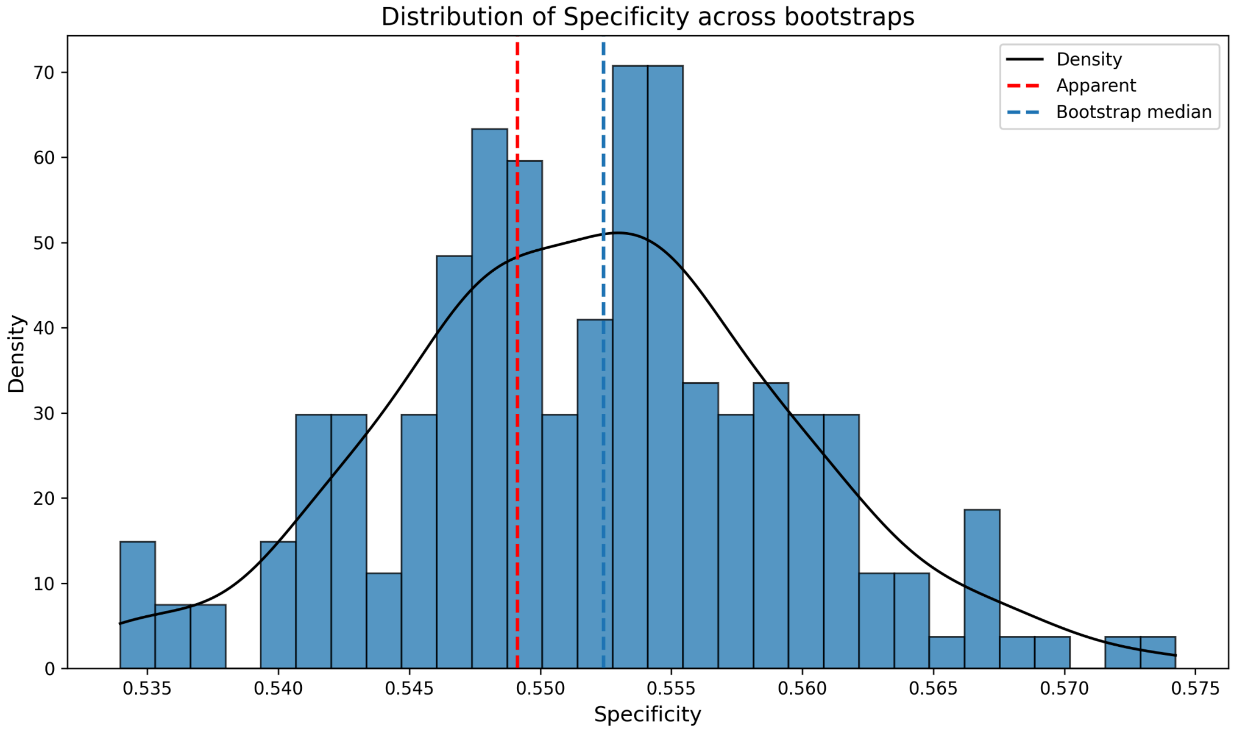


- Brier score

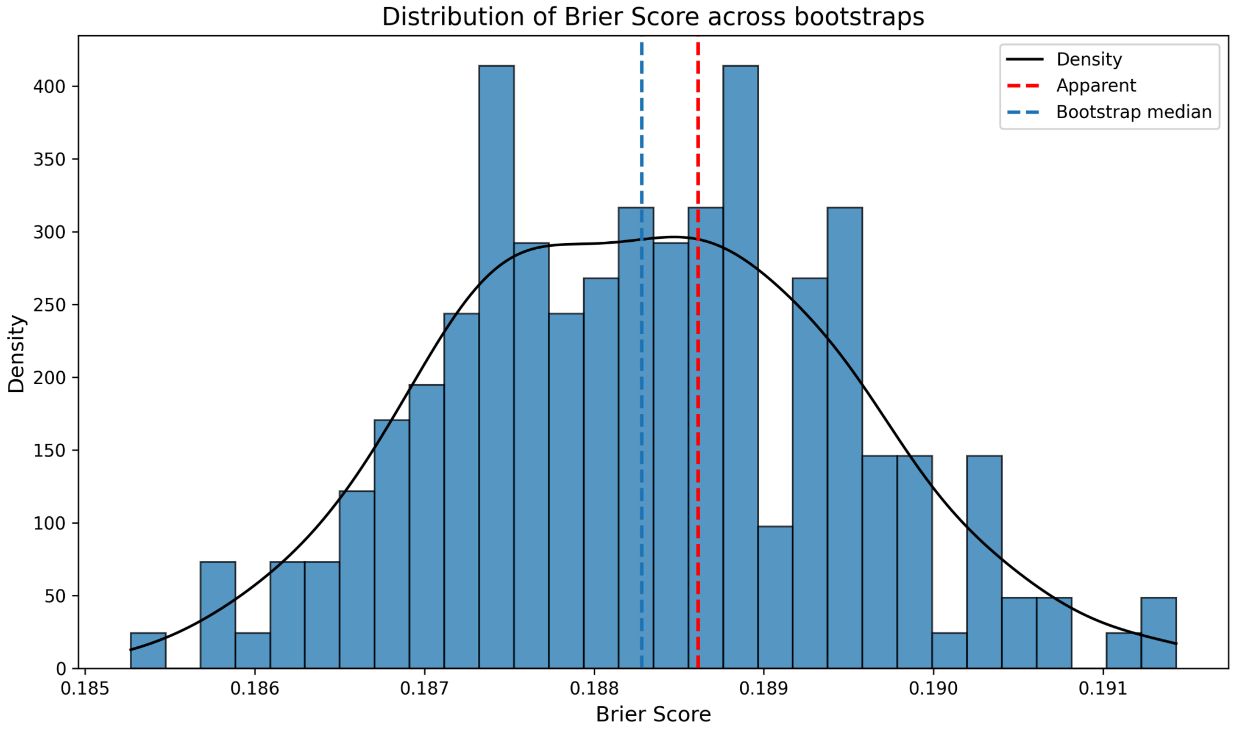

**Table 5: Prediction performance metrics for LASSO models of depression, anxiety, and functional impairment outcomes**

| **January 2018- August 2024** | | | | | | | | | |
|---|---|---|---|---|---|---|---|---|---|
| Outcome | N | **Optimism-corrected bootstrap metric** | | | **Median bootstrap metric **** | | | | |
| | | AUC * | Calibration Intercept | Calibration Slope | Sensitivity | Specificity | PPV | NPV | Brier Score |
| *Depression* | | | | | | | | | |
| **Reliable improvement** | 30,999 | 0.76 (0.76–0.76) | 0.00 | 0.97 | 0.76 (0.74–0.77) | 0.63 (0.61–0.64) | 0.65 (0.63–0.66) | 0.74 (0.72–0.75) | 0.20 (0.19–0.20) |
| **Recovery** | 23,283 | 0.72 (0.72–0.72) | 0.02 | 0.99 | 0.77 (0.76–0.78) | 0.54 (0.53–0.56) | 0.68 (0.67–0.69) | 0.65 (0.64–0.67) | 0.21 (0.21–0.21) |
| **Reliable recovery** | 23,283 | 0.63 (0.63–0.64) | 0.00 | 0.93 | 0.55 (0.53–0.57) | 0.64 (0.62–0.65) | 0.57 (0.55–0.59) | 0.62 (0.60–0.63) | 0.23 (0.23–0.24) |
| *Anxiety* | | | | | | | | | |
| **Reliable improvement** | 30,999 | 0.75 (0.75–0.75) | -0.01 | 1.00 | 0.86 (0.85–0.87) | 0.50 (0.48–0.51) | 0.70 (0.69–0.71) | 0.72 (0.70–0.74) | 0.19 (0.19–0.20) |
| **Recovery** | 25,101 | 0.71 (0.71 –0.72) | 0.00 | 1.00 | 0.72 (0.71–0.74) | 0.59 (0.58–0.60) | 0.66 (0.66–0.68) | 0.65 (0.64–0.67) | 0.21 (0.21–0.22) |
| **Reliable recovery** | 25,101 | 0.67 (0.67–0.67) | -0.00 | 1.00 | 0.64 (0.62–0.66) | 0.62 (0.60–0.63) | 0.62 (0.60–0.63) | 0.64 (0.62–0.66) | 0.23 (0.22–0.23) |
| *Functional impairment* | | | | | | | | | |
| **Functional impairment***** | 30,020 | 0.77 (0.77–0.77) | -0.00 | 1.00 | 0.83 (0.83–0.84) | 0.55 (0.54–0.56) | 0.72 (0.72–0.73) | 0.70 (0.70–0.72) | 0.19 (0.19–0.19) |

* Optimism-corrected AUCs are reported with 95% percentile intervals derived from the bootstrap distribution of optimism estimates (2.5th–97.5th percentiles).
** Median and 95% percentile interval (2.5th–97.5th percentiles) across bootstrap iterations
*** Defined as scoring >10 on the Work and Social Adjustment Scale (WSAS)

For each outcome, the table reports the sample size (N), optimism-corrected AUC with 95% bootstrap percentile intervals (2.5th–97.5th percentiles), optimism-corrected calibration intercept and slope, and median bootstrap estimates for sensitivity, specificity, PPV, NPV, and Brier score with corresponding percentile intervals. All metrics were obtained using nested bootstrap resampling and are rounded to two decimal places. AUC, area under the curve; PPV, positive predictor value; NPV, negative predictor value.

**Table 6: Prediction performance metrics for random forest models of depression, anxiety, and functional impairment outcomes**

| **January 2018- August 2024** | | | | | | | | | |
|---|---|---|---|---|---|---|---|---|---|
| Outcome | N | **Optimism-corrected bootstrap metric** | | | **Median bootstrap metric **** | | | | |
| | | AUC * | Calibration Intercept | Calibration Slope | Sensitivity | Specificity | PPV | NPV | Brier Score |
| *Depression* | | | | | | | | | |
| **Reliable improvement** | 30,999 | 0.79 (0.73–0.85) | -0.07 | 1.16 | 0.89 (0.88–0.91) | 0.74 (0.72–0.76) | 0.77 (0.76–0.79) | 0.87 (0.86–0.88) | 0.15 (0.15–0.15) |
| **Recovery** | 23,283 | 0.75 (0.68–0.81) | -0.19 | 1.36 | 0.88 (0.87–0.89) | 0.66 (0.64–0.68) | 0.77 (0.76–0.78) | 0.81 (0.79–0.83) | 0.17 (0.16–0.17) |
| **Reliable recovery** | 23,283 | 0.73 (0.62–0.84) | -0.23 | 1.54 | 0.83 (0.82–0.86) | 0.81 (0.78–0.83) | 0.79 (0.77–0.81) | 0.84 (0.82–0.86) | 0.17 (0.16–0.17) |
| *Anxiety* | | | | | | | | | |
| **Reliable improvement** | 30,999 | 0.76 (0.72–0.83) | -0.06 | 1.12 | 0.94 (0.93–0.95) | 0.61 (0.59–0.62) | 0.76 (0.75–0.77) | 0.90 (0.88–0.92) | 0.15 (0.15–0.15) |
| **Recovery** | 25,101 | 0.74 (0.67–0.80) | -0.16 | 1.30 | 0.83 (0.82–0.85) | 0.70 (0.68–0.72) | 0.75 (0.73–0.76) | 0.79 (0.77–0.81) | 0.17 (0.17–0.17) |
| **Reliable recovery** | 25,101 | 0.71 (0.63–0.79) | -0.20 | 1.44 | 0.80 (0.78–0.82) | 0.73 (0.71–0.75) | 0.75 (0.73–0.77) | 0.78 (0.76–0.80) | 0.18 (0.18–0.19) |
| *Functional impairment* | | | | | | | | | |
| **Functional impairment***** | 30,020 | 0.79 (0.73–0.85) | -0.05 | 1.10 | 0.92 (0.91–0.93) | 0.66 (0.63–0.68) | 0.79 (0.78–0.80) | 0.85 (0.83–0.87) | 0.14 (0.14–0.14) |

* Optimism-corrected AUCs are reported with 95% percentile intervals derived from the bootstrap distribution of optimism estimates (2.5th–97.5th percentiles).
** Median and 95% percentile interval (2.5th–97.5th percentiles) across bootstrap iterations
*** Defined as scoring >10 on the Work and Social Adjustment Scale (WSAS)

For each outcome, the table reports the sample size (N), optimism-corrected AUC with 95% bootstrap percentile intervals (2.5th–97.5th percentiles), optimism-corrected calibration intercept and slope, and median bootstrap estimates for sensitivity, specificity, PPV, NPV, and Brier score with corresponding percentile intervals. All metrics were obtained using nested bootstrap resampling and are rounded to two decimal places. AUC, area under the curve; PPV, positive predictor value; NPV, negative predictor value.

**Table 7: Prediction performance metrics for gradient boost models of depression, anxiety, and functional impairment outcomes**

| January 2018- August 2024 | | | | | | | | | |
|---|---|---|---|---|---|---|---|---|---|
| Outcome | N | **Optimism-corrected bootstrap metric** | | | **Median bootstrap metric **** | | | | |
| | | AUC * | Calibration Intercept | Calibration Slope | Sensitivity | Specificity | PPV | NPV | Brier Score |
| *Depression* | | | | | | | | | |
| **Reliable improvement** | 30,999 | 0.77 (0.75–0.78) | -0.02 | 1.05 | 0.77 (0.76–0.79) | 0.64 (0.63–0.66) | 0.66 (0.65–0.68) | 0.75 (0.73–0.76) | 0.19 (0.19–0.19) |
| **Recovery** | 23,283 | 0.72 (0.70–0.75) | 0.01 | 0.98 | 0.79 (0.78–0.80) | 0.60 (0.58–0.61) | 0.71 (0.70–0.72) | 0.69 (0.67–0.71) | 0.19 (0.19–0.20) |
| **Reliable recovery** | 23,283 | 0.66 (0.63–0.69) | -0.02 | 1.06 | 0.61 (0.59–0.63) | 0.69 (0.67–0.71) | 0.62 (0.60–0.64) | 0.68 (0.66–0.70) | 0.22 (0.22–0.22) |
| *Anxiety* | | | | | | | | | |
| **Reliable improvement** | 30,999 | 0.76 (0.75–0.77) | -0.02 | 1.04 | 0.87 (0.86–0.88) | 0.52 (0.50–0.53) | 0.71 (0.70–0.72) | 0.74 (0.71–0.76) | 0.18 (0.18–0.19) |
| **Recovery** | 25,101 | 0.72 (0.70–0.73) | 0.00 | 1.00 | 0.74 (0.72–0.75) | 0.62 (0.60–0.64) | 0.68 (0.67–0.69) | 0.69 (0.67–0.71) | 0.20 (0.20–0.21) |
| **Reliable recovery** | 25,101 | 0.69 (0.66–0.72) | 0.01 | 0.98 | 0.69 (0.67–0.70) | 0.67 (0.66–0.69) | 0.68 (0.65–0.69) | 0.68 (0.66–0.70) | 0.21 (0.20–0.21) |
| *Functional impairment* | | | | | | | | | |
| **Functional impairment***** | 30,020 | 0.77 (0.76–0.79) | -0.00 | 1.00 | 0.85 (0.84–0.86) | 0.58 (0.56–0.59) | 0.74 (0.73–0.75) | 0.75 (0.73–0.77) | 0.17 (0.17–0.18) |

* Optimism-corrected AUCs are reported with 95% percentile intervals derived from the bootstrap distribution of optimism estimates (2.5th–97.5th percentiles).
** Median and 95% percentile interval (2.5th–97.5th percentiles) across bootstrap iterations
*** Defined as scoring >10 on the Work and Social Adjustment Scale (WSAS)

For each outcome, the table reports the sample size (N), optimism-corrected AUC with 95% bootstrap percentile intervals (2.5th–97.5th percentiles), optimism-corrected calibration intercept and slope, and median bootstrap estimates for sensitivity, specificity, PPV, NPV, and Brier score with corresponding percentile intervals. All metrics were obtained using nested bootstrap resampling and are rounded to two decimal places. AUC, area under the curve; PPV, positive predictor value; NPV, negative predictor value.

**Table 8: Prediction performance metrics for depression, anxiety, and functional impairment outcomes, excluding the COVID-19 period, developed using elastic net logistic regression (sensitivity analysis)**

| **January 2018- August 2024 (excluding March 1, 2020, to December 31, 2021)** | | | | | | | | | |
|---|---|---|---|---|---|---|---|---|---|
| Outcome | N | **Optimism-corrected bootstrap metric** | | | **Median bootstrap metric **** | | | | |
| | | AUC * | Calibration Intercept | Calibration Slope | Sensitivity | Specificity | PPV | NPV | Brier Score |
| *Depression* | | | | | | | | | |
| **Reliable improvement** | 30,999 | 0.75 (0.75–0.76) | 0.00 | 1.00 | 0.75 (0.73–0.77) | 0.63 (0.61–0.65) | 0.65 (0.63–0.67) | 0.73 (0.71–0.76) | 0.20 (0.20–0.20) |
| **Recovery** | 23,283 | 0.72 (0.71–0.72) | 0.00 | 0.99 | 0.77 (0.76–0.78) | 0.54 (0.52–0.56) | 0.68 (0.67–0.69) | 0.65 (0.63–0.67) | 0.21 (0.21–0.21) |
| **Reliable recovery** | 23,283 | 0.63 (0.63–0.64) | -0.00 | 0.98 | 0.54 (0.51–0.57) | 0.65 (0.62–0.67) | 0.58 (0.54–0.60) | 0.62 (0.59–0.64) | 0.23 (0.23–0.24) |
| *Anxiety* | | | | | | | | | |
| **Reliable improvement** | 30,999 | 0.75 (0.74–0.75) | -0.00 | 1.00 | 0.86 (0.85–0.87) | 0.50 (0.48–0.51) | 0.70 (0.69–0.71) | 0.72 (0.70–0.74) | 0.19 (0.19–0.20) |
| **Recovery** | 25,101 | 0.71 (0.71–0.72) | -0.00 | 0.99 | 0.72 (0.70–0.73) | 0.60 (0.58–0.62) | 0.67 (0.65–0.68) | 0.66 (0.63–0.67) | 0.21 (0.21–0.22) |
| **Reliable recovery** | 25,101 | 0.67 (0.66–0.68) | -0.00 | 0.98 | 0.64 (0.62–0.66) | 0.62 (0.61–0.64) | 0.62 (0.61–0.64) | 0.64 (0.62–0.66) | 0.23 (0.22–0.23) |
| *Functional impairment* | | | | | | | | | |
| **Functional impairment***** | 30,020 | 0.77 (0.76–0.77) | 0.00 | 0.99 | 0.84 (0.83–0.85) | 0.54 (0.52–0.55) | 0.72 (0.71–0.72) | 0.71 (0.69–0.73) | 0.19 (0.19–0.19) |

* Optimism-corrected AUCs are reported with 95% percentile intervals derived from the bootstrap distribution of optimism estimates (2.5th–97.5th percentiles).
** Median and 95% percentile interval (2.5th–97.5th percentiles) across bootstrap iterations
*** Defined as scoring >10 on the Work and Social Adjustment Scale (WSAS)

For each outcome, the table reports the sample size (N), optimism-corrected AUC with 95% bootstrap percentile intervals (2.5th–97.5th percentiles), optimism-corrected calibration intercept and slope, and median bootstrap estimates for sensitivity, specificity, PPV, NPV, and Brier score with corresponding percentile intervals. All metrics were obtained using nested bootstrap resampling and are rounded to two decimal places. AUC, area under the curve; PPV, positive predictor value; NPV, negative predictor value.

### Table 9: TRIPOD+AI checklist

| Section/Topic | Item | Development /Evaluation | Cheklist item | Reported on page |
|---|---|---|---|---|
| **TITLE** | | | | |
| *Title* | 1 | D;E | Identify the study as developing or evaluating the performance of a multivariable prediction model, the target population, and the outcome to be predicted | 1 |
| **ABSTRACT** | | | | |
| *Abstract* | 2 | D;E | See TRIPOD+AI for Abstracts checklist | 2 |
| **INTRODUCTION** | | | | |
| *Background* | 3a | D;E | Explain the healthcare context (including whether diagnostic or prognostic) and rationale for developing or evaluating the prediction model, including references to existing models | 3/4 |
| | 3b | D;E | Describe the target population and the intended purpose of the prediction model in the context of the care pathway, including its intended users (e.g., healthcare professionals, patients, public) | 3/4 |
| | 3c | D;E | Describe any known health inequalities between sociodemographic groups | 3/4 |
| *Objectives* | 4 | D;E | Specify the study objectives, including whether the study describes the development or validation of a prediction model (or both) | 4 |
| **METHODS** | | | | |
| *Data* | 5a | D;E | Describe the sources of data separately for the development and evaluation datasets (e.g., randomised trial, cohort, routine care or registry data), the rationale for using these data, and representativeness of the data | 5 |
| | 5b | D;E | Specify the dates of the collected participant data, including start and end of participant accrual; and, if applicable, end of follow-up | 5 |
| *Participants* | 6a | D;E | Specify key elements of the study setting (e.g., primary care, secondary care, general population) including the number and location of centres | 5 |
| | 6b | D;E | Describe the eligibility criteria for study participants | 5 |
| | 6c | D;E | Give details of any treatments received, and how they were handled during model development or evaluation, if relevant | 5-6 |
| *Data preparation* | 7 | D;E | Describe any data pre-processing and quality checking, including whether this was similar across relevant sociodemographic groups | 5 |
| *Outcome* | 8a | D;E | Clearly define the outcome that is being predicted and the time horizon, including how and when assessed, the rationale for choosing this outcome, and whether the method of outcome assessment is consistent across sociodemographic groups | 5-6 |
| | 8b | D;E | If outcome assessment requires subjective interpretation, describe the qualifications and demographic characteristics of the outcome assessors | NA |
| | 8c | D;E | Report any actions to blind assessment of the outcome to be predicted | NA |
| *Predictors* | 9a | D | Describe the choice of initial predictors (e.g., literature, previous models, all available predictors) and any pre-selection of predictors before model building | 6 |
| | 9b | D;E | Clearly define all predictors, including how and when they were measured (and any actions to blind assessment of predictors for the outcome and other predictors) | 6 |
| | 9c | D;E | If predictor measurement requires subjective interpretation, describe the qualifications and demographic characteristics of the predictor assessors | NA |
| *Sample size* | 10 | D;E | Explain how the study size was arrived at (separately for development and evaluation), and justify that the study size was sufficient to answer the research question. Include details of any sample size calculation | 5 |
| *Missing data* | 11 | D;E | Describe how missing data were handled. Provide reasons for omitting any data | 6 |
| *Analytical methods* | 12a | D | Describe how the data were used (e.g., for development and evaluation of model performance) in the analysis, including whether the data were partitioned, considering any sample size requirements | 6-7-8 |
| | 12b | D | Depending on the type of model, describe how predictors were handled in the analyses (functional form, rescaling, transformation, or any standardisation). | 6 |

| | | | | |
|---|---|---|---|---|
| | 12c | D | Specify the type of model, rationale[2], all model-building steps, including any hyperparameter tuning, and method for internal validation | 6-7-8 |
| | 12d | D;E | Describe if and how any heterogeneity in estimates of model parameter values and model performance was handled and quantified across clusters (e.g., hospitals, countries). See TRIPOD-Cluster for additional considerations[3] | NA |
| | 12e | D;E | Specify all measures and plots used (and their rationale) to evaluate model performance (e.g., discrimination, calibration, clinical utility) and, if relevant, to compare multiple models | 7-8 |
| | 12f | E | Describe any model updating (e.g., recalibration) arising from the model evaluation, either overall or for particular sociodemographic groups or settings | NA |
| | 12g | E | For model evaluation, describe how the model predictions were calculated (e.g., formula, code, object, application programming interface) | 6-7-8 |
| *Class imbalance* | 13 | D;E | If class imbalance methods were used, state why and how this was done, and any subsequent methods to recalibrate the model or the model predictions | NA |
| *Fairness* | 14 | D;E | Describe any approaches that were used to address model fairness and their rationale | NA |
| *Model output* | 15 | D | Specify the output of the prediction model (e.g., probabilities, classification). Provide details and rationale for any classification and how the thresholds were identified | 6-7-8 |

Table showing adherence to Transparent Reporting of a multivariable prediction model for Individual Prognosis Or Diagnosis (TRIPOD+AI) checklist

**Table 10: Descriptive statistics for PHQ-9, GAD-7, and WSAS scores at baseline and last session for a) all patients and b) depression or anxiety cases**

**a) All**

| Measure | Timepoint | N | Mean (SD) | Median (IQR) |
|---|---|---|---|---|
| PHQ-9 | Baseline | 30,999 | 14.15 (6.22) | 14 (10-19) |
| | Last session | — | 8.56 (6.35) | 7 (4-12) |
| GAD-7 | Baseline | 30,999 | 12.76 (5.25) | 13 (9-17) |
| | Last session | — | 7.78 (5.59) | 7 (4-11) |
| WSAS | Baseline | 30,020 | 18.10 (9.49) | 18 (11-25) |
| | Last session | — | 13.19 (9.99) | 12 (5-20) |

**b) (N= cases)***

| Measure | Timepoint | N | Mean (SD) | Median (IQR) |
|---|---|---|---|---|
| PHQ-9 | Baseline | 23,283 | 16.84 (4.52) | 16 (13-20) |
| | Last session | — | 9.93 (6.41) | 9 (5-14) |
| GAD-7 | Baseline | 25,101 | 14.63 (3.85) | 15 (11-18) |
| | Last session | — | 8.62 (5.63) | 7 (4-13) |

The tables provide the sample size (N), mean, standard deviation (SD), median, and interquartile range (IQR).

Footnotes:

- Cases refer to participants meeting clinical criteria at baseline for each measure.
- * N/A for WSAS cases: meeting the baseline "caseness" criteria is not required to assess outcomes, making it inappropriate to focus only on this group.

**Table 11: Treatment outcomes of depression, anxiety, and functional impairment**

| Outcome | N Eligible | N Achieved Outcome (% of the sample) |
|---|---|---|
| *Depression* | | |
| **Reliable improvement** | 30,999 | 14,815 (47.78%) |
| **Recovery** | 23,283 | 12,964 (55.68%) |
| **Reliable recovery** | 23,283 | 10,914 (46.86%) |
| *Anxiety* | | |
| **Reliable improvement** | 30,999 | 17,954 (57.91%) |
| **Recovery** | 25,101 | 13,281 (52.92%) |
| **Reliable recovery** | 25,101 | 12,352 (49.20%) |
| *Functional impairment* | | |
| **Functional impairment** | 30,020 | 17,469 (58.17%) |

This table presents the number of eligible participants (N Eligible) and the number and percentage of participants who achieved each treatment outcome (N Achieved Outcome) for depression, anxiety, and functional impairment.

### Supplementary Methods

#### A) Bootstrap Validation Procedure

We applied Harrell's optimism-corrected bootstrap method (Harrell et al., 1996) to estimate out-of-sample model performance and adjust for overfitting. to estimate out-of-sample model performance and adjust for overfitting. A total of 200 bootstrap samples were drawn with replacement from the original dataset. For each iteration, the following steps were performed:

1. A new dataset was created by sampling from the original data with replacement.
2. A new elastic net model was trained on each resampled dataset, with the full modelling pipeline applied within each bootstrap sample, including missing data imputation and re-tuning of hyperparameters, to ensure the model was optimised for each specific sample.
3. The model was then used to generate predictions for both the bootstrap sample and the original dataset to assess apparent and test performance, respectively.
4. Key performance metrics were calculated at each bootstrap iteration, including AUC, sensitivity, specificity, and Brier score.

The optimism-corrected AUC was derived by subtracting the average optimism (the difference between the apparent and test AUCs) from the apparent AUC, known as Harrell's bias-corrected method (Harrell et al., 1996; Iba et al., 2021). The steps are as follows:

1. For each bootstrap iteration, a new model is retrained on a given bootstrap dataset, and the AUC is calculated for the original dataset (not the bootstrap dataset).
2. Each bootstrap iteration gives a "test" AUC based on the predictions made by the retrained model on the original data.
3. For each iteration, optimism is calculated: the difference between the apparent AUC and the test AUC for that iteration.
4. After repeating this for all bootstrap iterations, we compute the average optimism, the mean of these differences.

Median values were reported for sensitivity, specificity and Brier score, along with the upper and lower percentiles (2.5% and 97.5%) from bootstrap iterations (Iba et al., 2021; Noma et al., 2021). Distributions of these metrics per bootstrap are shown in **Figs. 6**, **7**, and **8** for depression, anxiety, and functional impairment outcomes, respectively.

**B) Calibration and Stability Analyses**

1. Calibration assessment

For each bootstrap iteration, we recalculated calibration metrics to estimate the optimism introduced by resampling (Harrell et al., 1996). Calibration was assessed using:

- **Calibration intercept and slope**: estimated using logistic recalibration, by fitting a logistic regression model with the observed outcome as the dependent variable and the logit of the predicted probability as the sole covariate. The intercept reflects systematic over- or under-prediction, while the slope indicates the degree of overfitting or underfitting (Steyerberg, 2019).

- **Flexible calibration curves**: generated using non-parametric locally weighted scatterplot smoothing (LOWESS) to evaluate the alignment between predicted probabilities and observed outcomes across the risk spectrum.

The optimism-corrected calibration curve was derived by computing the difference between the apparent and test LOWESS calibration curves at each bootstrap iteration and subtracting the average optimism across iterations from the apparent calibration curve. Optimism-corrected calibration intercepts and slopes were summarised for each outcome.

2. Prediction and calibration stability

We used the framework described by Riley & Collins (2023) to assess the stability of individual predictions and calibration estimates:

- Prediction stability captures the variability in individual-level predictions across bootstrap models. For each individual, we generated prediction instability plots showing the distribution of predicted values across the 200 bootstrap models (*B* predictions) against their original model prediction. A 95% stability interval, smoothed using a LOWESS curve, was included to aid interpretability.
- Calibration stability was evaluated using calibration instability plots, which overlaid the calibration curves from each bootstrap model alongside the original model's calibration curve. A wider spread of curves indicates a greater risk of miscalibration across resamples.
- We also calculated the Mean Absolute Prediction Error (MAPE) per individual, defined as the average absolute difference between the original prediction and the *B* bootstrap predictions. To explore where prediction instability was most pronounced, we produced MAPE instability plots, which display individual MAPE values plotted against their predicted risk from the original model.

All visualisations for calibration, prediction instability, calibration instability, and MAPE are presented in **Supplementary Figs. 6–8** for depression, anxiety, and functional impairment outcomes.

Harrell, F. E., Lee, K. L., & Mark, D. B. (1996). MULTIVARIABLE PROGNOSTIC MODELS: ISSUES IN DEVELOPING MODELS, EVALUATING ASSUMPTIONS AND ADEQUACY, AND MEASURING AND REDUCING ERRORS. *Statistics in Medicine*, *15*(4), 361–387. https://doi.org/10.1002/(SICI)1097-0258(19960229)15:4%253C361::AID-SIM168%253E3.0.CO;2-4

Iba, K., Shinozaki, T., Maruo, K., & Noma, H. (2021). Re-evaluation of the comparative effectiveness of bootstrap-based optimism correction methods in the development of multivariable clinical prediction models. *BMC Medical Research Methodology*, *21*(1), 9. https://doi.org/10.1186/s12874-020-01201-w

Noma, H., Shinozaki, T., Iba, K., Teramukai, S., & Furukawa, T. A. (2021). Confidence intervals of prediction accuracy measures for multivariable prediction models based on the bootstrap‐based optimism correction methods. *Statistics in Medicine*, *40*(26), 5691–5701. https://doi.org/10.1002/sim.9148

Steyerberg, E. W. (2019). *Clinical Prediction Models: A Practical Approach to Development, Validation, and Updating*. Springer International Publishing. https://doi.org/10.1007/978-3-030-16399-0
